# Associations between early-life mental health and abnormal sleep duration in midlife: findings from a prospective cohort study in Great Britain

**DOI:** 10.1101/2023.12.13.23299897

**Authors:** Thomas E. Metherell, George B. Ploubidis, Darío Moreno-Agostino

## Abstract

**Objectives:** To quantify the association between the state of mental health in childhood and sleep duration at age 46-48 in the 1970 British Cohort Study (BCS70).

**Design:** BCS70 is a prospective birth cohort study of a population-representative sample.

**Setting:** Data collection was conducted in participants’ homes across Great Britain.

**Participants:** All individuals born in one week in April 1970 and resident in the UK are eligible to take part, including those born outside the UK who subsequently immigrated. The study has more than 17,000 participants, of whom 8,581 responded at the last available sweep at age 46-48.

**Main outcome measures:** Average nightly sleep duration at age 46-48, as assessed by five separate methods: self-report, inference from a sleep diary and three algorithms used to process accelerometry data collected using a thigh-worn activPAL™ 3 device.

**Results:** All seven measures of childhood mental ill-health were positively associated with abnormal self-reported sleep duration. Five were associated with abnormal diary-derived estimates and four were associated with abnormal estimates from an accelerometry-based algorithm. Strengths of associations vary. Adjusting for adult mental health symptoms partially attenuated the associations with self-reported sleep duration but not with the more objective estimates.

**Conclusions:** Poor mental health in childhood is associated with abnormal nightly sleep duration at age 46-48. Post-hoc analyses suggest that this association might not be entirely mediated by poor mental health in adulthood. Future research should attempt to replicate these results in diverse samples and assess whether stronger interventions in early-life mental health might have substantial and long-lasting positive impacts on sleep.

**Summary:** **What is already known on this topic**

- The state of a person’s mental health in their early life is associated with a range of adult health outcomes.
- There exists a bidirectional and multifaceted relationship between the state of a person’s mental health and their sleep.
- The longitudinal association between early-life mental health and sleep in adulthood was not well understood.

**What this study adds**

- Our study demonstrates that early-life mental health is associated with an abnormal average nightly sleep duration at age 46 in this sample.
- Associations between early-life mental health and abnormal average nightly sleep at age 46 were not fully mediated by adult mental health.

## Introduction

The state of a person’s mental health in their early life is known to be associated with a range of adult characteristics. The presence of a mental disorder in early life is associated with lower income [1], greater risk of excessive alcohol consumption [2], premature mortality [3] and a higher risk of mental health problems in adulthood [4]. It is therefore critical to understand the nature and extent of associations between early-life mental health difficulties and outcomes later in life, in order to fully appreciate the potential value of early mental health interventions that aim to reduce the incidence of such difficulties. In addition, the rates of probable mental illness among young people are increasing rapidly in countries such as the United Kingdom [5], [6], which makes it ever more important to understand the potential long-term consequences. Sleep is critically important to normal physiological and neurobehavioural functioning, with a nightly duration of less than seven hours being associated with deficits in both domains [7]. Despite this, insufficient sleep is extremely common, with an estimated 29.2 % of US adults reporting sleeping for less than six hours per night in 2012 [8]. Therefore, it is not surprising that chronic sleep deprivation has been raised as a matter of significant public health concern. Sleep deprivation has been linked to neurobehavioural impairments in children [9], premature mortality [10] and a host of other molecular and immune changes that may facilitate the onset of disease [7]. An abnormally long sleep duration has also been linked to mortality and a number of physical health conditions [11], [12]. It follows that controlling the incidence of sleep disturbance in the population may go some way to reducing the burden of various physical and mental health disorders and therefore ought to be prioritised by researchers and policymakers.

The relationship between sleep duration and mental health is likely both bidirectional and multifaceted [13]–[15]. Improving sleep quality is known to positively impact mental health [16] and vice-versa [17]–[19]. There is also evidence to suggest that sleep problems in early life are associated with a later onset of mental health symptoms, which may have a long-lasting deleterious impact [9], [20]. However, the relationship between early-life mental health and sleep later in life is less well understood. Findings have pointed to a link between early-life trauma and sleep across the lifespan in rats [21], and between early-life stress load and sleep in psychiatric outpatients [22], but there is a lack of evidence regarding early-life mental health itself, and from representative human samples. The existence of a long-term relationship between early-life mental health and sleep would imply an impact of early-life mental health symptoms on sleep architecture that persists into adulthood. In accordance with the evidence outlined above, this could suggest a mechanism by which sleep mediates the relationship between mental health and physical illness and mortality [23], [24]. In large, representative study designs, sleep duration is often assessed via self-report, but this is unsatisfactory as these measures usually show poor agreement with objective measures [25]–[27]. However, applying the gold-standard process of polysomnography is infeasible in an epidemiological study of thousands of participants, which is needed to assess population-level associations between sleep duration and health outcomes. Some studies have asked participants to wear accelerometers for a given period of time, which primarily provides data related to physical activity but also offers the opportunity to derive a more objective measure of sleep duration [28]. A range of algorithms exist in the literature to derive an estimate of sleep duration from the raw data yielded by the accelerometer, some of which have been validated against polysomnography [29]–[31]. If the more objective measures from accelerometry were to yield similar associations to those derived from self-reported measures of sleep, this would strengthen the credibility of those associations.

Accordingly, this study aimed to robustly analyse the association at the population level between early-life mental health and sleep duration in adulthood using multiple sleep measures. To do so, we used data from the 1970 British Cohort Study (BCS70) [32]. We hypothesised that adverse mental health indicators in childhood would be associated with abnormal average sleep duration in adulthood, defined as either less than six, or more than nine, hours per night [33].

## Methods

### Sample & data characteristics

BCS70 comprises a sample of more than 17,000 residents of England, Scotland and Wales born in a single week in April 1970. Immigrants born in the reference week were also added to the sample [34]. Because of selective attrition, the sample has become less representative of the target population over time, but work on the similar National Child Development Study has indicated that leveraging the rich data available in the study in the form of auxiliary variables can restore sample representativeness and reduce bias [35].

Thus far, 11 sweeps of data collection have been conducted for BCS70. Our exposures and outcomes are derived from five sweeps as described in Table 1.

**Table 1.** Relevant sweeps of the 1970 British Cohort Study. Each sweep is primarily identified by the age at which it was conducted; this labelling age is given in bold. All other waves are detailed in Supplementary Table S1.

| **Cohort age(s)** | **Fieldwork dates** | **Survey mode** | **Sample** |
| --- | --- | --- | --- |
| **5** | 1975 | Face to face | 13,135 |
| **10–11** | 1980–1981 | Face to face | 14,875 |
| **15–16** | March–September 1986 | Face to face | 11,622 |
| **42–43** | May 2012 – April 2013 | Face to face | 9,841 |
| **46–48** | July 2016 – July 2018 | Face to face | 8,581 |

The target population of our analysis was the population alive and still residing in Great Britain at the age of 46–48 in 2016–18. Therefore, participants who had died or emigrated by the time of the age 46 sweep were excluded from analysis.

### Variables of interest

There are two key phases of the study which are relevant to this analysis. Firstly, mental health measures were sourced from the ages 5, 10 and 16 sweeps. These measures were identified with the help of the Catalogue of Mental Health Measures [36]. Where possible, these were standardised instruments, covering a range of informants. In total, we elected to use seven exposure variables, including one at age 5, two at age 10 and four at age 16. Instead of the raw Rutter Parent Questionnaire result at age 16, we chose to use the internalising and externalising behaviour measures derived and harmonised by Cohort and Longitudinal Studies Enhancement Resources (CLOSER) [37] based on that same questionnaire, to enable more straightforward comparison with other cohort studies. Table 2 details the chosen exposure variables.

**Table 2.**
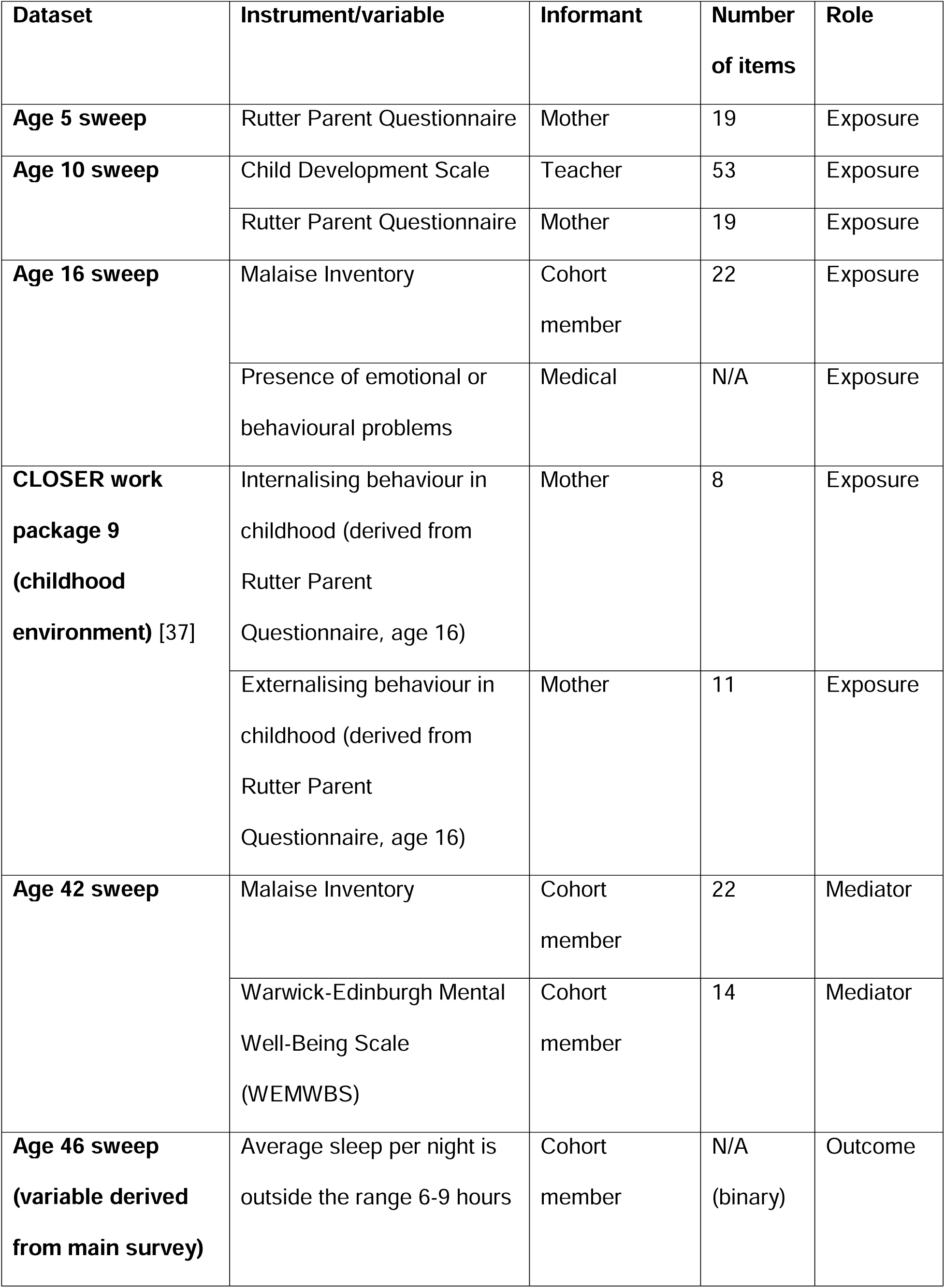
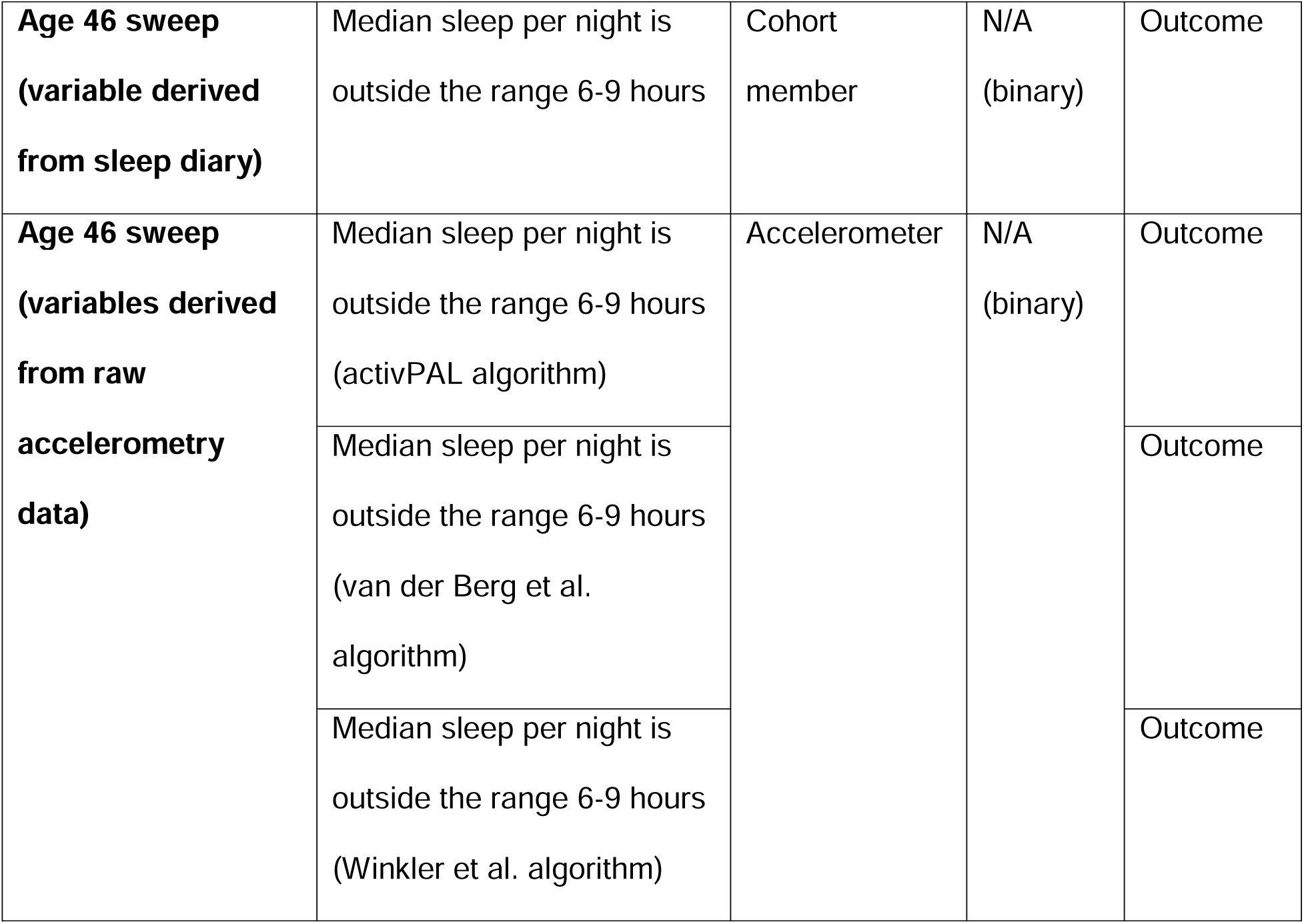
Variables of interest. In the age 16 sweep, the ‘presence of emotional or behavioural problems’ variable is a single binary item reported by the visiting clinician. For all mental health measures except the WEMWBS, which is a measure of positive mental wellbeing, a higher score indicates worse symptoms.

EARLY-LIFE MENTAL HEALTH AND SLEEP IN ADULTHOOD
| **Dataset** | **Instrument/variable** | **Informant** | **Number of items** | **Role** |
| --- | --- | --- | --- | --- |
| **Age 5 sweep** | Rutter Parent Questionnaire | Mother | 19 | Exposure |
| **Age 10 sweep** | Child Development Scale | Teacher | 53 | Exposure |
|  | Rutter Parent Questionnaire | Mother | 19 | Exposure |
| **Age 16 sweep** | Malaise Inventory | Cohort member | 22 | Exposure |
|  | Presence of emotional or behavioural problems | Medical | N/A | Exposure |
| **CLOSER work package 9 (childhood environment) [37]** | Internalising behaviour in childhood (derived from Rutter Parent Questionnaire, age 16) | Mother | 8 | Exposure |
|  | Externalising behaviour in childhood (derived from Rutter Parent Questionnaire, age 16) | Mother | 11 | Exposure |
| **Age 42 sweep** | Malaise Inventory | Cohort member | 22 | Mediator |
|  | Warwick-Edinburgh Mental Well-Being Scale (WEMWBS) | Cohort member | 14 | Mediator |
| **Age 46 sweep (variable derived from main survey)** | Average sleep per night is outside the range 6-9 hours | Cohort member | N/A (binary) | Outcome |

*Table 2.* Variables of interest.
|  |  |  |  |  |
| --- | --- | --- | --- | --- |
| **Age 46 sweep**<br>**(variable derived**<br>**from sleep diary)** | Median sleep per night is<br>outside the range 6-9 hours | Cohort<br>member | N/A<br>(binary) | Outcome |
| **Age 46 sweep**<br>**(variables derived**<br>**from raw**<br>**accelerometry**<br>**data)** | Median sleep per night is<br>outside the range 6-9 hours<br>(activPAL algorithm) | Accelerometer | N/A<br>(binary) | Outcome |
|  | Median sleep per night is<br>outside the range 6-9 hours<br>(van der Berg et al.<br>algorithm) |  |  | Outcome |
|  | Median sleep per night is<br>outside the range 6-9 hours<br>(Winkler et al. algorithm) |  |  | Outcome |

We also took into account a range of potential socioeconomic, perinatal and health-related confounders [2]. These are detailed in Table 3. Since a confounder has a causal association with both the exposure and the outcome, only covariates that refer to a time before the measurement of the exposure (and by extension, the outcome) were included.

**Table 3.**
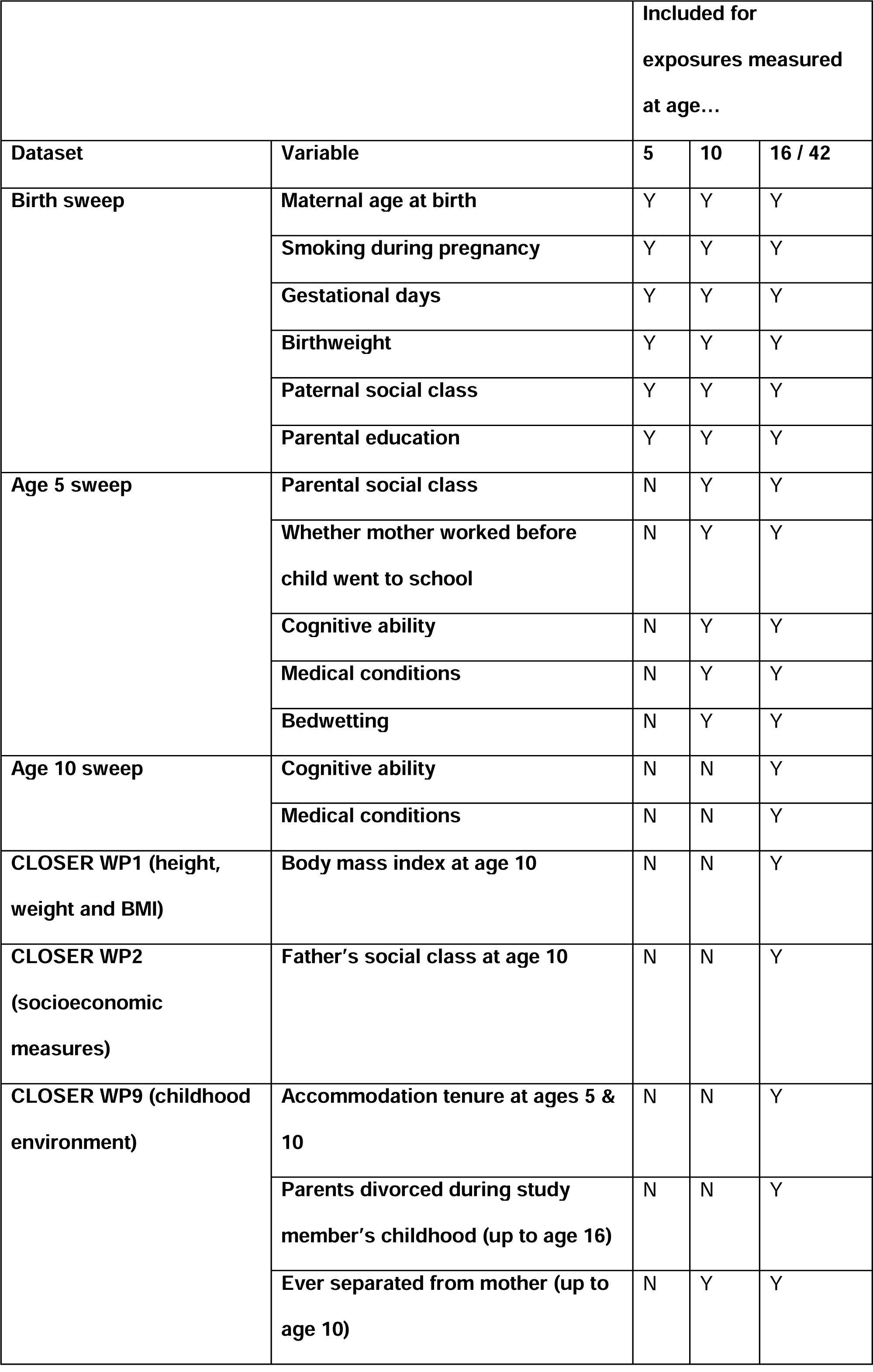
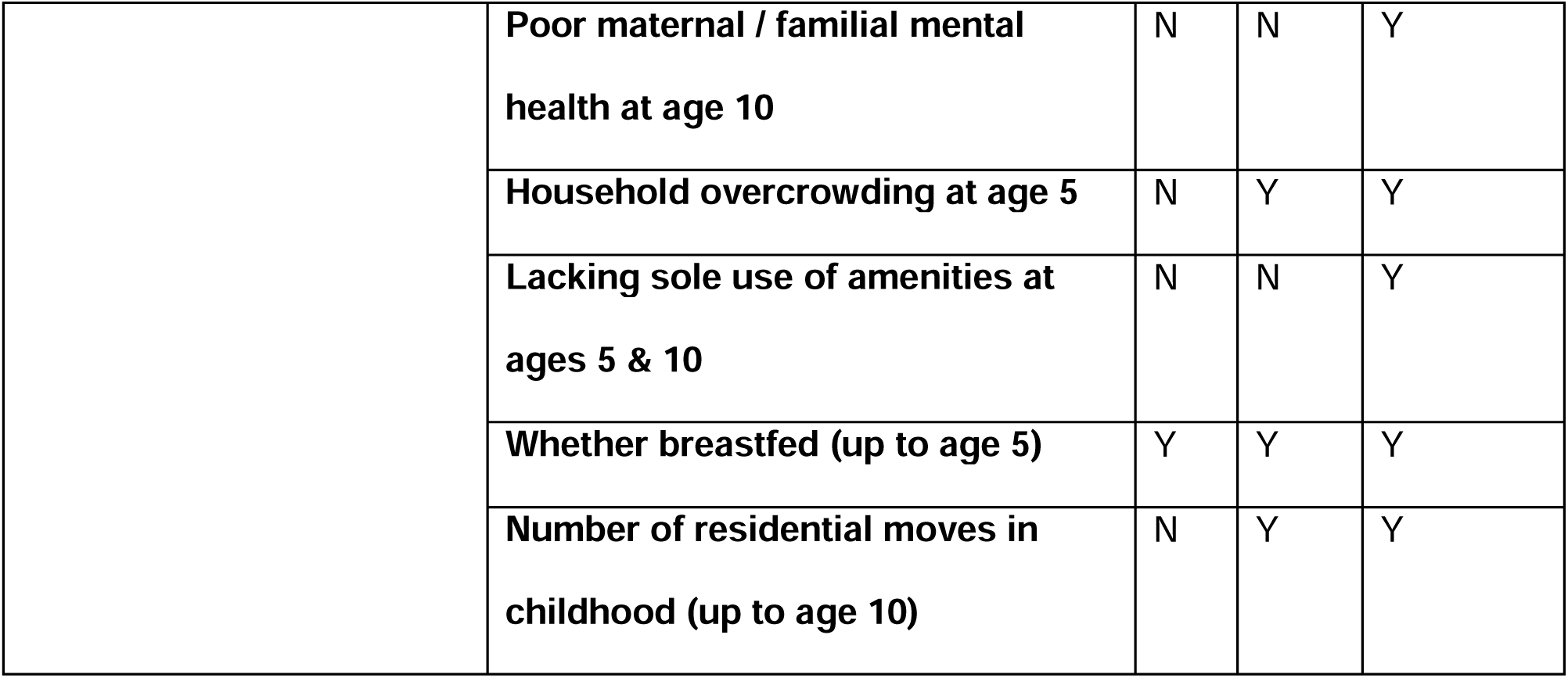
Potential socioeconomic, perinatal and health-related confounders for which models were adjusted.

|  |  | **Included for**<br>**exposures measured**<br>**at age...** |  |  |
| --- | --- | --- | --- | --- |
| **Dataset** | **Variable** | **5** | **10** | **16 / 42** |
| **Birth sweep** | **Maternal age at birth** | Y | Y | Y |
|  | **Smoking during pregnancy** | Y | Y | Y |
|  | **Gestational days** | Y | Y | Y |
|  | **Birthweight** | Y | Y | Y |
|  | **Paternal social class** | Y | Y | Y |
|  | **Parental education** | Y | Y | Y |
| **Age 5 sweep** | **Parental social class** | N | Y | Y |
|  | **Whether mother worked before child went to school** | N | Y | Y |
|  | **Cognitive ability** | N | Y | Y |
|  | **Medical conditions** | N | Y | Y |
|  | **Bedwetting** | N | Y | Y |
| **Age 10 sweep** | **Cognitive ability** | N | N | Y |
|  | **Medical conditions** | N | N | Y |
| **CLOSER WP1 (height, weight and BMI)** | **Body mass index at age 10** | N | N | Y |
| **CLOSER WP2 (socioeconomic measures)** | **Father's social class at age 10** | N | N | Y |
| **CLOSER WP9 (childhood environment)** | **Accommodation tenure at ages 5 & 10** | N | N | Y |
|  | **Parents divorced during study member's childhood (up to age 16)** | N | N | Y |
|  | **Ever separated from mother (up to age 10)** | N | Y | Y |

|  |  |  |  |  |
| --- | --- | --- | --- | --- |
|  | **Poor maternal / familial mental health at age 10** | N | N | Y |
|  | **Household overcrowding at age 5** | N | Y | Y |
|  | **Lacking sole use of amenities at ages 5 & 10** | N | N | Y |
|  | **Whether breastfed (up to age 5)** | Y | Y | Y |
|  | **Number of residential moves in childhood (up to age 10)** | N | Y | Y |

In the age 46 sweep, participants were issued with an activPAL™ 3 thigh-worn accelerometer to be worn continuously for a period of one week, and were asked to concurrently complete a sleep diary detailing the times at which they went to bed, fell asleep, woke up and got out of bed, together with a subjective rating of sleep quality and number of times awoken during the night.

Median nightly sleep duration was derived through five separate methods and each resulting measure was analysed separately. The first was simply the self-reported ‘average’ nightly sleep duration reported by cohort members in the main survey of the age 46 sweep. The second was derived from the sleep diary completed by participants during the time they were wearing the accelerometer using a bespoke algorithm (see Supplementary Data). The remaining three variables were derived from the accelerometer data according to three separate algorithms. One of these is the proprietary algorithm shipped by the accelerometer manufacturer, and the other two were sourced from literature. The available algorithms vary in complexity, and some employ high-resolution accelerometry data including the angle at which the accelerometer was positioned at any given time. To conserve computational resources, we allowed only those algorithms which use data at the level of “bouts” of sitting/lying and standing and their durations as derived by the activPAL™ device, as opposed to the raw binary data in their entirety. The two algorithms selected were published by van der Berg et al. (2016) [38] and Winkler et al. (2016) [39]. We used bespoke R code to implement these two algorithms (see Supplementary Data).

The five separate estimates of median sleep duration were manipulated to yield five separate binary indicators of abnormal sleep duration, plus five separate ternary indicators which treated abnormally short and abnormally long durations separately. These ten variables are the outcomes used in our regression analyses.

### Analysis procedure

With binary indicators of abnormal sleep duration as outcomes, we used modified Poisson regression with robust confidence intervals [40] to quantify the strength of association between early-life mental health measures and the adult sleep measures, adjusting for likely confounders. The modified Poisson approach allows the risk ratio to be directly estimated, and is straightforwardly implemented using the R packages *lmtest* [41] and *sandwich* [42]. For the ternary sleep indicators, we used multinomial logistic regression [43] via the R package *nnet* [44] to estimate relative risk ratios for abnormally short and long sleep durations compared with normal ones. The childhood mental health exposures were standardised such that the (relative) risk ratios refer to the additional risk conferred by a difference of one standard deviation in the mental health measure, in order to allow direct comparison of the associations of the various mental health variables employed in this study.

In a *post-hoc* manner, we completed two additional analysis steps. Firstly, we tested whether each childhood mental health variable in turn was associated with measures of psychological distress (Malaise Inventory score) and mental wellbeing (WEMWBS score) at age 42, controlling for the relevant confounders (see Table 3). Secondly, we included these adult mental health measures in the original analysis models. These two steps amount to an informal mediation analysis, allowing us to determine whether adult mental health might act as a mediator in the relationship between childhood mental health and sleep in adulthood.

As a sensitivity analysis check for potential residual confounding, for each regression analysis we also computed the E-value [45] to quantify the minimum strength of an association between an unmeasured confounder and both the exposure (childhood mental health) and the outcome (sleep in adulthood) that would be required to nullify the associations we identified.

To handle missing data (for proportions see Supplementary Table S2), the dataset was subjected to multiple imputation by chained equations (MICE) via the R package *mice* [46] (predictive mean matching, 180 imputations, 15 iterations). We assessed that the trace lines converged satisfactorily (see Supplementary Data). Auxiliary variables were employed based on guidance issued for the similar National Child Development Study [47] (see Supplementary Table S3), covering both predictors of non-response and likely predictors of the outcome.

All other data cleaning and analysis was conducted in R [48] version 4.2.0 and rendered in R Markdown [49] and Quarto [50] (see Supplementary Data).

The analysis procedure for this study was pre-registered after data cleaning had been completed at https://osf.io/tf6y3/. We have not deviated from that procedure.

## Results

### Outputs of accelerometry algorithms

Both the proprietary activPAL algorithm and the van der Berg et al. algorithm [38] yielded estimates of average sleep duration that tended to be significantly above both self-reported averages and diary-derived estimates (see Supplementary Table S4). By contrast, the Winkler et al. algorithm [39] yielded estimates that tended to be slightly below the self-report measures (see Supplementary Table S4). We assessed that the activPAL and van der Berg et al. algorithm-derived estimates in particular were likely to underestimate the association between early life mental health and adult sleep because a large number of normal sleep durations would be misclassified as abnormally long. The correlations among estimates of average nightly sleep duration were unexpectedly low (see Supplementary Tables S5–6). Nonetheless, we present the associations resulting from all three algorithms.

### Regression outcomes

Figure 1 shows the risk ratios of the associations between each of the seven early-life mental health measures and each of the five estimates of average sleep duration, together with the corresponding E-values [45]. All seven early-life mental health measures displayed positive associations with abnormal self-reported average sleep at age 46-48. In addition, five were associated with abnormal diary-derived estimates of average sleep and four with abnormal Winkler et al. algorithm-derived estimates. Multinomial logistic regressions indicated that these associations were primarily driven by abnormally short, rather than abnormally long, sleep durations (see Supplementary Figure S1). As might be expected given the misclassification issues described above, there were no associations between early-life mental health measures and abnormal average sleep duration as derived using the activPAL or van der Berg et al. algorithms. For the full raw model outputs, see Supplementary Tables S7–41.

**Figure 1.**
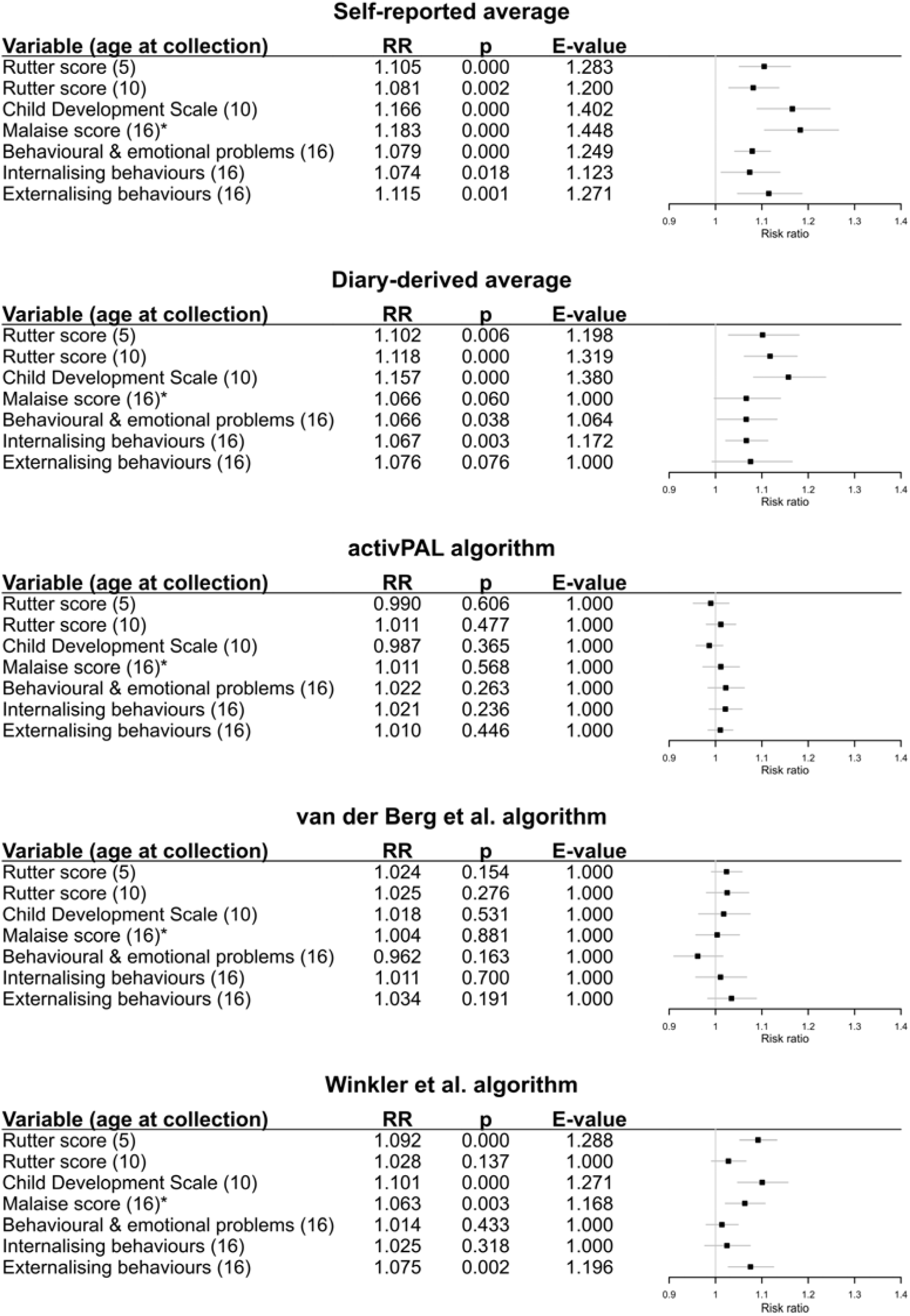
Estimated risk ratios quantifying the associations between the childhood mental health variables (standardised) and the presence of abnormal sleep duration in adulthood (derived from five separate measures of sleep). The E-value, quantifying the minimum unmeasured confounding risk ratio that would be needed to nullify each association estimated here, is also shown. Each exposure-outcome relationship was assessed in a separate model. All models were adjusted for a range of socioeconomic, perinatal and health-related covariates. Missing data were handled by multiple imputation. RR = risk ratio. * = self-reported.

### Post-hoc mediation analyses

To assess whether the associations between early-life mental health and sleep in adulthood might be explained solely by adult mental health, we performed a series of regression based informal mediation analyses. We firstly confirmed that all seven childhood mental health measures were associated with both Malaise and Warwick–Edinburgh Mental Well-Being Scale scores at age 42 (see Supplementary Table S42). Subsequently, we reproduced the regression models outlined above incorporating these two age-42 mental health measures as additional covariates. The results of these analyses are displayed in Figure 2. Note that, in models of the self-report and diary-derived sleep durations, the associations with childhood mental health are attenuated by the inclusion of these adult mental health variables. However, the point estimates of the strengths of the associations with the more objective Winkler et al. algorithm-derived sleep measures remain similar. For the full raw model outputs, see Supplementary Tables S43– 77.

**Figure 2.**
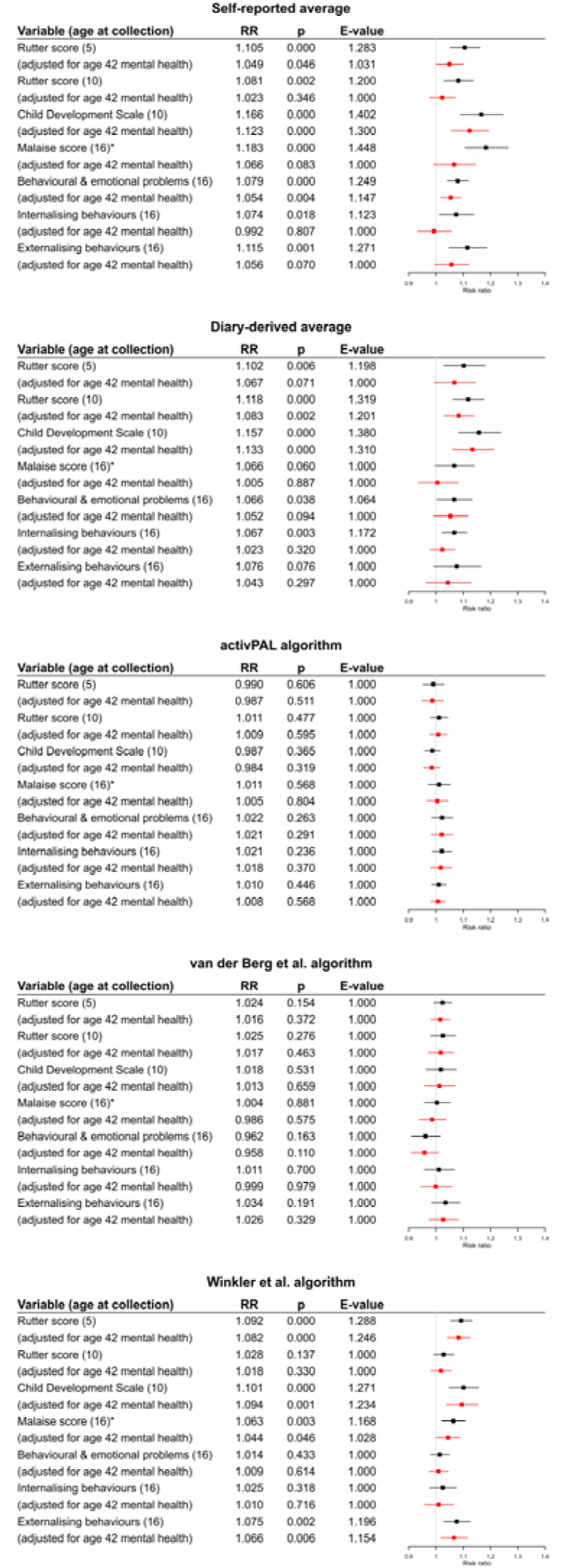
Estimated risk ratios quantifying the associations between the childhood mental health variables (standardised) and the status of sleep duration as abnormally short, normal or abnormally long in adulthood (derived from five separate measures of sleep), also including (in red) those adjusting for Malaise and Warwick*–*Edinburgh Mental Well-Being Scale scores at age 42. The E-value, quantifying the minimum unmeasured confounding risk ratio that would be needed to nullify each association estimated here, is also shown. Each exposure-outcome relationship was assessed in a separate model. All models were adjusted for a range of socioeconomic, perinatal and health-related covariates. Missing data were handled by multiple imputation. RR = risk ratio. * = self-reported.

### Results disaggregated by sex at birth

We estimated each model that we fit three times: once for the entire sample and once each for male and female participants. The models disaggregated by sex yielded very similar results to the models fit to the pooled sample. More details of this can be found in Supplementary Figures S2–7.

## Discussion

In this study we have demonstrated that there exists an association between several measures of childhood mental health symptoms and self-reported, diary-derived and accelerometry-derived estimates of nightly sleep duration at age 46. Multinomial logistic regression analyses indicated that these associations were driven by abnormally short, rather than abnormally long, sleep durations, with only a single association between a childhood mental health measure and self-reported and diary-derived abnormally long sleep duration at age 46 being found.

This study has a number of strengths, being the first population-representative study to assess the relationship between early-life mental health and sleep in adulthood. It benefits from a large sample size, with 8,581 total respondents at the age 46 sweep and 17,136 providing at least some data at some stage (with missing outcomes being imputed). The birth cohort design of BCS70 means that the study sample approximates the population born in Great Britain in 1970 and still alive and living in the UK in 2016 without the need for additional weighting procedures. The study also does not rely solely on self-reported estimates of sleep duration, which are subject to recall bias and are not necessarily reflective of participants’ true average sleep durations. We modelled the relationship between sleep duration in adulthood and a range of childhood mental health measures, collected both at various timepoints and from various informants, which allowed us to corroborate some of the associations that we did find. In addition, available in the study is a rich set of potential socioeconomic confounders, for which we could control in our models in order to reduce the impact of confounding on our estimates.

However, there are also important weaknesses that must be taken into account. Most notably, the correlations among the five measures of sleep duration that we used were low, making it difficult to assess which (if any) might be considered a reliable approximation of the true sleep duration. We also saw evidence of potential misclassification bias in some of the accelerometry algorithms. Because of practical constraints, polysomnography was not employed in the BCS70, meaning that we cannot compare against a ‘gold-standard’ measurement. In addition, our findings are applicable to those born in Great Britain in 1970 (if our assumptions about the nature of missing data hold) but cannot be generalised to other populations because of the homogeneous nature of the study sample. We have also dichotomised sleep duration into ‘normal’ and ‘abnormal’ categories (or at best summarised it as ‘abnormal’, ‘short’ or ‘long’), and so we cannot draw inferences about the size of the reduction in sleep duration associated with poor childhood mental health. Furthermore, as in any observational study, bias from residual confounding and various forms of measurement error cannot be ruled out. As in any longitudinal survey, missing data due to attrition are unavoidable. We used multiple imputation, augmenting our models with auxiliary variables in the imputation phase to maximize the plausibility of the missing-at-random assumption and restore sample representativeness, but bias due to a nonignorable missing data-generating mechanism is possible.

Previous literature had proposed mental health in adulthood may be a potential mediator of the association between childhood mental health and sleep in adulthood [4], [16]–[19]. However, our study suggests that adult mental health does not fully account for this relationship, because the associations with the more objective accelerometry-derived estimates of sleep duration persist even with the inclusion of adult mental health measures as covariates, while the associations with directly and indirectly self-reported measures of sleep duration do not to the same extent. This has two implications: firstly, mental health in adulthood is associated with self-reported nightly sleep duration and therefore with participants’ perceptions of their sleep durations. Secondly, it implies that early-life mental health is associated with sleep duration in adulthood independently of adult mental health status. This warrants further investigation, as it could signify that poor early-life mental health has long-lasting effects on sleep architecture and could therefore render sleep a plausible mediator of the deleterious relationship between early-life mental health and adult physical ill-health and mortality [23], [24]. Given socioeconomic disparities in the incidence of mental health disorders in childhood [51]–[54], it follows that long-lasting effects of early-life mental ill-health may explain part of the socioeconomic inequality in mental and physical health outcomes among adults. This would highlight the importance of prevention of mental ill-health, and equitable access to early-life mental health treatment, as public health interventions, not just for mental health but also for physical health. Alternatively, it could also suggest that developmental factors associated with poor early-life mental health are also associated with enduring abnormal sleep.

Future research should aim to confirm the associations we have found using polysomnography to measure sleep duration, and in so doing may also wish to consider the relationship between childhood mental health and sleep quality in adulthood, which we could not assess in this dataset. It should also attempt to quantify the association between poor sleep and ill-health and mortality going forward, and to assess whether sleep may indeed be a mediator of the relationship between poor childhood mental health and adult physical ill-health. It should attempt to replicate our results in other samples, including samples of adults of different ages and of different ethnic backgrounds and nationalities to those present in our sample, in order to investigate the generalisability of our findings to other populations. Finally, it should consider whether promotion of childhood mental wellbeing might lead to lasting improvements in sleep and thereby substantially improve mental and physical health in the population at large.

## Data Availability

The main survey data from BCS70 are available from the UK Data Service. The raw accelerometry data needed to derive four of the five estimates of nightly sleep duration, along with the estimates themselves, are available on application to the UCL Centre for Longitudinal Studies. All data analysis code and outputs used in this study are available at https://osf.io/a7pqd/.

https://doi.org/10.5255/UKDA-Series-200001

https://doi.org/10.5255/UKDA-Series-2000111

https://cls.ucl.ac.uk/data-access-training/data-access/accessing-data-directly-from-cls/

https://osf.io/a7pqd/

## Data sharing

The main survey data from BCS70 are available from the UK Data Service [32]. The raw accelerometry data needed to derive four of the five estimates of nightly sleep duration, along with the estimates themselves, are available on application to the UCL Centre for Longitudinal Studies. All data analysis code and outputs used in this study are available at https://osf.io/a7pqd/.

## Ethical review

Full details of the ethical review process for BCS70 are available in a report published by the UCL Centre for Longitudinal Studies [55]. A separate ethical review was not required for this study.

## Contributions

TEM (guarantor): conceptualisation, data curation, formal analysis, methodology, project administration, software, visualisation, writing – original draft, writing – review & editing GBP: conceptualisation, funding acquisition, investigation, methodology, supervision, writing – review & editing

DMA: conceptualisation, methodology, supervision, writing – review & editing

The corresponding author attests that all listed authors meet authorship criteria and that no others meeting the criteria have been omitted.

## Transparency declaration

The lead author affirms that this manuscript is an honest, accurate and transparent account of the study being reported; that no important aspects of the study have been omitted and that any discrepancies from the study as planned (and, if relevant, registered) have been explained.

## Patient and Public Involvement

Patients and the public were not involved in the research described in this article.

## Acknowledgements

The analyses in this work are based wholly on analysis of data from the 1970 British Cohort Study (BCS70). The data were deposited at the UK Data Archive by the Centre for Longitudinal Studies at the UCL Institute of Education, University of London. BCS70 is funded by the Economic and Social Research Council (ESRC). We would like to thank the study participants for their commitment to the study over the years, and the study team members for their work in curating the data.

We would also like to thank Danielle H. Bodicoat for making available the code used as the basis for our implementation of the Winkler et al. [39] algorithm.

## References

[1] N. Kawakami et al., “Early-Life Mental Disorders and Adult Household Income in the World Mental Health Surveys,” Biol. Psychiatry, vol. 72, no. 3, pp. 228–237, Aug. 2012, doi: 10.1016/j.biopsych.2012.03.009.

[2] K. Ning, P. Patalay, J. L. Maggs, and G. B. Ploubidis, “Early life mental health and problematic drinking in mid-adulthood: evidence from two British birth cohorts,” Soc. Psychiatry Psychiatr. Epidemiol., vol. 56, no. 10, pp. 1847–1858, Oct. 2021, doi: 10.1007/s00127-021-02063-3.

[3] G. B. Ploubidis, G. D. Batty, P. Patalay, D. Bann, and A. Goodman, “Association of Early-Life Mental Health With Biomarkers in Midlife and Premature Mortality: Evidence From the 1958 British Birth Cohort,” JAMA Psychiatry, vol. 78, no. 1, pp. 38–46, Jan. 2021, doi: 10.1001/jamapsychiatry.2020.2893.

[4] M. Mulraney et al., “A systematic review of the persistence of childhood mental health problems into adulthood,” Neurosci. Biobehav. Rev., vol. 129, pp. 182–205, Oct. 2021, doi: 10.1016/j.neubiorev.2021.07.030.

[5] T. Newlove-Delgado et al., “Mental Health of Children and Young People in England 2021 - wave 2 follow up to the 2017 survey,” NHS Digital. Accessed: Nov. 05, 2021. [Online]. Available: https://digital.nhs.uk/data-and-information/publications/statistical/mental-health-of-children-and-young-people-in-england/2021-follow-up-to-the-2017-survey

[6] E. McElroy, M. Tibber, P. Fearon, P. Patalay, and G. B. Ploubidis, “Socioeconomic and sex inequalities in parent-reported adolescent mental ill-health: time trends in four British birth cohorts,” J. Child Psychol. Psychiatry, vol. 64, no. 5, pp. 758–767, 2023, doi: 10.1111/jcpp.13730.

[7] F. S. Luyster, P. J. Strollo Jr., P. C. Zee, and J. K. Walsh, “Sleep: A Health Imperative,” Sleep, vol. 35, no. 6, pp. 727–734, Jun. 2012, doi: 10.5665/sleep.1846.

[8] E. S. Ford, T. J. Cunningham, and J. B. Croft, “Trends in Self-Reported Sleep Duration among US Adults from 1985 to 2012,” Sleep, vol. 38, no. 5, pp. 829–832, May 2015, doi: 10.5665/sleep.4684.

[9] K. P. Maski and S. V. Kothare, “Sleep deprivation and neurobehavioral functioning in children,” Int. J. Psychophysiol., vol. 89, no. 2, pp. 259–264, Aug. 2013, doi: 10.1016/j.ijpsycho.2013.06.019.

[10] L. Gallicchio and B. Kalesan, “Sleep duration and mortality: a systematic review and meta-analysis,” J. Sleep Res., vol. 18, no. 2, pp. 148–158, 2009, doi: 10.1111/j.1365-2869.2008.00732.x.

[11] S. R. Patel, A. Malhotra, D. J. Gottlieb, D. P. White, and F. B. Hu, “Correlates of Long Sleep Duration,” Sleep, vol. 29, no. 7, pp. 881–889, Jul. 2006, doi: 10.1093/sleep/29.7.881.

[12] M. Jike, O. Itani, N. Watanabe, D. J. Buysse, and Y. Kaneita, “Long sleep duration and health outcomes: A systematic review, meta-analysis and meta-regression,” Sleep Med. Rev., vol. 39, pp. 25–36, Jun. 2018, doi: 10.1016/j.smrv.2017.06.011.

[13] C. Baglioni et al., “Sleep and mental disorders: A meta-analysis of polysomnographic research,” Psychol. Bull., vol. 142, no. 9, pp. 969–990, 2016, doi: 10.1037/bul0000053.

[14] D. Freeman, B. Sheaves, F. Waite, A. G. Harvey, and P. J. Harrison, “Sleep disturbance and psychiatric disorders,” Lancet Psychiatry, vol. 7, no. 7, pp. 628–637, Jul. 2020, doi: 10.1016/S2215-0366(20)30136-X.

[15] P. K. Alvaro, R. M. Roberts, and J. K. Harris, “A Systematic Review Assessing Bidirectionality between Sleep Disturbances, Anxiety, and Depression,” Sleep, vol. 36, no. 7, pp. 1059–1068, Jul. 2013, doi: 10.5665/sleep.2810.

[16] A. J. Scott, T. L. Webb, M. Martyn-St James, G. Rowse, and S. Weich, “Improving sleep quality leads to better mental health: A meta-analysis of randomised controlled trials,” Sleep Med. Rev., vol. 60, p. 101556, Dec. 2021, doi: 10.1016/j.smrv.2021.101556.

[17] K. N. Anderson and A. J. Bradley, “Sleep disturbance in mental health problems and neurodegenerative disease,” Nat. Sci. Sleep, vol. 5, pp. 61–75, May 2013, doi: 10.2147/NSS.S34842.

[18] Y. Kaneita et al., “Associations between sleep disturbance and mental health status: A longitudinal study of Japanese junior high school students,” Sleep Med., vol. 10, no. 7, pp. 780–786, Aug. 2009, doi: 10.1016/j.sleep.2008.06.014.

[19] L. Yue, R. Zhao, Q. Xiao, Y. Zhuo, J. Yu, and X. Meng, “The effect of mental health on sleep quality of front-line medical staff during the COVID-19 outbreak in China: A cross-sectional study,” PLOS ONE, vol. 16, no. 6, p. e0253753, Jun. 2021, doi: 10.1371/journal.pone.0253753.

[20] I. Morales-Muñoz, M. R. Broome, and S. Marwaha, “Association of Parent-Reported Sleep Problems in Early Childhood With Psychotic and Borderline Personality Disorder Symptoms in Adolescence,” JAMA Psychiatry, vol. 77, no. 12, pp. 1256–1265, Dec. 2020, doi: 10.1001/jamapsychiatry.2020.1875.

[21] M. Lewin, J. Lopachin, J. Delorme, M. Opendak, R. M. Sullivan, and D. A. Wilson, “Early Life Trauma Has Lifelong Consequences for Sleep And Behavior,” Sci. Rep., vol. 9, no. 1, Art. no. 1, Nov. 2019, doi: 10.1038/s41598-019-53241-y.

[22] V. Schäfer and K. Bader, “Relationship Between Early-life Stress Load and Sleep in Psychiatric Outpatients: A Sleep Diary and Actigraphy Study,” Stress Health, vol. 29, no. 3, pp. 177–189, 2013, doi: 10.1002/smi.2438.

[23] L. S. Richmond-Rakerd, S. D’Souza, B. J. Milne, A. Caspi, and T. E. Moffitt, “Longitudinal Associations of Mental Disorders With Physical Diseases and Mortality Among 2.3 Million New Zealand Citizens,” *JAMA Netw*. Open, vol. 4, no. 1, p. e2033448, Jan. 2021, doi: 10.1001/jamanetworkopen.2020.33448.

[24] H. R. Colten, B. M. Altevogt, and Institute of Medicine (US) Committee on Sleep Medicine and Research, Extent and Health Consequences of Chronic Sleep Loss and Sleep Disorders. National Academies Press (US), 2006. Accessed: Dec. 01, 2022. [Online]. Available: https://www.ncbi.nlm.nih.gov/books/NBK19961/

[25] J. Girschik, L. Fritschi, J. Heyworth, and F. Waters, “Validation of Self-Reported Sleep Against Actigraphy,” J. Epidemiol., vol. 22, no. 5, pp. 462–468, 2012, doi: 10.2188/jea.JE20120012.

[26] D. S. Lauderdale, K. L. Knutson, L. L. Yan, K. Liu, and P. J. Rathouz, “Self-Reported and Measured Sleep Duration: How Similar Are They?,” Epidemiology, vol. 19, no. 6, pp. 838–845, 2008.

[27] H. N. Iskandar et al., “Self-reported sleep disturbance in Crohn’s disease is not confirmed by objective sleep measures,” Sci. Rep., vol. 10, no. 1, Art. no. 1, Feb. 2020, doi: 10.1038/s41598-020-58807-9.

[28] M. Quante, E. R. Kaplan, M. Rueschman, M. Cailler, O. M. Buxton, and S. Redline, “Practical considerations in using accelerometers to assess physical activity, sedentary behavior, and sleep,” Sleep Health, vol. 1, no. 4, pp. 275–284, Dec. 2015, doi: 10.1016/j.sleh.2015.09.002.

[29] K. M. Full et al., “Validation of a physical activity accelerometer device worn on the hip and wrist against polysomnography,” Sleep Health, vol. 4, no. 2, pp. 209–216, Apr. 2018, doi: 10.1016/j.sleh.2017.12.007.

[30] C. Smith, B. Galland, R. Taylor, and K. Meredith-Jones, “ActiGraph GT3X+ and Actical Wrist and Hip Worn Accelerometers for Sleep and Wake Indices in Young Children Using an Automated Algorithm: Validation With Polysomnography,” Front. Psychiatry, vol. 10, 2020, Accessed: Sep. 12, 2023. [Online]. Available: https://www.frontiersin.org/articles/10.3389/fpsyt.2019.00958

[31] P. J. Johansson et al., “Development and performance of a sleep estimation algorithm using a single accelerometer placed on the thigh: an evaluation against polysomnography,” J. Sleep Res., p. e13725, 2022, doi: 10.1111/jsr.13725.

[32] University College London, UCL Institute of Education, Centre for Longitudinal Studies, “1970 British Cohort Study.” UK Data Service, 2023. doi: 10.5255/UKDA-Series-200001.

[33] N. F. Watson et al., “Joint Consensus Statement of the American Academy of Sleep Medicine and Sleep Research Society on the Recommended Amount of Sleep for a Healthy Adult: Methodology and Discussion,” J. Clin. Sleep Med., vol. 11, no. 08, pp. 931–952, 2015, doi: 10.5664/jcsm.4950.

[34] A. Sullivan, M. Brown, M. Hamer, and G. B. Ploubidis, “Cohort Profile Update: The 1970 British Cohort Study (BCS70),” *Int. J. Epidemiol.*, p. dyac148, Jul. 2022, doi: 10.1093/ije/dyac148.

[35] T. Mostafa et al., “Missing at random assumption made more plausible: evidence from the 1958 British birth cohort,” J. Clin. Epidemiol., vol. 136, pp. 44–54, Aug. 2021, doi: 10.1016/j.jclinepi.2021.02.019.

[36] L. Arseneault et al., “Catalogue of Mental Health Measures.” Accessed: Sep. 12, 2023. [Online]. Available: https://www.cataloguementalhealth.ac.uk/

[37] N. Wood, M. Stafford, and D. O’Neill, “CLOSER Work Package 9: Harmonised Childhood Environment And Adult Wellbeing Measures User Guide,” 2019. [Online]. Available: https://doc.ukdataservice.ac.uk/doc/8553/mrdoc/pdf/closer_wp9_userguide_202203revision.pdf

[38] J. D. van der Berg et al., “Identifying waking time in 24-h accelerometry data in adults using an automated algorithm,” J. Sports Sci., vol. 34, no. 19, pp. 1867–1873, Oct. 2016, doi: 10.1080/02640414.2016.1140908.

[39] E. A. H. Winkler et al., “Identifying adults’ valid waking wear time by automated estimation in activPAL data collected with a 24 h wear protocol,” Physiol. Meas., vol. 37, no. 10, p. 1653, Sep. 2016, doi: 10.1088/0967-3334/37/10/1653.

[40] G. Zou, “A Modified Poisson Regression Approach to Prospective Studies with Binary Data,” Am. J. Epidemiol., vol. 159, no. 7, pp. 702–706, Apr. 2004, doi: 10.1093/aje/kwh090.

[41] T. Hothorn, A. Zeileis, R. W. Farebrother, C. Cummins, G. Millo, and D. Mitchell, “lmtest: Testing Linear Regression Models.” Mar. 21, 2022. Accessed: Dec. 02, 2022. [Online]. Available: https://CRAN.R-project.org/package=lmtest

[42] A. Zeileis, T. Lumley, N. Graham, and S. Koell, “sandwich: Robust Covariance Matrix Estimators.” Jun. 15, 2022. Accessed: Dec. 02, 2022. [Online]. Available: https://CRAN.R-project.org/package=sandwich

[43] J. Engel, “Polytomous logistic regression,” Stat. Neerlandica, vol. 42, no. 4, pp. 233–252, 1988, doi: 10.1111/j.1467-9574.1988.tb01238.x.

[44] B. Ripley and W. Venables, “nnet: Feed-Forward Neural Networks and Multinomial Log-Linear Models.” Sep. 28, 2022. Accessed: Feb. 02, 2023. [Online]. Available: https://CRAN.R-project.org/package=nnet

[45] T. J. VanderWeele and P. Ding, “Sensitivity Analysis in Observational Research: Introducing the E-Value,” Ann. Intern. Med., vol. 167, no. 4, pp. 268–274, Aug. 2017, doi: 10.7326/M16-2607.

[46] S. van Buuren and K. Groothuis-Oudshoorn, “mice: Multivariate Imputation by Chained Equations in R,” J. Stat. Softw., vol. 45, pp. 1–67, Dec. 2011, doi: 10.18637/jss.v045.i03.

[47] R. Silverwood, M. Narayanan, B. Dodgeon, and G. B. Ploubidis, “Handling missing data in the National Child Development Study,” UCL Institute of Education, 2021. Accessed: Nov. 23, 2022. [Online]. Available: https://cls.ucl.ac.uk/wp-content/uploads/2020/04/Handling-missing-data-in-the-National-Child-Development-Study-User-Guide.pdf

[48] R Core Team, R: A language and environment for statistical computing. Vienna: R Foundation for Statistical Computing, 2021. Accessed: Apr. 18, 2023. [Online]. Available: https://www.r-project.org/

[49] J. Allaire et al., “rmarkdown: Dynamic Documents for R.” Mar. 10, 2022. Accessed: Mar. 30, 2022. [Online]. Available: https://CRAN.R-project.org/package=rmarkdown

[50] J. J. Allaire, C. Teague, C. Scheidegger, Y. Xie, and C. Dervieux, “Quarto.” Jan. 2022. doi: 10.5281/zenodo.5960048.

[51] M. Vaalavuo, R. Niemi, and J. Suvisaari, “Growing up unequal? Socioeconomic disparities in mental disorders throughout childhood in Finland,” *SSM - Popul*. Health, vol. 20, p. 101277, Dec. 2022, doi: 10.1016/j.ssmph.2022.101277.

[52] E. Davis, M. G. Sawyer, S. K. Lo, N. Priest, and M. Wake, “Socioeconomic Risk Factors for Mental Health Problems in 4–5-Year-Old Children: Australian Population Study,” Acad. Pediatr., vol. 10, no. 1, pp. 41–47, Jan. 2010, doi: 10.1016/j.acap.2009.08.007.

[53] T. Bøe, S. Øverland, A. J. Lundervold, and M. Hysing, “Socioeconomic status and children’s mental health: results from the Bergen Child Study,” Soc. Psychiatry Psychiatr. Epidemiol., vol. 47, no. 10, pp. 1557–1566, Oct. 2012, doi: 10.1007/s00127-011-0462-9.

[54] M. J. Essex et al., “Exploring Risk Factors for the Emergence of Children’s Mental Health Problems,” Arch. Gen. Psychiatry, vol. 63, no. 11, pp. 1246–1256, Nov. 2006, doi: 10.1001/archpsyc.63.11.1246.

[55] P. Shepherd and E. Gilbert, “1970 British Cohort Study: Ethical Review and Consent,” UCL Centre for Longitudinal Studies, 2, 2019. Accessed: Sep. 19, 2023. [Online]. Available: https://cls.ucl.ac.uk/wp-content/uploads/2017/07/BCS70-Ethical-review-and-Consent-2019.pdf

