## Supplementary Information for "Associations between early-life mental health and abnormal sleep duration in midlife: findings from a prospective cohort study in Great Britain"

### Dataset information: Tables S1–3

| Cohort age(s) | Fieldwork dates | Survey mode | Sample |
|---|---|---|---|
| **Birth** | April 1970 | Face to face | 17 196 |
| **5** | 1975 | Face to face | 13 135 |
| **10**–11 | 1980–1981 | Face to face | 14 875 |
| 15–**16** | March–September 1986 | Face to face | 11 622 |
| **26** | April–September 1996 | Postal | 9 003 |
| 29–**30** | November 1999 – September 2000 | Face to face | 11 261 |
| **34**–35 | February 2004 – June 2005 | Face to face | 9 665 |
| **38**–39 | October 2008 – May 2009 | Telephone | 8 874 |
| **42**–43 | May 2012 – April 2013 | Face to face | 9 841 |
| **46**–48 | July 2016 – July 2018 | Face to face | 8 581 |
| **51**–53 | Summer 2021 – January 2024 | Face to face / video link | TBD |

*Table S1.* All sweeps of the 1970 British Cohort Study, 1970–2023. Each sweep is primarily identified by the age at which it was conducted; this labelling age is given in bold.

| Variable | ID in dataset | Missingness (%) | Variable | ID in dataset | Missingness (%) |
|---|---|---|---|---|---|
| Participant ID (not imputed) | bcsid | 0.0 | Residential moves in childhood | resmove | 24.8 |
| Maternal age at birth | a0005a | 8.8 | Separated from mother in childhood | sepmumbcs | 21.5 |
| Father's social class (1970) | a0014 | 15.9 | Housing tenure (childhood) | tenure | 35.3 |
| Smoking during pregnancy | a0043b | 8.8 | BMI (10) | bmi | 35.9 |
| Gestational age | a0195a | 12.9 | Father's social class (10) | fclrg90 | 24.5 |
| Genital tract bleeding | a0230 | 8.9 | Median sleep rating (46) | md_sleeprate | 66.4 |
| Birth sex | a0255 | 8.3 | CDS (10) | dv_cds_10 | 50.5 |
| Birthweight | a0278 | 8.4 | Father's education | dv_father_schl | 71.2 |
| Bedwetting | d016a | 25.5 | Medical conditions (5) | dv_med_5 | 29.7 |
| Rutter (5) | d119 | 25.6 | Medical conditions (10) | dv_med_10 | 25.6 |
| Mother's job (5) | e216a | 35.5 | Income source (16) | dv_incsource | 46.1 |
| Cognitive ability (5) | dv_cog_abil_5 | 76.2 | Positive activities | dv_pos_act | 25.8 |
| School attendance | j111 | 27.5 | Alcohol (16) | dv_alcohol | 65.6 |
| Cognitive ability (10) | dv_bas_g | 33.3 | Accidents (26) | dv_accidents | 51.4 |
| Rutter (10) | BD3MRUTT | 27.1 | Hospital admissions (30) | dv_hospital | 35.1 |
| Behavioural / emotional problems (16) | rd6m_1 | 66.3 | Work-related training | dv_training | 35.1 |
| Arithmetic test (16) | mathscore | 78.8 | BCS participation | dv_participation | 0.0 |
| Malaise (16) | BD4MAL | 68.6 | Organisation membership | dv_organisations | 50.2 |
| School type | BDSTYPE | 28.9 | Marital status (42) | dv_marital | 43.1 |
| People for advice / support | emosup | 35.0 | Parental education (birth) | dv_par_edu_birth | 9.1 |
| Voted in 1997 GE | vote97 | 35.2 | Social class (5) | dv_sc_age_5 | 68.7 |
| Computer ownership | B9SCQ17 | 49.2 | Social class (16) | dv_sc_age_16 | 63.4 |
| No. cars / vans owned | B9SCQ19 | 49.1 | Self-reported sleep (binary) | dv_sr_sleep_abn | 49.7 |
| Housing tenure (42) | B9TEN | 42.8 | Sleep diary sleep (binary) | dv_sd_sleep_abn | 66.7 |
| Malaise (42) | BD9MAL | 49.7 | activPAL algorithm sleep (binary) | dv_pal_sleep_abn | 68.4 |
| WEMWBS (42) | BD9WEMWB | 52.7 | van der Berg et al. algorithm sleep (binary) | dv_vdb_sleep_abn | 70.5 |
| Sole use of amenities | ameni | 23.7 | Winkler et al. algorithm sleep (binary) | dv_winkler_sleep_abn | 69.2 |
| Breastfeeding | brfed | 25.8 | Self-reported sleep (binary) | dv_sr_sleep_abn_cat | 49.7 |
| Overcrowding | crowd | 26.1 | Sleep diary sleep (ternary) | dv_sd_sleep_abn_cat | 66.7 |
| Parental divorce | divorce | 14.3 | activPAL algorithm sleep (ternary) | dv_pal_sleep_abn_cat | 68.4 |
| Externalising behaviour | extbcsz | 53.4 | van der Berg et al. algorithm sleep (ternary) | dv_vdb_sleep_abn_cat | 70.5 |
| Internalising behaviour | intbcsz | 53.2 | Winkler et al. algorithm sleep (ternary) | dv_winkler_sleep_abn_cat | 69.2 |
| Poor familial mental health | prmnh | 27.8 | | **TOTAL** | **38.4** |

*Table S2.* Summary of the percentage of missing data in each variable in the study datasets.

| Dataset | Variable | Variable ID(s) | Recoding |
|---|---|---|---|
| **Birth sweep** | Occurrence of genital tract bleeding | A0230 | |
| **Age 10 sweep** | Total days missed schooling | j111 | |
| | Positive activities outside school | m84-m101 | Summation |
| **Age 16 sweep** | Alcohol consumption in past 7 days | hd5.1-hd5.8 | Binary: if any alcohol consumption in the last 7 days ‘1’, else ‘0’ |
| | Sources of income | oe1.1-oe1.19 | Binary: whether or not there is income from employment or investments |
| | Arithmetic test score | mathscore | |
| | School type | BDSTYPE | |
| **Age 26 sweep** | Accidents and assaults | b960539; b960540 | Combined into one variable describing the number of incidents (including 0) |
| **Age 30 sweep** | Apprenticeships and work-related training | aptrain; wrktrain | Combined such that if either is “yes”, the result is ’1’, else ‘0’ |
| | Availability of people for advice/support | emosup | |
| | Number of hospital admissions | hospital; numadmn | Combined into one variable describing the number of admissions (including 0) |
| | Whether voted in 1997 General Election | vote97 | |
| **Age 42 sweep** | Marital status | B9HMS; B9MARCHK; B9DIVCHK | Combined into one variable describing marital status as “married”, “divorced/separated/widowed” or “single” |
| | Organisational membership | B9SCQ8A-B9SCQ8P | Binary: if participant is a member of any organisation, the result is ‘1’, else ‘0’ |
| | Computer ownership | B9SCQ17 | |
| | Car/van ownership | B9SCQ19 | Binary: if any are owned ‘1’, else ‘0’ |
| | Housing tenure | B9TEN | Grouping into “owned” vs “not owned” |
| **Age 46 sweep** | Energy/fatigue score | BD10ENFA | |
| | Malaise Inventory | BD10MAL | |
| | Body mass index (nurse-measured) | BD10MBMI | |
| | Seen doctor/specialist for mental health problems | B10MHPRB1-B10MHPRB6 | Binary: if any are true ‘1’, else ‘0’ |
| | Warwick Edinburgh Mental Well-Being Scale | B10WEMWB | |
| **Age 46 sweep (sleep diary)** | Average self-reported sleep quality | sleeprate1-sleeprate8 | Average rating, ignoring missing values |
| **Response dataset** | Response outcomes by wave | OUTCME01-OUTCME10 | Binary: if participant was productive at all sweeps ‘1’, else ‘0’ |

*Table S3.* Auxiliary variables included in the study datasets to render the “missing at random” assumption more credible and thereby improve the validity of multiple imputation. Selected variables were recoded as specified to aid convergence.

### Sleep measure summaries: Tables S4–6

| | Binary classifications (%) | | Ternary classifications (%) | | |
|---|---|---|---|---|---|
| | Normal | Abnormal | Short | Normal | Long |
| **Self-report** | 84.8 | 15.2 | 14.1 | 84.9 | 1.1 |
| **Sleep diary** | 85.3 | 14.7 | 11.1 | 85.7 | 3.2 |
| **activPAL** | 62.0 | 38.0 | 8.3 | 61.9 | 29.9 |
| **van der Berg et al.** | 66.6 | 33.4 | 3.3 | 66.9 | 29.8 |
| **Winkler et al.** | 65.1 | 34.9 | 30.4 | 65.5 | 4.1 |

*Table S4.* Summaries of percentages of participants allocated to each category of sleep in each measure. The slight differences in percentages classified as "normal" cf. "abnormal" between binary and ternary measures is because these were imputed in separate datasets.

| | **Self-report** | **Sleep diary** | **activPAL algorithm** | **van der Berg et al. algorithm** | **Winkler et al. algorithm** |
|---|---|---|---|---|---|
| **Self-report** | 1.00 | | | | |
| **Sleep diary** | 0.52 | 1.00 | | | |
| **activPAL algorithm** | 0.07 | 0.09 | 1.00 | | |
| **van der Berg et al. algorithm** | 0.06 | 0.10 | 0.36 | 1.00 | |
| **Winkler et al. algorithm** | 0.25 | 0.28 | 0.19 | 0.13 | 1.00 |

*Table S5.* Tetrachoric correlation coefficients among the binary measures of abnormal sleep, where a value of 0 indicates an average sleep duration between 6 and 9 hours, and a value of 1 indicates an average sleep duration outside that range. Values are pooled from the imputed datasets.

| | Self-report | Sleep diary | activPAL algorithm | van der Berg et al. algorithm | Winkler et al. algorithm |
|---|---|---|---|---|---|
| Self-report | 1.00 | | | | |
| Sleep diary | 0.52 | 1.00 | | | |
| activPAL algorithm | 0.10 | 0.15 | 1.00 | | |
| van der Berg et al. algorithm | 0.08 | 0.14 | 0.23 | 1.00 | |
| Winkler et al. algorithm | 0.23 | 0.25 | 0.24 | 0.14 | 1.00 |

*Table S6.* Polychoric correlation coefficients among the ternary measures of abnormal sleep duration, where the average sleep duration is classified as “short” (< 6 hours), “normal” (between 6 and 9 hours) or “long” (> 9 hours). Values are pooled from the imputed datasets.

### Multinomial regressions: Figure S1

#### Abnormally short sleep duration

**Self-reported average**

| Variable (age at collection) | RRR | p | E-value |
|---|---|---|---|
| Rutter score (5) | 1.142 | 0.000 | 1.332 |
| Rutter score (10) | 1.105 | 0.003 | 1.223 |
| Child Development Scale (10) | 1.212 | 0.000 | 1.462 |
| Malaise score (16)* | 1.241 | 0.000 | 1.541 |
| Behavioural & emotional problems (16) | 1.090 | 0.005 | 1.192 |
| Internalising behaviours (16) | 1.077 | 0.059 | 1.000 |
| Externalising behaviours (16) | 1.138 | 0.000 | 1.341 |

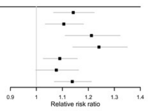

**Diary-derived average**

| Variable (age at collection) | RRR | p | E-value |
|---|---|---|---|
| Rutter score (5) | 1.078 | 0.059 | 1.000 |
| Rutter score (10) | 1.099 | 0.031 | 1.106 |
| Child Development Scale (10) | 1.182 | 0.003 | 1.308 |
| Malaise score (16)* | 1.059 | 0.368 | 1.000 |
| Behavioural & emotional problems (16) | 1.101 | 0.106 | 1.000 |
| Internalising behaviours (16) | 1.041 | 0.464 | 1.000 |
| Externalising behaviours (16) | 1.059 | 0.195 | 1.000 |

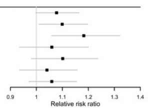

**activPAL algorithm**

| Variable (age at collection) | RRR | p | E-value |
|---|---|---|---|
| Rutter score (5) | 0.989 | 0.835 | 1.000 |
| Rutter score (10) | 1.013 | 0.830 | 1.000 |
| Child Development Scale (10) | 0.996 | 0.956 | 1.000 |
| Malaise score (16)* | 1.045 | 0.342 | 1.000 |
| Behavioural & emotional problems (16) | 1.092 | 0.101 | 1.000 |
| Internalising behaviours (16) | 1.026 | 0.568 | 1.000 |
| Externalising behaviours (16) | 1.023 | 0.637 | 1.000 |

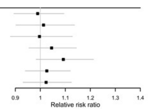

**van der Berg et al. algorithm**

| Variable (age at collection) | RRR | p | E-value |
|---|---|---|---|
| Rutter score (5) | 1.051 | 0.409 | 1.000 |
| Rutter score (10) | 1.091 | 0.188 | 1.000 |
| Child Development Scale (10) | 1.080 | 0.374 | 1.000 |
| Malaise score (16)* | 1.041 | 0.529 | 1.000 |
| Behavioural & emotional problems (16) | 1.008 | 0.914 | 1.000 |
| Internalising behaviours (16) | 1.068 | 0.391 | 1.000 |
| Externalising behaviours (16) | 1.086 | 0.349 | 1.000 |

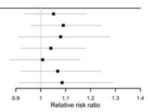

**Winkler et al. algorithm**

| Variable (age at collection) | RRR | p | E-value |
|---|---|---|---|
| Rutter score (5) | 1.136 | 0.007 | 1.232 |
| Rutter score (10) | 1.040 | 0.095 | 1.000 |
| Child Development Scale (10) | 1.178 | 0.000 | 1.362 |
| Malaise score (16)* | 1.081 | 0.011 | 1.153 |
| Behavioural & emotional problems (16) | 1.013 | 0.688 | 1.000 |
| Internalising behaviours (16) | 1.038 | 0.409 | 1.000 |
| Externalising behaviours (16) | 1.118 | 0.003 | 1.239 |

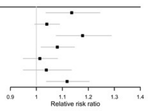

#### Abnormally long sleep duration

**Self-reported average**

| Variable (age at collection) | RRR | p | E-value |
|---|---|---|---|
| Rutter score (5) | 1.098 | 0.328 | 1.000 |
| Rutter score (10) | 1.003 | 0.976 | 1.000 |
| Child Development Scale (10) | 1.091 | 0.492 | 1.000 |
| Malaise score (16)* | 1.103 | 0.510 | 1.000 |
| Behavioural & emotional problems (16) | 1.107 | 0.171 | 1.000 |
| Internalising behaviours (16) | 1.184 | 0.210 | 1.000 |
| Externalising behaviours (16) | 1.213 | 0.033 | 1.145 |

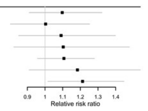

**Diary-derived average**

| Variable (age at collection) | RRR | p | E-value |
|---|---|---|---|
| Rutter score (5) | 1.074 | 0.273 | 1.000 |
| Rutter score (10) | 1.072 | 0.255 | 1.000 |
| Child Development Scale (10) | 1.134 | 0.044 | 1.064 |
| Malaise score (16)* | 1.060 | 0.603 | 1.000 |
| Behavioural & emotional problems (16) | 1.015 | 0.847 | 1.000 |
| Internalising behaviours (16) | 0.997 | 0.958 | 1.000 |
| Externalising behaviours (16) | 1.005 | 0.932 | 1.000 |

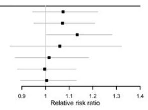

**activPAL algorithm**

| Variable (age at collection) | RRR | p | E-value |
|---|---|---|---|
| Rutter score (5) | 0.986 | 0.613 | 1.000 |
| Rutter score (10) | 1.007 | 0.844 | 1.000 |
| Child Development Scale (10) | 0.985 | 0.607 | 1.000 |
| Malaise score (16)* | 1.027 | 0.455 | 1.000 |
| Behavioural & emotional problems (16) | 1.043 | 0.221 | 1.000 |
| Internalising behaviours (16) | 1.014 | 0.609 | 1.000 |
| Externalising behaviours (16) | 1.000 | 0.990 | 1.000 |

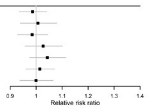

**van der Berg et al. algorithm**

| Variable (age at collection) | RRR | p | E-value |
|---|---|---|---|
| Rutter score (5) | 1.033 | 0.241 | 1.000 |
| Rutter score (10) | 1.047 | 0.181 | 1.000 |
| Child Development Scale (10) | 1.025 | 0.621 | 1.000 |
| Malaise score (16)* | 1.010 | 0.827 | 1.000 |
| Behavioural & emotional problems (16) | 0.993 | 0.889 | 1.000 |
| Internalising behaviours (16) | 1.050 | 0.227 | 1.000 |
| Externalising behaviours (16) | 1.051 | 0.236 | 1.000 |

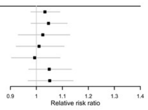

**Winkler et al. algorithm**

| Variable (age at collection) | RRR | p | E-value |
|---|---|---|---|
| Rutter score (5) | 1.017 | 0.730 | 1.000 |
| Rutter score (10) | 0.996 | 0.941 | 1.000 |
| Child Development Scale (10) | 0.990 | 0.888 | 1.000 |
| Malaise score (16)* | 1.050 | 0.311 | 1.000 |
| Behavioural & emotional problems (16) | 0.978 | 0.690 | 1.000 |
| Internalising behaviours (16) | 0.997 | 0.940 | 1.000 |
| Externalising behaviours (16) | 1.005 | 0.922 | 1.000 |

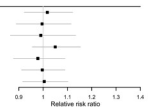

*Figure S1.* Estimated relative risk ratios quantifying the associations between the childhood mental health variables (standardised) and the presence of abnormally short, and abnormally long, sleep duration in adulthood (derived from five separate measures of sleep). The E-value, quantifying the minimum unmeasured confounding risk ratio that would be needed to nullify each association estimated here, is also shown. Each exposure-outcome relationship was assessed in a separate model. All models were adjusted for a range of socioeconomic, perinatal and health-related covariates. Missing data were handled by multiple imputation. RRR = relative risk ratio. * = self-reported.

### Raw Poisson regression outputs (initial): Tables S7–41

The following pages contain raw model outputs for the initial modified Poisson regressions used to produce main text Figure 1. The tables are as follows:

| **Sleep measure ⇒**<br><br>**Mental health measure ⇓** | **Self-report** | **Sleep diary** | **activPAL algorithm** | **van der Berg et al. algorithm** | **Winkler et al. algorithm** |
|---|---|---|---|---|---|
| **Rutter score (age 5)** | Table S7 | Table S8 | Table S9 | Table S10 | Table S11 |
| **Rutter score (age 10)** | Table S12 | Table S13 | Table S14 | Table S15 | Table S16 |
| **Child Development Scale (age 10)** | Table S17 | Table S18 | Table S19 | Table S20 | Table S21 |
| **Malaise Inventory (age 16)** | Table S22 | Table S23 | Table S24 | Table S25 | Table S26 |
| **Behavioural & emotional problems (age 16)** | Table S27 | Table S28 | Table S29 | Table S30 | Table S31 |
| **Internalising behaviour (age 16)** | Table S32 | Table S33 | Table S34 | Table S35 | Table S36 |
| **Externalising behaviour (age 16)** | Table S37 | Table S38 | Table S39 | Table S40 | Table S41 |

| Term | Estimate | Standard error | Statistic | Degrees of freedom | p-value |
|---|---|---|---|---|---|
| **(Intercept)** | -1.03 | 0.41 | -2.51 | 2502.77 | 0.01 |
| **d119** | 0.10 | 0.03 | 3.92 | 3072.16 | 0.00 |
| **a0005a** | -0.38 | 0.15 | -2.56 | 8857.21 | 0.01 |
| **a0043b.L** | 0.26 | 0.07 | 3.91 | 2792.54 | 0.00 |
| **a0043b.Q** | 0.12 | 0.09 | 1.31 | 2888.61 | 0.19 |
| **a0043b.C** | 0.05 | 0.08 | 0.67 | 3363.75 | 0.51 |
| **a0043b^4** | -0.04 | 0.10 | -0.42 | 2224.17 | 0.68 |
| **a0043b^5** | -0.06 | 0.12 | -0.49 | 2658.81 | 0.62 |
| **a0195a** | -0.10 | 0.53 | -0.19 | 4210.16 | 0.85 |
| **a0278** | -0.84 | 0.24 | -3.55 | 10466.60 | 0.00 |
| **a0014.L** | 0.54 | 0.08 | 6.93 | 4775.40 | 0.00 |
| **a0014.Q** | 0.03 | 0.06 | 0.53 | 6047.06 | 0.60 |
| **a0014.C** | 0.07 | 0.08 | 0.90 | 3372.73 | 0.37 |
| **a0014^4** | 0.09 | 0.06 | 1.46 | 2966.67 | 0.14 |
| **dv_par_edu_birth** | -2.20 | 1.60 | -1.37 | 2640.51 | 0.17 |
| **brfed.L** | 0.12 | 0.05 | 2.25 | 3601.43 | 0.02 |
| **brfed.Q** | -0.03 | 0.06 | -0.56 | 3634.88 | 0.58 |

*Table S7.* Raw output from modified Poisson regression (with log link) of Rutter score at age 5 (d119) against self-reported average sleep duration (binarised to normal vs abnormal) at age 46, adjusted for potential confounders (see main text Table 3). The parameter estimates in the "Estimate" column and the associated standard errors have not been exponentiated.

| Term | Estimate | Standard error | Statistic | Degrees of freedom | p-value |
|---|---|---|---|---|---|
| **(Intercept)** | -1.86 | 0.46 | -4.08 | 1971.84 | 0.00 |
| **d119** | 0.10 | 0.04 | 2.74 | 1770.63 | 0.01 |
| **a0005a** | 0.35 | 0.22 | 1.60 | 2419.27 | 0.11 |
| **a0043b.L** | 0.23 | 0.07 | 3.13 | 2414.36 | 0.00 |
| **a0043b.Q** | 0.20 | 0.10 | 2.00 | 2542.72 | 0.05 |
| **a0043b.C** | 0.06 | 0.09 | 0.69 | 2449.99 | 0.49 |
| **a0043b^4** | -0.12 | 0.09 | -1.23 | 3008.41 | 0.22 |
| **a0043b^5** | -0.06 | 0.12 | -0.50 | 2904.42 | 0.61 |
| **a0195a** | -0.07 | 0.63 | -0.10 | 3155.99 | 0.92 |
| **a0278** | -0.13 | 0.29 | -0.45 | 4735.12 | 0.65 |
| **a0014.L** | 0.56 | 0.10 | 5.70 | 2363.54 | 0.00 |
| **a0014.Q** | 0.10 | 0.08 | 1.28 | 2592.25 | 0.20 |
| **a0014.C** | 0.14 | 0.09 | 1.57 | 2610.94 | 0.12 |
| **a0014^4** | 0.12 | 0.05 | 2.29 | 5356.55 | 0.02 |
| **dv_par_edu_birth** | -0.73 | 1.35 | -0.54 | 2334.04 | 0.59 |
| **brfed.L** | 0.05 | 0.07 | 0.73 | 2102.69 | 0.47 |
| **brfed.Q** | 0.01 | 0.10 | 0.05 | 1482.08 | 0.96 |

*Table S8.* Raw output from modified Poisson regression (with log link) of Rutter score at age 5 (d119) against median sleep duration (binarised to normal vs abnormal) derived from the sleep diary at age 46, adjusted for potential confounders (see main text Table 3). The parameter estimates in the “Estimate” column and the associated standard errors have not been exponentiated.

| Term | Estimate | Standard error | Statistic | Degrees of freedom | p-value |
|---|---|---|---|---|---|
| **(Intercept)** | -0.48 | 0.21 | -2.27 | 2238.75 | 0.02 |
| **d119** | -0.01 | 0.02 | -0.52 | 1841.01 | 0.61 |
| **a0005a** | -0.32 | 0.10 | -3.25 | 4072.13 | 0.00 |
| **a0043b.L** | -0.03 | 0.05 | -0.54 | 1825.10 | 0.59 |
| **a0043b.Q** | -0.01 | 0.06 | -0.26 | 2171.98 | 0.79 |
| **a0043b.C** | 0.01 | 0.06 | 0.11 | 1638.93 | 0.91 |
| **a0043b^4** | -0.06 | 0.04 | -1.31 | 3719.87 | 0.19 |
| **a0043b^5** | -0.02 | 0.07 | -0.25 | 2257.65 | 0.80 |
| **a0195a** | -0.41 | 0.47 | -0.87 | 1691.12 | 0.38 |
| **a0278** | -0.23 | 0.22 | -1.07 | 2016.68 | 0.28 |
| **a0014.L** | 0.07 | 0.06 | 1.23 | 1980.31 | 0.22 |
| **a0014.Q** | 0.04 | 0.05 | 0.72 | 1877.85 | 0.47 |
| **a0014.C** | 0.00 | 0.05 | 0.07 | 2507.33 | 0.94 |
| **a0014^4** | -0.01 | 0.04 | -0.15 | 1834.72 | 0.88 |
| **dv_par_edu_birth** | -0.51 | 0.50 | -1.03 | 2706.73 | 0.30 |
| **brfed.L** | 0.04 | 0.03 | 1.10 | 2173.50 | 0.27 |
| **brfed.Q** | -0.01 | 0.03 | -0.43 | 3351.97 | 0.67 |

*Table S9.* Raw output from modified Poisson regression (with log link) of Rutter score at age 5 (d119) against median sleep duration (binarised to normal vs abnormal) derived from the activPAL algorithm at age 46, adjusted for potential confounders (see main text Table 3). The parameter estimates in the “Estimate” column and the associated standard errors have not been exponentiated.

| Term | Estimate | Standard error | Statistic | Degrees of freedom | p-value |
|---|---|---|---|---|---|
| **(Intercept)** | -1.01 | 0.26 | -3.90 | 1926.57 | 0.00 |
| **d119** | 0.02 | 0.02 | 1.43 | 3165.09 | 0.15 |
| **a0005a** | -0.10 | 0.15 | -0.66 | 1860.44 | 0.51 |
| **a0043b.L** | 0.03 | 0.06 | 0.44 | 1666.25 | 0.66 |
| **a0043b.Q** | -0.07 | 0.07 | -0.96 | 1723.34 | 0.33 |
| **a0043b.C** | -0.17 | 0.07 | -2.61 | 1672.56 | 0.01 |
| **a0043b^4** | 0.10 | 0.06 | 1.57 | 2015.29 | 0.12 |
| **a0043b^5** | 0.03 | 0.08 | 0.42 | 2116.11 | 0.67 |
| **a0195a** | -0.26 | 0.45 | -0.57 | 2173.57 | 0.57 |
| **a0278** | 0.24 | 0.18 | 1.34 | 3794.35 | 0.18 |
| **a0014.L** | 0.23 | 0.09 | 2.50 | 1190.40 | 0.01 |
| **a0014.Q** | 0.06 | 0.08 | 0.77 | 1195.15 | 0.44 |
| **a0014.C** | -0.04 | 0.05 | -0.89 | 2827.30 | 0.38 |
| **a0014^4** | 0.01 | 0.04 | 0.30 | 2702.75 | 0.76 |
| **dv_par_edu_birth** | -0.36 | 0.43 | -0.83 | 4367.81 | 0.40 |
| **brfed.L** | 0.03 | 0.04 | 0.60 | 1647.43 | 0.55 |
| **brfed.Q** | 0.05 | 0.05 | 1.04 | 2278.21 | 0.30 |

*Table S10.* Raw output from modified Poisson regression (with log link) of Rutter score at age 5 (d119) against median sleep duration (binarised to normal vs abnormal) derived from the van der Berg et al. algorithm at age 46, adjusted for potential confounders (see main text Table 3). The parameter estimates in the “Estimate” column and the associated standard errors have not been exponentiated.

| Term | Estimate | Standard error | Statistic | Degrees of freedom | p-value |
|---|---|---|---|---|---|
| **(Intercept)** | -1.07 | 0.15 | -7.24 | 7106.07 | 0.00 |
| **d119** | 0.09 | 0.02 | 4.71 | 2050.82 | 0.00 |
| **a0005a** | 0.06 | 0.18 | 0.31 | 1334.32 | 0.76 |
| **a0043b.L** | -0.02 | 0.04 | -0.48 | 2832.88 | 0.63 |
| **a0043b.Q** | 0.02 | 0.06 | 0.37 | 2228.28 | 0.71 |
| **a0043b.C** | 0.07 | 0.04 | 1.78 | 4944.71 | 0.08 |
| **a0043b^4** | 0.03 | 0.05 | 0.51 | 2682.87 | 0.61 |
| **a0043b^5** | 0.04 | 0.07 | 0.60 | 2458.84 | 0.55 |
| **a0195a** | -0.06 | 0.34 | -0.19 | 3882.10 | 0.85 |
| **a0278** | -0.11 | 0.27 | -0.43 | 1599.62 | 0.67 |
| **a0014.L** | 0.13 | 0.06 | 2.02 | 1904.29 | 0.04 |
| **a0014.Q** | 0.01 | 0.06 | 0.15 | 1691.60 | 0.88 |
| **a0014.C** | -0.04 | 0.08 | -0.49 | 1138.70 | 0.63 |
| **a0014^4** | 0.04 | 0.05 | 0.78 | 1613.74 | 0.43 |
| **dv_par_edu_birth** | 0.45 | 0.46 | 0.97 | 1664.30 | 0.33 |
| **brfed.L** | 0.04 | 0.04 | 1.09 | 2206.98 | 0.28 |
| **brfed.Q** | -0.04 | 0.05 | -0.81 | 1598.45 | 0.42 |

*Table S11.* Raw output from modified Poisson regression (with log link) of Rutter score at age 5 (d119) against median sleep duration (binarised to normal vs abnormal) derived from the Winkler et al. algorithm at age 46, adjusted for potential confounders (see main text Table 3). The parameter estimates in the "Estimate" column and the associated standard errors have not been exponentiated.

| Term | Estimate | Standard error | Statistic | Degrees of freedom | p-value |
|---|---|---|---|---|---|
| **(Intercept)** | -0.73 | 0.38 | -1.93 | 2754.78 | 0.05 |
| **BD3MRUTT** | 0.08 | 0.03 | 3.07 | 3242.76 | 0.00 |
| **a0005a** | -0.26 | 0.16 | -1.65 | 8602.01 | 0.10 |
| **a0043b.L** | 0.21 | 0.07 | 3.05 | 2835.57 | 0.00 |
| **a0043b.Q** | 0.09 | 0.09 | 1.05 | 2890.83 | 0.30 |
| **a0043b.C** | 0.04 | 0.08 | 0.48 | 3424.05 | 0.63 |
| **a0043b^4** | -0.03 | 0.10 | -0.27 | 2173.65 | 0.79 |
| **a0043b^5** | -0.06 | 0.12 | -0.53 | 2560.18 | 0.60 |
| **a0195a** | -0.05 | 0.52 | -0.10 | 4129.12 | 0.92 |
| **a0278** | -0.63 | 0.24 | -2.56 | 8191.39 | 0.01 |
| **a0014.L** | 0.19 | 0.11 | 1.82 | 2949.17 | 0.07 |
| **a0014.Q** | 0.01 | 0.06 | 0.13 | 6698.28 | 0.89 |
| **a0014.C** | 0.11 | 0.08 | 1.40 | 3172.93 | 0.16 |
| **a0014^4** | 0.05 | 0.07 | 0.82 | 2570.83 | 0.41 |
| **dv_par_edu_birth** | -1.51 | 1.35 | -1.11 | 2820.83 | 0.27 |
| **dv_sc_age_5.L** | 0.23 | 0.09 | 2.56 | 1812.99 | 0.01 |
| **dv_sc_age_5.Q** | -0.06 | 0.04 | -1.46 | 4472.01 | 0.14 |
| **e216aNone** | -0.03 | 0.05 | -0.57 | 3743.84 | 0.57 |
| **dv_cog_abil_5** | -0.90 | 0.27 | -3.26 | 1909.74 | 0.00 |
| **dv_med_51** | 0.05 | 0.06 | 0.93 | 4083.48 | 0.35 |
| **d016a** | 0.22 | 0.05 | 4.05 | 4315.35 | 0.00 |
| **sepmumbcs1** | 0.17 | 0.11 | 1.51 | 4646.41 | 0.13 |
| **crowdUp to 1** | -0.23 | 0.06 | -3.64 | 3375.70 | 0.00 |
| **brfed.L** | 0.09 | 0.05 | 1.76 | 3878.71 | 0.08 |
| **brfed.Q** | -0.05 | 0.06 | -0.74 | 3586.49 | 0.46 |
| **resmove.L** | 0.16 | 0.06 | 2.74 | 3965.33 | 0.01 |
| **resmove.Q** | -0.02 | 0.03 | -0.76 | 10634.90 | 0.45 |

*Table S12.* Raw output from modified Poisson regression (with log link) of Rutter score at age 10 (BD3MRUTT) against self-reported average sleep duration (binarised to normal vs abnormal) at age 46, adjusted for potential confounders (see main text Table 3). The parameter estimates in the "Estimate" column and the associated standard errors have not been exponentiated.

| Term | Estimate | Standard error | Statistic | Degrees of freedom | p-value |
|---|---|---|---|---|---|
| **(Intercept)** | -1.36 | 0.50 | -2.75 | 1613.17 | 0.01 |
| **BD3MRUTT** | 0.11 | 0.03 | 4.27 | 3200.09 | 0.00 |
| **a0005a** | 0.51 | 0.22 | 2.27 | 2436.07 | 0.02 |
| **a0043b.L** | 0.19 | 0.07 | 2.62 | 2662.03 | 0.01 |
| **a0043b.Q** | 0.18 | 0.10 | 1.84 | 2554.82 | 0.07 |
| **a0043b.C** | 0.04 | 0.09 | 0.48 | 2307.18 | 0.63 |
| **a0043b^4** | -0.10 | 0.09 | -1.10 | 2847.24 | 0.27 |
| **a0043b^5** | -0.07 | 0.12 | -0.56 | 3048.87 | 0.58 |
| **a0195a** | -0.01 | 0.66 | -0.02 | 2518.53 | 0.98 |
| **a0278** | 0.17 | 0.27 | 0.64 | 5734.57 | 0.53 |
| **a0014.L** | 0.26 | 0.14 | 1.80 | 1669.39 | 0.07 |
| **a0014.Q** | 0.06 | 0.08 | 0.76 | 2514.89 | 0.45 |
| **a0014.C** | 0.15 | 0.09 | 1.80 | 2751.96 | 0.07 |
| **a0014^4** | 0.09 | 0.06 | 1.63 | 4008.42 | 0.10 |
| **dv_par_edu_birth** | -0.18 | 1.04 | -0.17 | 2552.69 | 0.86 |
| **dv_sc_age_5.L** | 0.16 | 0.11 | 1.43 | 1351.56 | 0.15 |
| **dv_sc_age_5.Q** | 0.00 | 0.05 | -0.07 | 3249.24 | 0.94 |
| **e216aNone** | 0.08 | 0.08 | 1.11 | 1994.63 | 0.27 |
| **dv_cog_abil_5** | -1.65 | 0.39 | -4.25 | 1205.88 | 0.00 |
| **dv_med_51** | -0.01 | 0.10 | -0.09 | 1515.90 | 0.93 |
| **d016a** | 0.11 | 0.07 | 1.67 | 2745.67 | 0.10 |
| **sepmumbcs1** | 0.35 | 0.12 | 2.96 | 3254.24 | 0.00 |
| **crowdUp to 1** | -0.15 | 0.06 | -2.38 | 3459.86 | 0.02 |
| **brfed.L** | 0.01 | 0.06 | 0.12 | 2143.88 | 0.91 |
| **brfed.Q** | 0.00 | 0.10 | -0.05 | 1401.73 | 0.96 |
| **resmove.L** | 0.21 | 0.09 | 2.49 | 1740.00 | 0.01 |
| **resmove.Q** | 0.06 | 0.04 | 1.30 | 3862.85 | 0.20 |

*Table S13.* Raw output from modified Poisson regression (with log link) of Rutter score at age 10 (BD3MRUTT) against median sleep duration (binarised to normal vs abnormal) derived from the sleep diary at age 46, adjusted for potential confounders (see main text Table 3). The parameter estimates in the “Estimate” column and the associated standard errors have not been exponentiated.

| Term | Estimate | Standard error | Statistic | Degrees of freedom | p-value |
|---|---|---|---|---|---|
| **(Intercept)** | -0.41 | 0.23 | -1.77 | 2035.57 | 0.08 |
| **BD3MRUTT** | 0.01 | 0.02 | 0.71 | 2682.38 | 0.48 |
| **a0005a** | -0.37 | 0.10 | -3.86 | 5688.69 | 0.00 |
| **a0043b.L** | -0.04 | 0.05 | -0.72 | 1708.02 | 0.47 |
| **a0043b.Q** | -0.02 | 0.06 | -0.35 | 2180.65 | 0.73 |
| **a0043b.C** | 0.00 | 0.06 | 0.05 | 1646.00 | 0.96 |
| **a0043b^4** | -0.05 | 0.04 | -1.24 | 3578.76 | 0.22 |
| **a0043b^5** | -0.02 | 0.07 | -0.23 | 2198.01 | 0.82 |
| **a0195a** | -0.38 | 0.47 | -0.81 | 1678.63 | 0.42 |
| **a0278** | -0.22 | 0.21 | -1.06 | 2184.19 | 0.29 |
| **a0014.L** | 0.04 | 0.07 | 0.54 | 1983.44 | 0.59 |
| **a0014.Q** | 0.04 | 0.05 | 0.72 | 1964.33 | 0.47 |
| **a0014.C** | 0.01 | 0.04 | 0.19 | 2744.75 | 0.85 |
| **a0014^4** | -0.01 | 0.04 | -0.27 | 1964.85 | 0.79 |
| **dv_par_edu_birth** | -0.46 | 0.48 | -0.96 | 2724.66 | 0.34 |
| **dv_sc_age_5.L** | 0.00 | 0.05 | 0.02 | 1959.66 | 0.99 |
| **dv_sc_age_5.Q** | 0.00 | 0.03 | -0.04 | 3829.46 | 0.96 |
| **e216aNone** | -0.01 | 0.05 | -0.24 | 1562.00 | 0.81 |
| **dv_cog_abil_5** | -0.05 | 0.25 | -0.22 | 964.99 | 0.83 |
| **dv_med_51** | 0.01 | 0.04 | 0.33 | 3073.06 | 0.74 |
| **d016a** | 0.06 | 0.03 | 1.83 | 3472.36 | 0.07 |
| **sepmumbcs1** | -0.04 | 0.09 | -0.45 | 2526.33 | 0.65 |
| **crowdUp to 1** | -0.07 | 0.05 | -1.46 | 2239.40 | 0.15 |
| **brfed.L** | 0.03 | 0.03 | 0.99 | 2074.51 | 0.32 |
| **brfed.Q** | -0.01 | 0.03 | -0.43 | 3357.43 | 0.67 |
| **resmove.L** | -0.02 | 0.04 | -0.51 | 2570.02 | 0.61 |
| **resmove.Q** | 0.06 | 0.02 | 2.57 | 4381.88 | 0.01 |

*Table S14.* Raw output from modified Poisson regression (with log link) of Rutter score at age 10 (BD3MRUTT) against median sleep duration (binarised to normal vs abnormal) derived from the activPAL algorithm at age 46, adjusted for potential confounders (see main text Table 3). The parameter estimates in the "Estimate" column and the associated standard errors have not been exponentiated.

| Term | Estimate | Standard error | Statistic | Degrees of freedom | p-value |
|---|---|---|---|---|---|
| **(Intercept)** | -1.09 | 0.31 | -3.47 | 1527.66 | 0.00 |
| **BD3MRUTT** | 0.02 | 0.02 | 1.09 | 1673.88 | 0.28 |
| **a0005a** | -0.06 | 0.17 | -0.35 | 1743.57 | 0.72 |
| **a0043b.L** | 0.01 | 0.06 | 0.20 | 1695.90 | 0.84 |
| **a0043b.Q** | -0.07 | 0.07 | -1.09 | 1814.06 | 0.27 |
| **a0043b.C** | -0.18 | 0.06 | -2.79 | 1745.36 | 0.01 |
| **a0043b^4** | 0.11 | 0.06 | 1.74 | 2080.17 | 0.08 |
| **a0043b^5** | 0.03 | 0.08 | 0.42 | 2144.89 | 0.68 |
| **a0195a** | -0.22 | 0.46 | -0.47 | 2084.77 | 0.64 |
| **a0278** | 0.31 | 0.18 | 1.77 | 4237.23 | 0.08 |
| **a0014.L** | 0.05 | 0.10 | 0.56 | 1418.58 | 0.58 |
| **a0014.Q** | 0.05 | 0.07 | 0.69 | 1463.83 | 0.49 |
| **a0014.C** | -0.02 | 0.05 | -0.38 | 2849.70 | 0.71 |
| **a0014^4** | -0.01 | 0.04 | -0.21 | 2799.13 | 0.83 |
| **dv_par_edu_birth** | -0.18 | 0.40 | -0.45 | 4241.08 | 0.65 |
| **dv_sc_age_5.L** | 0.17 | 0.07 | 2.34 | 1241.35 | 0.02 |
| **dv_sc_age_5.Q** | -0.04 | 0.04 | -1.11 | 1864.87 | 0.27 |
| **e216aNone** | 0.06 | 0.05 | 1.19 | 1789.72 | 0.24 |
| **dv_cog_abil_5** | -0.17 | 0.23 | -0.74 | 1172.90 | 0.46 |
| **dv_med_51** | 0.05 | 0.05 | 0.93 | 1967.23 | 0.35 |
| **d016a** | 0.09 | 0.04 | 2.03 | 2361.48 | 0.04 |
| **sepmumbcs1** | -0.03 | 0.13 | -0.25 | 1683.64 | 0.80 |
| **crowdUp to 1** | 0.01 | 0.06 | 0.14 | 1772.97 | 0.89 |
| **brfed.L** | 0.02 | 0.04 | 0.36 | 1584.76 | 0.72 |
| **brfed.Q** | 0.04 | 0.05 | 0.90 | 2215.08 | 0.37 |
| **resmove.L** | 0.02 | 0.04 | 0.37 | 3072.42 | 0.71 |
| **resmove.Q** | -0.06 | 0.03 | -2.06 | 3541.08 | 0.04 |

*Table S15.* Raw output from modified Poisson regression (with log link) of Rutter score at age 10 (BD3MRUTT) against median sleep duration (binarised to normal vs abnormal) derived from the van der Berg et al. algorithm at age 46, adjusted for potential confounders (see main text Table 3). The parameter estimates in the “Estimate” column and the associated standard errors have not been exponentiated.

| Term | Estimate | Standard error | Statistic | Degrees of freedom | p-value |
|---|---|---|---|---|---|
| **(Intercept)** | -0.96 | 0.21 | -4.56 | 2676.77 | 0.00 |
| **BD3MRUTT** | 0.03 | 0.02 | 1.49 | 2203.63 | 0.14 |
| **a0005a** | 0.12 | 0.20 | 0.61 | 1267.42 | 0.54 |
| **a0043b.L** | -0.03 | 0.04 | -0.73 | 2745.95 | 0.47 |
| **a0043b.Q** | 0.02 | 0.06 | 0.32 | 2166.84 | 0.75 |
| **a0043b.C** | 0.06 | 0.04 | 1.59 | 4275.25 | 0.11 |
| **a0043b^4** | 0.03 | 0.05 | 0.54 | 2585.33 | 0.59 |
| **a0043b^5** | 0.04 | 0.07 | 0.54 | 2487.49 | 0.59 |
| **a0195a** | -0.07 | 0.34 | -0.21 | 3668.59 | 0.83 |
| **a0278** | -0.07 | 0.28 | -0.24 | 1507.13 | 0.81 |
| **a0014.L** | 0.08 | 0.08 | 1.06 | 1816.10 | 0.29 |
| **a0014.Q** | 0.00 | 0.07 | 0.07 | 1466.33 | 0.95 |
| **a0014.C** | -0.04 | 0.09 | -0.42 | 1108.92 | 0.67 |
| **a0014^4** | 0.04 | 0.05 | 0.73 | 1624.08 | 0.47 |
| **dv_par_edu_birth** | 0.40 | 0.45 | 0.89 | 1737.63 | 0.37 |
| **dv_sc_age_5.L** | 0.01 | 0.08 | 0.16 | 1058.70 | 0.87 |
| **dv_sc_age_5.Q** | -0.01 | 0.05 | -0.21 | 1462.78 | 0.83 |
| **e216aNone** | -0.13 | 0.06 | -2.38 | 1317.84 | 0.02 |
| **dv_cog_abil_5** | -0.33 | 0.25 | -1.33 | 1022.04 | 0.18 |
| **dv_med_51** | 0.04 | 0.04 | 0.91 | 2664.85 | 0.36 |
| **d016a** | 0.19 | 0.05 | 3.54 | 1451.95 | 0.00 |
| **sepmumbcs1** | 0.21 | 0.09 | 2.20 | 1928.86 | 0.03 |
| **crowdUp to 1** | 0.05 | 0.05 | 1.14 | 2487.70 | 0.25 |
| **brfed.L** | 0.04 | 0.04 | 1.03 | 2253.04 | 0.30 |
| **brfed.Q** | -0.05 | 0.05 | -0.88 | 1607.28 | 0.38 |
| **resmove.L** | 0.02 | 0.06 | 0.40 | 1364.41 | 0.69 |
| **resmove.Q** | 0.02 | 0.02 | 1.02 | 5839.80 | 0.31 |

*Table S16.* Raw output from modified Poisson regression (with log link) of Rutter score at age 10 (BD3MRUTT) against median sleep duration (binarised to normal vs abnormal) derived from the Winkler et al. algorithm at age 46, adjusted for potential confounders (see main text Table 3). The parameter estimates in the "Estimate" column and the associated standard errors have not been exponentiated.

| Term | Estimate | Standard error | Statistic | Degrees of freedom | p-value |
|---|---|---|---|---|---|
| **(Intercept)** | -0.87 | 0.41 | -2.13 | 2355.65 | 0.03 |
| **dv_cds_10** | 0.15 | 0.03 | 4.45 | 1725.68 | 0.00 |
| **a0005a** | -0.29 | 0.16 | -1.87 | 8515.41 | 0.06 |
| **a0043b.L** | 0.20 | 0.07 | 2.94 | 2775.12 | 0.00 |
| **a0043b.Q** | 0.09 | 0.09 | 1.05 | 2791.38 | 0.30 |
| **a0043b.C** | 0.03 | 0.08 | 0.41 | 3414.12 | 0.68 |
| **a0043b^4** | -0.03 | 0.10 | -0.26 | 2148.59 | 0.79 |
| **a0043b^5** | -0.06 | 0.12 | -0.49 | 2595.59 | 0.63 |
| **a0195a** | 0.02 | 0.53 | 0.03 | 3867.79 | 0.98 |
| **a0278** | -0.60 | 0.24 | -2.54 | 9383.60 | 0.01 |
| **a0014.L** | 0.19 | 0.11 | 1.84 | 2875.32 | 0.07 |
| **a0014.Q** | 0.01 | 0.06 | 0.17 | 6796.45 | 0.87 |
| **a0014.C** | 0.11 | 0.08 | 1.35 | 3155.82 | 0.18 |
| **a0014^4** | 0.05 | 0.07 | 0.70 | 2579.56 | 0.48 |
| **dv_par_edu_birth** | -1.41 | 1.34 | -1.05 | 2800.99 | 0.29 |
| **dv_sc_age_5.L** | 0.22 | 0.09 | 2.48 | 1808.40 | 0.01 |
| **dv_sc_age_5.Q** | -0.06 | 0.04 | -1.43 | 4603.72 | 0.15 |
| **e216aNone** | -0.03 | 0.05 | -0.62 | 3755.95 | 0.54 |
| **dv_cog_abil_5** | -0.74 | 0.28 | -2.64 | 1886.37 | 0.01 |
| **dv_med_51** | 0.06 | 0.06 | 0.97 | 4252.09 | 0.33 |
| **d016a** | 0.21 | 0.06 | 3.61 | 3358.88 | 0.00 |
| **sepmumbcs1** | 0.14 | 0.11 | 1.24 | 4450.43 | 0.21 |
| **crowdUp to 1** | -0.22 | 0.06 | -3.61 | 3432.12 | 0.00 |
| **brfed.L** | 0.09 | 0.05 | 1.84 | 3901.94 | 0.07 |
| **brfed.Q** | -0.05 | 0.06 | -0.75 | 3610.53 | 0.45 |
| **resmove.L** | 0.15 | 0.06 | 2.75 | 4812.97 | 0.01 |
| **resmove.Q** | -0.02 | 0.03 | -0.69 | 10364.55 | 0.49 |

*Table S17.* Raw output from modified Poisson regression (with log link) of Child Development Scale score at age 10 (dv_cds_10) against self-reported average sleep duration (binarised to normal vs abnormal) at age 46, adjusted for potential confounders (see main text Table 3). The parameter estimates in the “Estimate” column and the associated standard errors have not been exponentiated.

| Term | Estimate | Standard error | Statistic | Degrees of freedom | p-value |
|---|---|---|---|---|---|
| **(Intercept)** | -1.45 | 0.50 | -2.90 | 1590.08 | 0.00 |
| **dv_cds_10** | 0.15 | 0.03 | 4.26 | 1841.00 | 0.00 |
| **a0005a** | 0.46 | 0.23 | 2.05 | 2412.93 | 0.04 |
| **a0043b.L** | 0.18 | 0.07 | 2.56 | 2586.17 | 0.01 |
| **a0043b.Q** | 0.19 | 0.10 | 1.85 | 2484.33 | 0.06 |
| **a0043b.C** | 0.04 | 0.09 | 0.45 | 2423.55 | 0.65 |
| **a0043b^4** | -0.10 | 0.10 | -1.09 | 2773.14 | 0.27 |
| **a0043b^5** | -0.06 | 0.12 | -0.51 | 3013.77 | 0.61 |
| **a0195a** | 0.05 | 0.66 | 0.07 | 2554.06 | 0.94 |
| **a0278** | 0.19 | 0.26 | 0.71 | 6183.79 | 0.48 |
| **a0014.L** | 0.26 | 0.14 | 1.84 | 1667.69 | 0.07 |
| **a0014.Q** | 0.06 | 0.08 | 0.77 | 2512.41 | 0.44 |
| **a0014.C** | 0.15 | 0.09 | 1.75 | 2707.98 | 0.08 |
| **a0014^4** | 0.09 | 0.06 | 1.48 | 3882.38 | 0.14 |
| **dv_par_edu_birth** | -0.11 | 1.02 | -0.10 | 2552.40 | 0.92 |
| **dv_sc_age_5.L** | 0.16 | 0.11 | 1.43 | 1376.35 | 0.15 |
| **dv_sc_age_5.Q** | 0.00 | 0.05 | -0.07 | 3252.06 | 0.95 |
| **e216aNone** | 0.08 | 0.08 | 1.05 | 2036.71 | 0.30 |
| **dv_cog_abil_5** | -1.53 | 0.39 | -3.96 | 1236.10 | 0.00 |
| **dv_med_51** | 0.00 | 0.10 | -0.04 | 1498.52 | 0.97 |
| **d016a** | 0.11 | 0.06 | 1.74 | 2891.65 | 0.08 |
| **sepmumbcs1** | 0.33 | 0.12 | 2.71 | 3061.06 | 0.01 |
| **crowdUp to 1** | -0.15 | 0.06 | -2.39 | 3526.53 | 0.02 |
| **brfed.L** | 0.01 | 0.06 | 0.18 | 2164.43 | 0.86 |
| **brfed.Q** | -0.01 | 0.10 | -0.07 | 1427.08 | 0.94 |
| **resmove.L** | 0.21 | 0.08 | 2.50 | 1813.68 | 0.01 |
| **resmove.Q** | 0.06 | 0.04 | 1.42 | 3989.44 | 0.15 |

*Table S18.* Raw output from modified Poisson regression (with log link) of Child Development Scale score at age 10 (dv_cds_10) against median sleep duration (binarised to normal vs abnormal) derived from the sleep diary at age 46, adjusted for potential confounders (see main text Table 3). The parameter estimates in the “Estimate” column and the associated standard errors have not been exponentiated.

| Term | Estimate | Standard error | Statistic | Degrees of freedom | p-value |
|---|---|---|---|---|---|
| **(Intercept)** | -0.39 | 0.23 | -1.67 | 2041.52 | 0.09 |
| **dv_cds_10** | -0.01 | 0.01 | -0.91 | 3429.27 | 0.36 |
| **a0005a** | -0.37 | 0.10 | -3.91 | 5772.42 | 0.00 |
| **a0043b.L** | -0.03 | 0.05 | -0.69 | 1749.06 | 0.49 |
| **a0043b.Q** | -0.02 | 0.06 | -0.34 | 2174.05 | 0.73 |
| **a0043b.C** | 0.00 | 0.06 | 0.07 | 1635.59 | 0.94 |
| **a0043b^4** | -0.05 | 0.04 | -1.23 | 3542.18 | 0.22 |
| **a0043b^5** | -0.02 | 0.07 | -0.24 | 2199.44 | 0.81 |
| **a0195a** | -0.38 | 0.47 | -0.82 | 1671.10 | 0.41 |
| **a0278** | -0.22 | 0.21 | -1.07 | 2161.67 | 0.29 |
| **a0014.L** | 0.04 | 0.07 | 0.56 | 1977.61 | 0.58 |
| **a0014.Q** | 0.04 | 0.05 | 0.70 | 1968.85 | 0.48 |
| **a0014.C** | 0.01 | 0.04 | 0.19 | 2754.57 | 0.85 |
| **a0014^4** | -0.01 | 0.04 | -0.25 | 1977.84 | 0.80 |
| **dv_par_edu_birth** | -0.47 | 0.48 | -0.98 | 2703.80 | 0.33 |
| **dv_sc_age_5.L** | 0.00 | 0.05 | 0.08 | 1923.40 | 0.94 |
| **dv_sc_age_5.Q** | 0.00 | 0.03 | -0.07 | 3811.57 | 0.95 |
| **e216aNone** | -0.01 | 0.05 | -0.26 | 1554.92 | 0.79 |
| **dv_cog_abil_5** | -0.08 | 0.24 | -0.33 | 992.68 | 0.74 |
| **dv_med_51** | 0.01 | 0.04 | 0.38 | 3024.85 | 0.71 |
| **d016a** | 0.07 | 0.03 | 2.00 | 3717.57 | 0.05 |
| **sepmumbcs1** | -0.04 | 0.09 | -0.41 | 2469.23 | 0.68 |
| **crowdUp to 1** | -0.07 | 0.05 | -1.50 | 2216.75 | 0.13 |
| **brfed.L** | 0.03 | 0.03 | 0.99 | 2061.85 | 0.32 |
| **brfed.Q** | -0.02 | 0.03 | -0.47 | 3446.73 | 0.64 |
| **resmove.L** | -0.02 | 0.04 | -0.45 | 2650.78 | 0.66 |
| **resmove.Q** | 0.06 | 0.02 | 2.58 | 4275.26 | 0.01 |

*Table S19.* Raw output from modified Poisson regression (with log link) of Child Development Scale score at age 10 (dv_cds_10) against median sleep duration (binarised to normal vs abnormal) derived from the activPAL algorithm at age 46, adjusted for potential confounders (see main text Table 3). The parameter estimates in the “Estimate” column and the associated standard errors have not been exponentiated.

| Term | Estimate | Standard error | Statistic | Degrees of freedom | p-value |
|---|---|---|---|---|---|
| **(Intercept)** | -1.09 | 0.33 | -3.33 | 1446.43 | 0.00 |
| **dv_cds_10** | 0.02 | 0.03 | 0.63 | 1289.33 | 0.53 |
| **a0005a** | -0.07 | 0.17 | -0.41 | 1760.80 | 0.68 |
| **a0043b.L** | 0.01 | 0.06 | 0.21 | 1716.08 | 0.83 |
| **a0043b.Q** | -0.07 | 0.07 | -1.09 | 1832.19 | 0.27 |
| **a0043b.C** | -0.18 | 0.07 | -2.76 | 1723.49 | 0.01 |
| **a0043b^4** | 0.11 | 0.06 | 1.74 | 2079.71 | 0.08 |
| **a0043b^5** | 0.03 | 0.08 | 0.42 | 2165.19 | 0.67 |
| **a0195a** | -0.21 | 0.47 | -0.46 | 2030.55 | 0.65 |
| **a0278** | 0.31 | 0.18 | 1.77 | 4252.61 | 0.08 |
| **a0014.L** | 0.05 | 0.10 | 0.57 | 1417.90 | 0.57 |
| **a0014.Q** | 0.05 | 0.07 | 0.68 | 1445.87 | 0.50 |
| **a0014.C** | -0.02 | 0.05 | -0.38 | 2820.57 | 0.70 |
| **a0014^4** | -0.01 | 0.04 | -0.23 | 2763.53 | 0.82 |
| **dv_par_edu_birth** | -0.18 | 0.41 | -0.44 | 4093.69 | 0.66 |
| **dv_sc_age_5.L** | 0.17 | 0.07 | 2.40 | 1275.28 | 0.02 |
| **dv_sc_age_5.Q** | -0.04 | 0.04 | -1.12 | 1851.46 | 0.26 |
| **e216aNone** | 0.06 | 0.05 | 1.15 | 1770.76 | 0.25 |
| **dv_cog_abil_5** | -0.17 | 0.24 | -0.70 | 1164.36 | 0.49 |
| **dv_med_51** | 0.05 | 0.05 | 0.96 | 1947.62 | 0.34 |
| **d016a** | 0.09 | 0.04 | 2.03 | 2224.67 | 0.04 |
| **sepmumbcs1** | -0.03 | 0.13 | -0.26 | 1665.58 | 0.79 |
| **crowdUp to 1** | 0.01 | 0.06 | 0.13 | 1848.65 | 0.90 |
| **brfed.L** | 0.02 | 0.04 | 0.39 | 1589.79 | 0.70 |
| **brfed.Q** | 0.04 | 0.05 | 0.87 | 2209.17 | 0.38 |
| **resmove.L** | 0.02 | 0.04 | 0.40 | 3056.12 | 0.69 |
| **resmove.Q** | -0.05 | 0.03 | -2.03 | 3601.81 | 0.04 |

*Table S20.* Raw output from modified Poisson regression (with log link) of Child Development Scale score at age 10 (dv_cds_10) against median sleep duration (binarised to normal vs abnormal) derived from the van der Berg et al. algorithm at age 46, adjusted for potential confounders (see main text Table 3). The parameter estimates in the "Estimate" column and the associated standard errors have not been exponentiated.

| Term | Estimate | Standard error | Statistic | Degrees of freedom | p-value |
|---|---|---|---|---|---|
| **(Intercept)** | -1.05 | 0.22 | -4.68 | 2345.23 | 0.00 |
| **dv_cds_10** | 0.10 | 0.03 | 3.81 | 1321.41 | 0.00 |
| **a0005a** | 0.11 | 0.20 | 0.54 | 1255.20 | 0.59 |
| **a0043b.L** | -0.04 | 0.04 | -0.88 | 2861.34 | 0.38 |
| **a0043b.Q** | 0.02 | 0.06 | 0.32 | 2163.78 | 0.75 |
| **a0043b.C** | 0.06 | 0.04 | 1.45 | 3981.10 | 0.15 |
| **a0043b^4** | 0.03 | 0.05 | 0.56 | 2580.54 | 0.58 |
| **a0043b^5** | 0.04 | 0.07 | 0.59 | 2509.95 | 0.56 |
| **a0195a** | -0.03 | 0.34 | -0.09 | 3543.65 | 0.93 |
| **a0278** | -0.05 | 0.28 | -0.20 | 1486.32 | 0.84 |
| **a0014.L** | 0.08 | 0.08 | 1.03 | 1816.85 | 0.30 |
| **a0014.Q** | 0.01 | 0.07 | 0.09 | 1431.88 | 0.93 |
| **a0014.C** | -0.04 | 0.09 | -0.45 | 1110.89 | 0.65 |
| **a0014^4** | 0.03 | 0.05 | 0.63 | 1661.91 | 0.53 |
| **dv_par_edu_birth** | 0.45 | 0.45 | 0.99 | 1699.94 | 0.32 |
| **dv_sc_age_5.L** | 0.01 | 0.08 | 0.08 | 1048.60 | 0.94 |
| **dv_sc_age_5.Q** | -0.01 | 0.05 | -0.18 | 1464.02 | 0.86 |
| **e216aNone** | -0.13 | 0.06 | -2.41 | 1332.81 | 0.02 |
| **dv_cog_abil_5** | -0.22 | 0.25 | -0.86 | 1033.84 | 0.39 |
| **dv_med_51** | 0.04 | 0.04 | 0.91 | 2980.42 | 0.36 |
| **d016a** | 0.18 | 0.05 | 3.43 | 1472.68 | 0.00 |
| **sepmumbcs1** | 0.19 | 0.10 | 1.99 | 1916.92 | 0.05 |
| **crowdUp to 1** | 0.06 | 0.05 | 1.25 | 2469.37 | 0.21 |
| **brfed.L** | 0.04 | 0.04 | 1.08 | 2210.93 | 0.28 |
| **brfed.Q** | -0.04 | 0.05 | -0.87 | 1610.85 | 0.38 |
| **resmove.L** | 0.02 | 0.06 | 0.30 | 1406.56 | 0.76 |
| **resmove.Q** | 0.02 | 0.02 | 1.04 | 5527.14 | 0.30 |

*Table S21.* Raw output from modified Poisson regression (with log link) of Child Development Scale score at age 10 (dv_cds_10) against median sleep duration (binarised to normal vs abnormal) derived from the Winkler et al. algorithm at age 46, adjusted for potential confounders (see main text Table 3). The parameter estimates in the “Estimate” column and the associated standard errors have not been exponentiated.

| Term | Estimate | Standard error | Statistic | Degrees of freedom | p-value |
|---|---|---|---|---|---|
| **(Intercept)** | -0.69 | 0.42 | -1.64 | 2443.53 | 0.10 |
| **BD4MAL** | 0.17 | 0.03 | 4.88 | 1508.67 | 0.00 |
| **a0005a** | -0.21 | 0.15 | -1.34 | 8605.60 | 0.18 |
| **a0043b.L** | 0.18 | 0.07 | 2.67 | 3015.10 | 0.01 |
| **a0043b.Q** | 0.12 | 0.09 | 1.26 | 2589.26 | 0.21 |
| **a0043b.C** | 0.02 | 0.08 | 0.21 | 2720.35 | 0.83 |
| **a0043b^4** | -0.05 | 0.10 | -0.46 | 2176.97 | 0.64 |
| **a0043b^5** | -0.04 | 0.12 | -0.32 | 2630.65 | 0.75 |
| **a0195a** | -0.07 | 0.50 | -0.15 | 4487.91 | 0.88 |
| **a0278** | -0.53 | 0.25 | -2.14 | 7466.66 | 0.03 |
| **a0014.L** | 0.14 | 0.10 | 1.34 | 3341.06 | 0.18 |
| **a0014.Q** | 0.00 | 0.06 | 0.03 | 6039.94 | 0.98 |
| **a0014.C** | 0.11 | 0.08 | 1.47 | 3434.52 | 0.14 |
| **a0014^4** | 0.04 | 0.07 | 0.56 | 2717.15 | 0.57 |
| **dv_par_edu_birth** | -1.12 | 1.23 | -0.91 | 2851.17 | 0.36 |
| **dv_sc_age_5.L** | 0.16 | 0.10 | 1.56 | 1704.86 | 0.12 |
| **dv_sc_age_5.Q** | -0.05 | 0.05 | -1.02 | 3580.71 | 0.31 |
| **e216aNone** | -0.02 | 0.06 | -0.29 | 3486.15 | 0.77 |
| **dv_cog_abil_5** | -0.60 | 0.29 | -2.11 | 1917.56 | 0.03 |
| **dv_med_51** | 0.03 | 0.06 | 0.57 | 3769.70 | 0.57 |
| **d016a** | 0.21 | 0.05 | 3.86 | 4218.88 | 0.00 |
| **dv_bas_g** | -0.59 | 0.23 | -2.63 | 2441.87 | 0.01 |
| **dv_med_101** | 0.01 | 0.04 | 0.31 | 8090.14 | 0.75 |
| **bmi** | 0.00 | 0.27 | 0.00 | 2891.00 | 1.00 |
| **fclrg90.L** | 0.05 | 0.09 | 0.56 | 3521.89 | 0.57 |
| **fclrg90.Q** | 0.02 | 0.06 | 0.44 | 6723.71 | 0.66 |
| **fclrg90.C** | -0.07 | 0.07 | -0.95 | 3973.66 | 0.34 |
| **fclrg90^4** | 0.02 | 0.06 | 0.42 | 4886.92 | 0.67 |
| **tenure.L** | 0.12 | 0.05 | 2.25 | 2673.46 | 0.02 |
| **tenure.Q** | -0.06 | 0.07 | -0.89 | 3458.34 | 0.38 |
| **divorce1** | 0.09 | 0.06 | 1.56 | 6279.39 | 0.12 |
| **sepmumbcs1** | 0.12 | 0.11 | 1.07 | 5755.57 | 0.29 |
| **prmnh1** | 0.21 | 0.13 | 1.68 | 2367.92 | 0.09 |
| **crowdUp to 1** | -0.15 | 0.06 | -2.70 | 5486.98 | 0.01 |
| **ameniNo occasions** | -0.07 | 0.12 | -0.58 | 2533.76 | 0.56 |
| **brfed.L** | 0.07 | 0.05 | 1.40 | 3577.51 | 0.16 |
| **brfed.Q** | -0.04 | 0.06 | -0.69 | 3770.04 | 0.49 |
| **resmove.L** | 0.13 | 0.06 | 2.09 | 3613.88 | 0.04 |
| **resmove.Q** | -0.02 | 0.03 | -0.68 | 8587.13 | 0.49 |

*Table S22.* Raw output from modified Poisson regression (with log link) of Malaise Inventory score at age 16 (BD4MAL) against self-reported average sleep duration (binarised to normal vs abnormal) at age 46, adjusted for potential confounders (see main text Table 3). The parameter estimates in the “Estimate” column and the associated standard errors have not been exponentiated.

| Term | Estimate | Standard error | Statistic | Degrees of freedom | p-value |
|---|---|---|---|---|---|
| **(Intercept)** | -1.11 | 0.46 | -2.41 | 1992.21 | 0.02 |
| **BD4MAL** | 0.06 | 0.03 | 1.88 | 1801.03 | 0.06 |
| **a0005a** | 0.49 | 0.22 | 2.22 | 2518.52 | 0.03 |
| **a0043b.L** | 0.17 | 0.07 | 2.41 | 2680.54 | 0.02 |
| **a0043b.Q** | 0.19 | 0.10 | 1.94 | 2608.28 | 0.05 |
| **a0043b.C** | 0.04 | 0.09 | 0.41 | 2236.76 | 0.68 |
| **a0043b^4** | -0.11 | 0.09 | -1.21 | 2936.42 | 0.23 |
| **a0043b^5** | -0.06 | 0.12 | -0.48 | 2978.72 | 0.63 |
| **a0195a** | -0.06 | 0.66 | -0.09 | 2525.24 | 0.93 |
| **a0278** | 0.30 | 0.28 | 1.06 | 5024.91 | 0.29 |
| **a0014.L** | 0.17 | 0.15 | 1.17 | 1696.05 | 0.24 |
| **a0014.Q** | 0.05 | 0.08 | 0.69 | 2777.47 | 0.49 |
| **a0014.C** | 0.15 | 0.09 | 1.65 | 2409.81 | 0.10 |
| **a0014^4** | 0.08 | 0.06 | 1.44 | 4088.01 | 0.15 |
| **dv_par_edu_birth** | 0.11 | 0.91 | 0.12 | 2639.21 | 0.90 |
| **dv_sc_age_5.L** | 0.08 | 0.13 | 0.60 | 1253.59 | 0.55 |
| **dv_sc_age_5.Q** | 0.01 | 0.05 | 0.27 | 3847.95 | 0.79 |
| **e216aNone** | 0.08 | 0.07 | 1.13 | 2163.36 | 0.26 |
| **dv_cog_abil_5** | -1.43 | 0.44 | -3.23 | 1106.92 | 0.00 |
| **dv_med_51** | -0.02 | 0.10 | -0.18 | 1461.12 | 0.86 |
| **d016a** | 0.12 | 0.07 | 1.75 | 2724.45 | 0.08 |
| **dv_bas_g** | -0.58 | 0.32 | -1.84 | 1532.39 | 0.07 |
| **dv_med_101** | 0.05 | 0.06 | 0.76 | 2994.75 | 0.45 |
| **bmi** | -0.49 | 0.29 | -1.70 | 2931.46 | 0.09 |
| **fclrg90.L** | 0.24 | 0.10 | 2.42 | 2887.26 | 0.02 |
| **fclrg90.Q** | 0.05 | 0.07 | 0.69 | 3597.75 | 0.49 |
| **fclrg90.C** | -0.04 | 0.11 | -0.35 | 1838.92 | 0.72 |
| **fclrg90^4** | -0.02 | 0.08 | -0.23 | 1965.83 | 0.81 |
| **tenure.L** | 0.06 | 0.07 | 0.82 | 1664.26 | 0.41 |
| **tenure.Q** | -0.05 | 0.09 | -0.54 | 2004.49 | 0.59 |
| **divorce1** | 0.04 | 0.07 | 0.62 | 3537.94 | 0.53 |
| **sepmumbcs1** | 0.34 | 0.14 | 2.53 | 2669.57 | 0.01 |
| **prmnh1** | 0.37 | 0.14 | 2.59 | 1836.06 | 0.01 |
| **crowdUp to 1** | -0.10 | 0.06 | -1.60 | 3807.24 | 0.11 |
| **ameniNo occasions** | 0.01 | 0.10 | 0.05 | 4312.02 | 0.96 |
| **brfed.L** | -0.01 | 0.06 | -0.12 | 2173.04 | 0.90 |
| **brfed.Q** | 0.00 | 0.11 | -0.02 | 1362.20 | 0.99 |
| **resmove.L** | 0.20 | 0.09 | 2.33 | 1747.39 | 0.02 |
| **resmove.Q** | 0.06 | 0.04 | 1.38 | 3581.30 | 0.17 |

*Table S23.* Raw output from modified Poisson regression (with log link) of Malaise Inventory score at age 16 (BD4MAL) against median sleep duration (binarised to normal vs abnormal) derived from the sleep diary at age 46, adjusted for potential confounders (see main text Table 3). The parameter estimates in the “Estimate” column and the associated standard errors have not been exponentiated.

| Term | Estimate | Standard error | Statistic | Degrees of freedom | p-value |
|---|---|---|---|---|---|
| **(Intercept)** | -0.36 | 0.26 | -1.39 | 1945.32 | 0.16 |
| **BD4MAL** | 0.01 | 0.02 | 0.57 | 1744.73 | 0.57 |
| **a0005a** | -0.35 | 0.10 | -3.55 | 4848.87 | 0.00 |
| **a0043b.L** | -0.05 | 0.05 | -0.99 | 1827.99 | 0.32 |
| **a0043b.Q** | -0.02 | 0.06 | -0.35 | 2163.39 | 0.72 |
| **a0043b.C** | 0.00 | 0.06 | -0.01 | 1681.63 | 0.99 |
| **a0043b^4** | -0.06 | 0.04 | -1.32 | 3580.45 | 0.19 |
| **a0043b^5** | -0.01 | 0.07 | -0.16 | 2099.48 | 0.87 |
| **a0195a** | -0.37 | 0.46 | -0.81 | 1675.18 | 0.42 |
| **a0278** | -0.21 | 0.20 | -1.06 | 2412.07 | 0.29 |
| **a0014.L** | 0.02 | 0.07 | 0.22 | 1961.90 | 0.83 |
| **a0014.Q** | 0.03 | 0.05 | 0.59 | 1996.27 | 0.55 |
| **a0014.C** | 0.00 | 0.05 | 0.05 | 2569.62 | 0.96 |
| **a0014^4** | -0.01 | 0.04 | -0.27 | 1753.27 | 0.79 |
| **dv_par_edu_birth** | -0.38 | 0.46 | -0.83 | 2720.04 | 0.40 |
| **dv_sc_age_5.L** | -0.02 | 0.07 | -0.27 | 1473.17 | 0.79 |
| **dv_sc_age_5.Q** | 0.00 | 0.03 | -0.15 | 2880.54 | 0.88 |
| **e216aNone** | -0.01 | 0.05 | -0.15 | 1553.55 | 0.88 |
| **dv_cog_abil_5** | 0.04 | 0.27 | 0.15 | 893.37 | 0.88 |
| **dv_med_51** | 0.00 | 0.04 | 0.13 | 3065.70 | 0.89 |
| **d016a** | 0.06 | 0.03 | 1.68 | 3508.20 | 0.09 |
| **dv_bas_g** | -0.23 | 0.16 | -1.42 | 1744.62 | 0.16 |
| **dv_med_101** | 0.01 | 0.03 | 0.33 | 2922.12 | 0.74 |
| **bmi** | 0.19 | 0.17 | 1.13 | 2183.50 | 0.26 |
| **fclrg90.L** | 0.03 | 0.06 | 0.45 | 2495.54 | 0.65 |
| **fclrg90.Q** | 0.05 | 0.04 | 1.06 | 2481.91 | 0.29 |
| **fclrg90.C** | 0.06 | 0.04 | 1.53 | 4089.50 | 0.13 |
| **fclrg90^4** | 0.01 | 0.05 | 0.26 | 1743.54 | 0.79 |
| **tenure.L** | 0.01 | 0.03 | 0.47 | 2363.95 | 0.64 |
| **tenure.Q** | -0.03 | 0.05 | -0.69 | 2273.54 | 0.49 |
| **divorce1** | 0.03 | 0.04 | 0.75 | 3959.77 | 0.45 |
| **sepmumbcs1** | -0.06 | 0.09 | -0.68 | 2906.20 | 0.50 |
| **prmnh1** | 0.18 | 0.10 | 1.85 | 1730.12 | 0.06 |
| **crowdUp to 1** | -0.05 | 0.05 | -0.87 | 1855.08 | 0.38 |
| **ameniNo occasions** | -0.08 | 0.07 | -1.06 | 2147.60 | 0.29 |
| **brfed.L** | 0.03 | 0.03 | 0.81 | 2084.67 | 0.42 |
| **brfed.Q** | -0.01 | 0.03 | -0.40 | 3445.57 | 0.69 |
| **resmove.L** | -0.03 | 0.04 | -0.74 | 2766.51 | 0.46 |
| **resmove.Q** | 0.06 | 0.02 | 2.49 | 4256.02 | 0.01 |

*Table S24.* Raw output from modified Poisson regression (with log link) of Malaise Inventory score at age 16 (BD4MAL) against median sleep duration (binarised to normal vs abnormal) derived from the activPAL algorithm at age 46, adjusted for potential confounders (see main text Table 3). The parameter estimates in the “Estimate” column and the associated standard errors have not been exponentiated.

| Term | Estimate | Standard error | Statistic | Degrees of freedom | p-value |
|---|---|---|---|---|---|
| **(Intercept)** | -0.89 | 0.34 | -2.60 | 1500.62 | 0.01 |
| **BD4MAL** | 0.00 | 0.02 | 0.15 | 1541.28 | 0.88 |
| **a0005a** | -0.01 | 0.17 | -0.08 | 1753.42 | 0.93 |
| **a0043b.L** | 0.01 | 0.06 | 0.12 | 1592.59 | 0.91 |
| **a0043b.Q** | -0.06 | 0.07 | -0.90 | 1786.49 | 0.37 |
| **a0043b.C** | -0.18 | 0.07 | -2.80 | 1730.46 | 0.01 |
| **a0043b^4** | 0.10 | 0.06 | 1.61 | 2069.00 | 0.11 |
| **a0043b^5** | 0.04 | 0.08 | 0.47 | 2149.16 | 0.64 |
| **a0195a** | -0.25 | 0.47 | -0.54 | 1972.69 | 0.59 |
| **a0278** | 0.36 | 0.18 | 2.01 | 3930.01 | 0.04 |
| **a0014.L** | 0.03 | 0.09 | 0.35 | 1564.76 | 0.73 |
| **a0014.Q** | 0.04 | 0.06 | 0.65 | 1569.01 | 0.52 |
| **a0014.C** | -0.01 | 0.05 | -0.17 | 2634.29 | 0.87 |
| **a0014^4** | -0.02 | 0.04 | -0.49 | 2364.92 | 0.62 |
| **dv_par_edu_birth** | -0.12 | 0.41 | -0.29 | 3842.73 | 0.77 |
| **dv_sc_age_5.L** | 0.15 | 0.08 | 1.86 | 1173.63 | 0.06 |
| **dv_sc_age_5.Q** | -0.03 | 0.04 | -0.94 | 2201.79 | 0.35 |
| **e216aNone** | 0.06 | 0.05 | 1.30 | 1834.48 | 0.19 |
| **dv_cog_abil_5** | -0.01 | 0.26 | -0.04 | 1094.17 | 0.97 |
| **dv_med_51** | 0.04 | 0.05 | 0.84 | 1865.34 | 0.40 |
| **d016a** | 0.09 | 0.04 | 2.00 | 2389.02 | 0.05 |
| **dv_bas_g** | -0.42 | 0.22 | -1.89 | 1252.48 | 0.06 |
| **dv_med_101** | 0.04 | 0.05 | 0.86 | 1517.26 | 0.39 |
| **bmi** | -0.25 | 0.24 | -1.07 | 1603.51 | 0.28 |
| **fclrg90.L** | -0.01 | 0.06 | -0.12 | 2666.32 | 0.91 |
| **fclrg90.Q** | -0.03 | 0.06 | -0.44 | 1667.44 | 0.66 |
| **fclrg90.C** | -0.10 | 0.06 | -1.75 | 2143.98 | 0.08 |
| **fclrg90^4** | 0.01 | 0.05 | 0.21 | 2098.55 | 0.83 |
| **tenure.L** | 0.03 | 0.05 | 0.71 | 1421.27 | 0.48 |
| **tenure.Q** | 0.09 | 0.06 | 1.62 | 2266.23 | 0.10 |
| **divorce1** | 0.14 | 0.05 | 3.17 | 2879.61 | 0.00 |
| **sepmumbcs1** | -0.11 | 0.14 | -0.80 | 1588.73 | 0.42 |
| **prmnh1** | -0.21 | 0.18 | -1.15 | 1125.98 | 0.25 |
| **crowdUp to 1** | 0.03 | 0.06 | 0.47 | 1893.62 | 0.64 |
| **ameniNo occasions** | -0.09 | 0.08 | -1.10 | 2405.81 | 0.27 |
| **brfed.L** | 0.01 | 0.04 | 0.23 | 1618.43 | 0.82 |
| **brfed.Q** | 0.04 | 0.05 | 0.93 | 2232.93 | 0.35 |
| **resmove.L** | 0.00 | 0.04 | 0.03 | 3478.73 | 0.97 |
| **resmove.Q** | -0.06 | 0.03 | -2.21 | 3756.20 | 0.03 |

*Table S25.* Raw output from modified Poisson regression (with log link) of Malaise Inventory score at age 16 (BD4MAL) against median sleep duration (binarised to normal vs abnormal) derived from the van der Berg et al. algorithm at age 46, adjusted for potential confounders (see main text Table 3). The parameter estimates in the "Estimate" column and the associated standard errors have not been exponentiated.

| Term | Estimate | Standard error | Statistic | Degrees of freedom | p-value |
|---|---|---|---|---|---|
| **(Intercept)** | -0.91 | 0.29 | -3.13 | 1704.80 | 0.00 |
| **BD4MAL** | 0.06 | 0.02 | 2.97 | 1686.15 | 0.00 |
| **a0005a** | 0.14 | 0.20 | 0.72 | 1263.25 | 0.47 |
| **a0043b.L** | -0.04 | 0.04 | -1.06 | 3091.69 | 0.29 |
| **a0043b.Q** | 0.03 | 0.06 | 0.49 | 2201.08 | 0.62 |
| **a0043b.C** | 0.06 | 0.04 | 1.43 | 4150.69 | 0.15 |
| **a0043b^4** | 0.02 | 0.05 | 0.42 | 2550.16 | 0.67 |
| **a0043b^5** | 0.05 | 0.07 | 0.71 | 2550.18 | 0.48 |
| **a0195a** | -0.05 | 0.33 | -0.16 | 3913.76 | 0.87 |
| **a0278** | -0.09 | 0.27 | -0.32 | 1606.00 | 0.75 |
| **a0014.L** | 0.08 | 0.07 | 1.11 | 2257.94 | 0.27 |
| **a0014.Q** | 0.01 | 0.06 | 0.22 | 1580.89 | 0.83 |
| **a0014.C** | -0.04 | 0.09 | -0.42 | 1095.19 | 0.67 |
| **a0014^4** | 0.03 | 0.05 | 0.61 | 1593.11 | 0.54 |
| **dv_par_edu_birth** | 0.46 | 0.43 | 1.08 | 1784.44 | 0.28 |
| **dv_sc_age_5.L** | 0.01 | 0.09 | 0.15 | 1015.92 | 0.88 |
| **dv_sc_age_5.Q** | -0.01 | 0.05 | -0.14 | 1406.97 | 0.89 |
| **e216aNone** | -0.13 | 0.05 | -2.32 | 1358.55 | 0.02 |
| **dv_cog_abil_5** | -0.25 | 0.27 | -0.94 | 1007.52 | 0.35 |
| **dv_med_51** | 0.03 | 0.04 | 0.63 | 2676.60 | 0.53 |
| **d016a** | 0.19 | 0.05 | 3.55 | 1448.97 | 0.00 |
| **dv_bas_g** | -0.26 | 0.18 | -1.46 | 1582.39 | 0.14 |
| **dv_med_101** | 0.03 | 0.05 | 0.69 | 1584.49 | 0.49 |
| **bmi** | 0.53 | 0.20 | 2.65 | 1756.56 | 0.01 |
| **fclrg90.L** | -0.12 | 0.09 | -1.35 | 1547.67 | 0.18 |
| **fclrg90.Q** | -0.05 | 0.06 | -0.90 | 1761.91 | 0.37 |
| **fclrg90.C** | 0.03 | 0.07 | 0.39 | 1475.27 | 0.70 |
| **fclrg90^4** | 0.01 | 0.05 | 0.12 | 1756.92 | 0.90 |
| **tenure.L** | 0.04 | 0.04 | 0.88 | 1474.16 | 0.38 |
| **tenure.Q** | -0.04 | 0.06 | -0.78 | 1646.33 | 0.44 |
| **divorce1** | -0.02 | 0.07 | -0.28 | 1479.47 | 0.78 |
| **sepmumbcs1** | 0.21 | 0.09 | 2.21 | 2074.35 | 0.03 |
| **prmnh1** | -0.06 | 0.10 | -0.56 | 2139.37 | 0.58 |
| **crowdUp to 1** | 0.08 | 0.05 | 1.45 | 2001.30 | 0.15 |
| **ameniNo occasions** | -0.17 | 0.09 | -1.78 | 1553.99 | 0.07 |
| **brfed.L** | 0.03 | 0.03 | 0.92 | 2352.25 | 0.36 |
| **brfed.Q** | -0.05 | 0.05 | -0.88 | 1578.80 | 0.38 |
| **resmove.L** | 0.02 | 0.06 | 0.32 | 1450.58 | 0.75 |
| **resmove.Q** | 0.02 | 0.02 | 1.07 | 5179.87 | 0.28 |

*Table S26.* Raw output from modified Poisson regression (with log link) of Malaise Inventory score at age 16 (BD4MAL) against median sleep duration (binarised to normal vs abnormal) derived from the Winkler et al. algorithm at age 46, adjusted for potential confounders (see main text Table 3). The parameter estimates in the “Estimate” column and the associated standard errors have not been exponentiated.

| Term | Estimate | Standard error | Statistic | Degrees of freedom | p-value |
|---|---|---|---|---|---|
| **(Intercept)** | -0.60 | 0.41 | -1.46 | 2542.14 | 0.14 |
| **rd6m_1** | 0.08 | 0.02 | 4.17 | 2570.11 | 0.00 |
| **a0005a** | -0.23 | 0.16 | -1.44 | 8331.65 | 0.15 |
| **a0043b.L** | 0.18 | 0.07 | 2.70 | 2931.25 | 0.01 |
| **a0043b.Q** | 0.10 | 0.09 | 1.14 | 2754.70 | 0.25 |
| **a0043b.C** | 0.03 | 0.08 | 0.39 | 3269.98 | 0.69 |
| **a0043b^4** | -0.04 | 0.10 | -0.39 | 2145.10 | 0.70 |
| **a0043b^5** | -0.06 | 0.12 | -0.50 | 2575.11 | 0.62 |
| **a0195a** | -0.11 | 0.51 | -0.22 | 4269.24 | 0.83 |
| **a0278** | -0.57 | 0.25 | -2.29 | 7840.58 | 0.02 |
| **a0014.L** | 0.13 | 0.10 | 1.34 | 3774.31 | 0.18 |
| **a0014.Q** | 0.00 | 0.06 | -0.06 | 7360.35 | 0.95 |
| **a0014.C** | 0.11 | 0.08 | 1.46 | 3377.64 | 0.14 |
| **a0014^4** | 0.04 | 0.07 | 0.65 | 2760.85 | 0.52 |
| **dv_par_edu_birth** | -1.28 | 1.29 | -0.99 | 2858.96 | 0.32 |
| **dv_sc_age_5.L** | 0.18 | 0.10 | 1.73 | 1660.40 | 0.08 |
| **dv_sc_age_5.Q** | -0.05 | 0.05 | -1.04 | 3552.86 | 0.30 |
| **e216aNone** | -0.02 | 0.06 | -0.39 | 3636.09 | 0.70 |
| **dv_cog_abil_5** | -0.63 | 0.27 | -2.31 | 2068.27 | 0.02 |
| **dv_med_51** | 0.04 | 0.06 | 0.70 | 3885.52 | 0.48 |
| **d016a** | 0.20 | 0.06 | 3.66 | 3980.00 | 0.00 |
| **dv_bas_g** | -0.62 | 0.21 | -2.94 | 2850.45 | 0.00 |
| **dv_med_101** | 0.02 | 0.05 | 0.52 | 6255.26 | 0.60 |
| **bmi** | 0.01 | 0.27 | 0.04 | 3069.53 | 0.96 |
| **fclrg90.L** | 0.04 | 0.09 | 0.39 | 3154.95 | 0.70 |
| **fclrg90.Q** | 0.03 | 0.06 | 0.58 | 6542.85 | 0.56 |
| **fclrg90.C** | -0.07 | 0.07 | -0.99 | 3989.65 | 0.32 |
| **fclrg90^4** | 0.03 | 0.06 | 0.49 | 4917.46 | 0.62 |
| **tenure.L** | 0.12 | 0.05 | 2.22 | 2649.30 | 0.03 |
| **tenure.Q** | -0.06 | 0.06 | -0.93 | 3852.95 | 0.35 |
| **divorce1** | 0.11 | 0.06 | 1.97 | 5662.31 | 0.05 |
| **sepmumbcs1** | 0.07 | 0.11 | 0.64 | 5124.46 | 0.52 |
| **prmnh1** | 0.29 | 0.14 | 2.05 | 1905.33 | 0.04 |
| **crowdUp to 1** | -0.17 | 0.06 | -3.05 | 5269.54 | 0.00 |
| **ameniNo occasions** | -0.03 | 0.11 | -0.26 | 3132.20 | 0.79 |
| **brfed.L** | 0.07 | 0.05 | 1.36 | 3873.00 | 0.17 |
| **brfed.Q** | -0.05 | 0.06 | -0.79 | 3466.28 | 0.43 |
| **resmove.L** | 0.13 | 0.06 | 2.24 | 4028.85 | 0.03 |
| **resmove.Q** | -0.02 | 0.03 | -0.71 | 8968.13 | 0.48 |

*Table S27.* Raw output from modified Poisson regression (with log link) of behavioural and emotional problems at age 16 (rd6m_1) against self-reported average sleep duration (binarised to normal vs abnormal) at age 46, adjusted for potential confounders (see main text Table 3). The parameter estimates in the “Estimate” column and the associated standard errors have not been exponentiated.

| Term | Estimate | Standard error | Statistic | Degrees of freedom | p-value |
|---|---|---|---|---|---|
| **(Intercept)** | -1.10 | 0.46 | -2.37 | 1989.58 | 0.02 |
| **rd6m_1** | 0.06 | 0.03 | 2.08 | 1292.88 | 0.04 |
| **a0005a** | 0.48 | 0.22 | 2.16 | 2493.07 | 0.03 |
| **a0043b.L** | 0.18 | 0.07 | 2.52 | 2785.61 | 0.01 |
| **a0043b.Q** | 0.19 | 0.10 | 1.91 | 2653.27 | 0.06 |
| **a0043b.C** | 0.04 | 0.09 | 0.47 | 2236.15 | 0.64 |
| **a0043b^4** | -0.11 | 0.09 | -1.20 | 3024.88 | 0.23 |
| **a0043b^5** | -0.07 | 0.12 | -0.55 | 2850.41 | 0.58 |
| **a0195a** | -0.08 | 0.66 | -0.12 | 2516.07 | 0.90 |
| **a0278** | 0.29 | 0.27 | 1.06 | 5348.77 | 0.29 |
| **a0014.L** | 0.17 | 0.15 | 1.15 | 1708.56 | 0.25 |
| **a0014.Q** | 0.05 | 0.08 | 0.66 | 2776.44 | 0.51 |
| **a0014.C** | 0.15 | 0.09 | 1.66 | 2429.65 | 0.10 |
| **a0014^4** | 0.09 | 0.06 | 1.49 | 4099.26 | 0.14 |
| **dv_par_edu_birth** | 0.07 | 0.93 | 0.07 | 2617.88 | 0.94 |
| **dv_sc_age_5.L** | 0.08 | 0.12 | 0.66 | 1303.99 | 0.51 |
| **dv_sc_age_5.Q** | 0.01 | 0.05 | 0.26 | 3884.42 | 0.80 |
| **e216aNone** | 0.08 | 0.07 | 1.11 | 2155.63 | 0.27 |
| **dv_cog_abil_5** | -1.43 | 0.44 | -3.27 | 1112.36 | 0.00 |
| **dv_med_51** | -0.02 | 0.10 | -0.19 | 1467.86 | 0.85 |
| **d016a** | 0.11 | 0.07 | 1.57 | 2587.50 | 0.12 |
| **dv_bas_g** | -0.58 | 0.31 | -1.89 | 1612.69 | 0.06 |
| **dv_med_101** | 0.05 | 0.06 | 0.82 | 3024.84 | 0.41 |
| **bmi** | -0.48 | 0.29 | -1.70 | 2950.25 | 0.09 |
| **fclrg90.L** | 0.23 | 0.10 | 2.42 | 2944.87 | 0.02 |
| **fclrg90.Q** | 0.05 | 0.07 | 0.79 | 3735.99 | 0.43 |
| **fclrg90.C** | -0.04 | 0.10 | -0.37 | 1879.16 | 0.71 |
| **fclrg90^4** | -0.02 | 0.08 | -0.21 | 1943.54 | 0.84 |
| **tenure.L** | 0.06 | 0.07 | 0.82 | 1687.45 | 0.41 |
| **tenure.Q** | -0.05 | 0.09 | -0.55 | 2004.30 | 0.58 |
| **divorce1** | 0.05 | 0.07 | 0.71 | 3516.67 | 0.48 |
| **sepmumbcs1** | 0.32 | 0.14 | 2.34 | 2681.48 | 0.02 |
| **prmnh1** | 0.40 | 0.15 | 2.73 | 1714.00 | 0.01 |
| **crowdUp to 1** | -0.11 | 0.07 | -1.69 | 3582.60 | 0.09 |
| **ameniNo occasions** | 0.03 | 0.11 | 0.28 | 4004.58 | 0.78 |
| **brfed.L** | -0.01 | 0.07 | -0.16 | 2133.44 | 0.87 |
| **brfed.Q** | -0.01 | 0.11 | -0.05 | 1360.62 | 0.96 |
| **resmove.L** | 0.21 | 0.09 | 2.39 | 1769.84 | 0.02 |
| **resmove.Q** | 0.06 | 0.04 | 1.40 | 3673.28 | 0.16 |

*Table S28.* Raw output from modified Poisson regression (with log link) of behavioural and emotional problems at age 16 (rd6m_1) against median sleep duration (binarised to normal vs abnormal) derived from the sleep diary at age 46, adjusted for potential confounders (see main text Table 3). The parameter estimates in the “Estimate” column and the associated standard errors have not been exponentiated.

| Term | Estimate | Standard error | Statistic | Degrees of freedom | p-value |
|---|---|---|---|---|---|
| **(Intercept)** | -0.36 | 0.26 | -1.40 | 1948.00 | 0.16 |
| **rd6m_1** | 0.02 | 0.02 | 1.12 | 1392.59 | 0.26 |
| **a0005a** | -0.36 | 0.10 | -3.60 | 5052.11 | 0.00 |
| **a0043b.L** | -0.05 | 0.05 | -0.97 | 1831.54 | 0.33 |
| **a0043b.Q** | -0.02 | 0.06 | -0.37 | 2147.59 | 0.71 |
| **a0043b.C** | 0.00 | 0.06 | 0.00 | 1664.87 | 1.00 |
| **a0043b^4** | -0.06 | 0.04 | -1.31 | 3592.73 | 0.19 |
| **a0043b^5** | -0.01 | 0.07 | -0.19 | 2138.99 | 0.85 |
| **a0195a** | -0.38 | 0.46 | -0.82 | 1679.69 | 0.41 |
| **a0278** | -0.21 | 0.20 | -1.07 | 2437.65 | 0.28 |
| **a0014.L** | 0.02 | 0.07 | 0.22 | 1966.30 | 0.83 |
| **a0014.Q** | 0.03 | 0.05 | 0.58 | 1958.42 | 0.56 |
| **a0014.C** | 0.00 | 0.05 | 0.05 | 2570.78 | 0.96 |
| **a0014^4** | -0.01 | 0.04 | -0.26 | 1785.71 | 0.79 |
| **dv_par_edu_birth** | -0.39 | 0.46 | -0.84 | 2695.45 | 0.40 |
| **dv_sc_age_5.L** | -0.02 | 0.06 | -0.27 | 1510.49 | 0.78 |
| **dv_sc_age_5.Q** | 0.00 | 0.03 | -0.14 | 2894.17 | 0.89 |
| **e216aNone** | -0.01 | 0.05 | -0.14 | 1549.63 | 0.89 |
| **dv_cog_abil_5** | 0.04 | 0.28 | 0.16 | 887.70 | 0.87 |
| **dv_med_51** | 0.00 | 0.04 | 0.13 | 3064.80 | 0.90 |
| **d016a** | 0.05 | 0.03 | 1.69 | 4138.99 | 0.09 |
| **dv_bas_g** | -0.23 | 0.16 | -1.45 | 1825.92 | 0.15 |
| **dv_med_101** | 0.01 | 0.03 | 0.33 | 2824.65 | 0.74 |
| **bmi** | 0.19 | 0.17 | 1.12 | 2135.11 | 0.26 |
| **fclrg90.L** | 0.03 | 0.06 | 0.46 | 2471.69 | 0.64 |
| **fclrg90.Q** | 0.05 | 0.04 | 1.09 | 2462.20 | 0.28 |
| **fclrg90.C** | 0.06 | 0.04 | 1.53 | 4046.02 | 0.13 |
| **fclrg90^4** | 0.01 | 0.05 | 0.29 | 1784.38 | 0.77 |
| **tenure.L** | 0.01 | 0.03 | 0.46 | 2315.82 | 0.65 |
| **tenure.Q** | -0.03 | 0.05 | -0.70 | 2284.63 | 0.48 |
| **divorce1** | 0.03 | 0.04 | 0.75 | 3570.53 | 0.45 |
| **sepmumbcs1** | -0.07 | 0.09 | -0.77 | 2911.09 | 0.44 |
| **prmnh1** | 0.18 | 0.10 | 1.91 | 1746.53 | 0.06 |
| **crowdUp to 1** | -0.05 | 0.05 | -0.90 | 1888.51 | 0.37 |
| **ameniNo occasions** | -0.07 | 0.07 | -1.01 | 2197.44 | 0.31 |
| **brfed.L** | 0.03 | 0.03 | 0.79 | 2093.50 | 0.43 |
| **brfed.Q** | -0.01 | 0.03 | -0.43 | 3443.69 | 0.67 |
| **resmove.L** | -0.03 | 0.04 | -0.71 | 2734.58 | 0.48 |
| **resmove.Q** | 0.06 | 0.02 | 2.52 | 4360.14 | 0.01 |

*Table S29.* Raw output from modified Poisson regression (with log link) of behavioural and emotional problems at age 16 (rd6m_1) against median sleep duration (binarised to normal vs abnormal) derived from the activPAL algorithm at age 46, adjusted for potential confounders (see main text Table 3). The parameter estimates in the “Estimate” column and the associated standard errors have not been exponentiated.

| Term | Estimate | Standard error | Statistic | Degrees of freedom | p-value |
|---|---|---|---|---|---|
| **(Intercept)** | -0.88 | 0.34 | -2.58 | 1509.65 | 0.01 |
| **rd6m_1** | -0.04 | 0.03 | -1.40 | 1460.21 | 0.16 |
| **a0005a** | -0.01 | 0.17 | -0.08 | 1788.47 | 0.94 |
| **a0043b.L** | 0.01 | 0.06 | 0.08 | 1592.54 | 0.93 |
| **a0043b.Q** | -0.06 | 0.07 | -0.90 | 1770.43 | 0.37 |
| **a0043b.C** | -0.18 | 0.06 | -2.80 | 1732.05 | 0.01 |
| **a0043b^4** | 0.10 | 0.06 | 1.61 | 2070.86 | 0.11 |
| **a0043b^5** | 0.04 | 0.08 | 0.47 | 2144.04 | 0.64 |
| **a0195a** | -0.25 | 0.47 | -0.53 | 1963.59 | 0.60 |
| **a0278** | 0.36 | 0.18 | 1.98 | 3884.97 | 0.05 |
| **a0014.L** | 0.03 | 0.09 | 0.34 | 1551.11 | 0.73 |
| **a0014.Q** | 0.04 | 0.07 | 0.65 | 1553.81 | 0.51 |
| **a0014.C** | -0.01 | 0.05 | -0.14 | 2677.45 | 0.88 |
| **a0014^4** | -0.02 | 0.04 | -0.49 | 2400.32 | 0.63 |
| **dv_par_edu_birth** | -0.12 | 0.41 | -0.29 | 3921.66 | 0.77 |
| **dv_sc_age_5.L** | 0.16 | 0.08 | 1.85 | 1134.12 | 0.07 |
| **dv_sc_age_5.Q** | -0.03 | 0.04 | -0.95 | 2216.80 | 0.34 |
| **e216aNone** | 0.06 | 0.05 | 1.28 | 1871.77 | 0.20 |
| **dv_cog_abil_5** | -0.02 | 0.26 | -0.06 | 1103.67 | 0.95 |
| **dv_med_51** | 0.05 | 0.05 | 0.89 | 1895.23 | 0.37 |
| **d016a** | 0.09 | 0.04 | 2.08 | 2337.35 | 0.04 |
| **dv_bas_g** | -0.43 | 0.22 | -1.94 | 1257.88 | 0.05 |
| **dv_med_101** | 0.05 | 0.05 | 0.91 | 1573.39 | 0.36 |
| **bmi** | -0.25 | 0.24 | -1.04 | 1566.27 | 0.30 |
| **fclrg90.L** | -0.01 | 0.06 | -0.17 | 2595.25 | 0.87 |
| **fclrg90.Q** | -0.03 | 0.06 | -0.47 | 1678.47 | 0.64 |
| **fclrg90.C** | -0.10 | 0.06 | -1.78 | 2169.52 | 0.08 |
| **fclrg90^4** | 0.01 | 0.05 | 0.19 | 2100.02 | 0.85 |
| **tenure.L** | 0.03 | 0.05 | 0.70 | 1397.94 | 0.48 |
| **tenure.Q** | 0.09 | 0.05 | 1.68 | 2343.02 | 0.09 |
| **divorce1** | 0.15 | 0.05 | 3.24 | 2899.34 | 0.00 |
| **sepmumbcs1** | -0.10 | 0.14 | -0.72 | 1567.61 | 0.47 |
| **prmnh1** | -0.21 | 0.18 | -1.17 | 1152.16 | 0.24 |
| **crowdUp to 1** | 0.03 | 0.06 | 0.45 | 1833.26 | 0.66 |
| **ameniNo occasions** | -0.09 | 0.08 | -1.18 | 2420.09 | 0.24 |
| **brfed.L** | 0.01 | 0.04 | 0.25 | 1621.47 | 0.80 |
| **brfed.Q** | 0.04 | 0.05 | 0.93 | 2219.02 | 0.35 |
| **resmove.L** | 0.00 | 0.04 | 0.02 | 3426.48 | 0.98 |
| **resmove.Q** | -0.06 | 0.03 | -2.22 | 3747.41 | 0.03 |

*Table S30.* Raw output from modified Poisson regression (with log link) of behavioural and emotional problems at age 16 (rd6m_1) against median sleep duration (binarised to normal vs abnormal) derived from the van der Berg et al. algorithm at age 46, adjusted for potential confounders (see main text Table 3). The parameter estimates in the "Estimate" column and the associated standard errors have not been exponentiated.

| Term | Estimate | Standard error | Statistic | Degrees of freedom | p-value |
|---|---|---|---|---|---|
| **(Intercept)** | -0.88 | 0.29 | -2.99 | 1669.75 | 0.00 |
| **rd6m_1** | 0.01 | 0.02 | 0.78 | 1789.73 | 0.43 |
| **a0005a** | 0.14 | 0.20 | 0.68 | 1240.71 | 0.50 |
| **a0043b.L** | -0.04 | 0.04 | -1.06 | 3017.63 | 0.29 |
| **a0043b.Q** | 0.02 | 0.06 | 0.42 | 2212.57 | 0.68 |
| **a0043b.C** | 0.06 | 0.04 | 1.56 | 4302.55 | 0.12 |
| **a0043b^4** | 0.02 | 0.05 | 0.47 | 2619.99 | 0.64 |
| **a0043b^5** | 0.04 | 0.07 | 0.58 | 2484.28 | 0.56 |
| **a0195a** | -0.06 | 0.33 | -0.17 | 3967.41 | 0.86 |
| **a0278** | -0.11 | 0.27 | -0.39 | 1607.47 | 0.69 |
| **a0014.L** | 0.08 | 0.07 | 1.11 | 2309.70 | 0.27 |
| **a0014.Q** | 0.01 | 0.06 | 0.20 | 1591.82 | 0.84 |
| **a0014.C** | -0.04 | 0.09 | -0.40 | 1087.47 | 0.69 |
| **a0014^4** | 0.03 | 0.05 | 0.64 | 1564.22 | 0.52 |
| **dv_par_edu_birth** | 0.44 | 0.44 | 1.00 | 1748.01 | 0.32 |
| **dv_sc_age_5.L** | 0.02 | 0.09 | 0.23 | 1005.11 | 0.82 |
| **dv_sc_age_5.Q** | -0.01 | 0.05 | -0.15 | 1417.55 | 0.88 |
| **e216aNone** | -0.13 | 0.05 | -2.39 | 1386.75 | 0.02 |
| **dv_cog_abil_5** | -0.26 | 0.27 | -0.98 | 1011.73 | 0.32 |
| **dv_med_51** | 0.03 | 0.04 | 0.75 | 2781.47 | 0.45 |
| **d016a** | 0.19 | 0.05 | 3.56 | 1447.65 | 0.00 |
| **dv_bas_g** | -0.28 | 0.19 | -1.50 | 1514.46 | 0.13 |
| **dv_med_101** | 0.04 | 0.05 | 0.77 | 1571.96 | 0.44 |
| **bmi** | 0.53 | 0.20 | 2.64 | 1711.24 | 0.01 |
| **fclrg90.L** | -0.12 | 0.08 | -1.44 | 1555.06 | 0.15 |
| **fclrg90.Q** | -0.05 | 0.06 | -0.88 | 1807.17 | 0.38 |
| **fclrg90.C** | 0.03 | 0.07 | 0.37 | 1483.99 | 0.71 |
| **fclrg90^4** | 0.01 | 0.05 | 0.13 | 1753.34 | 0.90 |
| **tenure.L** | 0.04 | 0.04 | 0.88 | 1504.33 | 0.38 |
| **tenure.Q** | -0.04 | 0.06 | -0.76 | 1613.68 | 0.45 |
| **divorce1** | -0.01 | 0.06 | -0.13 | 1537.71 | 0.89 |
| **sepmumbcs1** | 0.20 | 0.10 | 2.09 | 2030.85 | 0.04 |
| **prmnh1** | -0.03 | 0.11 | -0.30 | 1989.66 | 0.76 |
| **crowdUp to 1** | 0.07 | 0.05 | 1.32 | 2067.39 | 0.19 |
| **ameniNo occasions** | -0.16 | 0.10 | -1.62 | 1468.55 | 0.10 |
| **brfed.L** | 0.03 | 0.04 | 0.87 | 2278.45 | 0.38 |
| **brfed.Q** | -0.05 | 0.05 | -0.91 | 1612.03 | 0.36 |
| **resmove.L** | 0.02 | 0.06 | 0.33 | 1424.92 | 0.74 |
| **resmove.Q** | 0.02 | 0.02 | 1.07 | 5290.50 | 0.29 |

*Table S31.* Raw output from modified Poisson regression (with log link) of behavioural and emotional problems at age 16 (rd6m_1) against median sleep duration (binarised to normal vs abnormal) derived from the Winkler et al. algorithm at age 46, adjusted for potential confounders (see main text Table 3). The parameter estimates in the “Estimate” column and the associated standard errors have not been exponentiated.

| Term | Estimate | Standard error | Statistic | Degrees of freedom | p-value |
|---|---|---|---|---|---|
| **(Intercept)** | -0.62 | 0.42 | -1.46 | 2427.42 | 0.14 |
| **intbcsz** | 0.07 | 0.03 | 2.36 | 2075.85 | 0.02 |
| **a0005a** | -0.21 | 0.16 | -1.31 | 7730.55 | 0.19 |
| **a0043b.L** | 0.17 | 0.07 | 2.56 | 3055.43 | 0.01 |
| **a0043b.Q** | 0.10 | 0.09 | 1.11 | 2671.35 | 0.27 |
| **a0043b.C** | 0.03 | 0.08 | 0.41 | 3216.16 | 0.68 |
| **a0043b^4** | -0.04 | 0.10 | -0.40 | 2123.27 | 0.69 |
| **a0043b^5** | -0.06 | 0.12 | -0.52 | 2673.37 | 0.60 |
| **a0195a** | -0.10 | 0.52 | -0.20 | 4060.17 | 0.85 |
| **a0278** | -0.56 | 0.25 | -2.28 | 8015.94 | 0.02 |
| **a0014.L** | 0.13 | 0.10 | 1.35 | 3715.22 | 0.18 |
| **a0014.Q** | 0.00 | 0.06 | -0.01 | 7398.83 | 0.99 |
| **a0014.C** | 0.12 | 0.08 | 1.54 | 3505.96 | 0.12 |
| **a0014^4** | 0.04 | 0.07 | 0.65 | 2724.10 | 0.51 |
| **dv_par_edu_birth** | -1.25 | 1.26 | -0.99 | 2950.05 | 0.32 |
| **dv_sc_age_5.L** | 0.18 | 0.10 | 1.84 | 1728.31 | 0.07 |
| **dv_sc_age_5.Q** | -0.05 | 0.05 | -1.05 | 3531.76 | 0.29 |
| **e216aNone** | -0.02 | 0.06 | -0.35 | 3308.82 | 0.72 |
| **dv_cog_abil_5** | -0.64 | 0.28 | -2.25 | 1953.35 | 0.02 |
| **dv_med_51** | 0.04 | 0.06 | 0.61 | 3567.68 | 0.54 |
| **d016a** | 0.21 | 0.05 | 3.87 | 4096.24 | 0.00 |
| **dv_bas_g** | -0.61 | 0.21 | -2.85 | 2772.34 | 0.00 |
| **dv_med_101** | 0.02 | 0.05 | 0.37 | 6540.44 | 0.71 |
| **bmi** | 0.03 | 0.27 | 0.11 | 2876.19 | 0.92 |
| **fclrg90.L** | 0.02 | 0.09 | 0.24 | 3366.28 | 0.81 |
| **fclrg90.Q** | 0.03 | 0.06 | 0.48 | 7058.57 | 0.63 |
| **fclrg90.C** | -0.08 | 0.08 | -1.05 | 3830.15 | 0.30 |
| **fclrg90^4** | 0.03 | 0.06 | 0.45 | 4858.81 | 0.65 |
| **tenure.L** | 0.12 | 0.05 | 2.23 | 2695.98 | 0.03 |
| **tenure.Q** | -0.06 | 0.07 | -0.90 | 3660.57 | 0.37 |
| **divorce1** | 0.11 | 0.06 | 1.81 | 5525.61 | 0.07 |
| **sepmumbcs1** | 0.11 | 0.11 | 0.96 | 5412.99 | 0.33 |
| **prmnh1** | 0.25 | 0.14 | 1.70 | 1856.58 | 0.09 |
| **crowdUp to 1** | -0.17 | 0.06 | -3.02 | 5388.08 | 0.00 |
| **ameniNo occasions** | -0.04 | 0.11 | -0.39 | 2832.00 | 0.70 |
| **brfed.L** | 0.07 | 0.05 | 1.41 | 3756.06 | 0.16 |
| **brfed.Q** | -0.04 | 0.06 | -0.69 | 3354.20 | 0.49 |
| **resmove.L** | 0.13 | 0.06 | 2.17 | 3979.35 | 0.03 |
| **resmove.Q** | -0.03 | 0.03 | -0.75 | 9504.53 | 0.46 |

*Table S32.* Raw output from modified Poisson regression (with log link) of internalising behaviour at age 16 (intbcsz) against self-reported average sleep duration (binarised to normal vs abnormal) at age 46, adjusted for potential confounders (see main text Table 3). The parameter estimates in the “Estimate” column and the associated standard errors have not been exponentiated.

| Term | Estimate | Standard error | Statistic | Degrees of freedom | p-value |
|---|---|---|---|---|---|
| **(Intercept)** | -1.12 | 0.46 | -2.43 | 2012.62 | 0.02 |
| **intbcsz** | 0.06 | 0.02 | 2.97 | 5359.54 | 0.00 |
| **a0005a** | 0.50 | 0.22 | 2.22 | 2455.43 | 0.03 |
| **a0043b.L** | 0.17 | 0.07 | 2.35 | 2712.63 | 0.02 |
| **a0043b.Q** | 0.19 | 0.10 | 1.87 | 2582.91 | 0.06 |
| **a0043b.C** | 0.04 | 0.09 | 0.47 | 2242.99 | 0.64 |
| **a0043b^4** | -0.11 | 0.09 | -1.20 | 2947.69 | 0.23 |
| **a0043b^5** | -0.07 | 0.12 | -0.57 | 3006.99 | 0.57 |
| **a0195a** | -0.07 | 0.66 | -0.10 | 2509.24 | 0.92 |
| **a0278** | 0.30 | 0.27 | 1.08 | 5342.52 | 0.28 |
| **a0014.L** | 0.17 | 0.15 | 1.14 | 1672.77 | 0.26 |
| **a0014.Q** | 0.05 | 0.08 | 0.69 | 2787.34 | 0.49 |
| **a0014.C** | 0.15 | 0.09 | 1.69 | 2452.54 | 0.09 |
| **a0014^4** | 0.09 | 0.06 | 1.48 | 3915.85 | 0.14 |
| **dv_par_edu_birth** | 0.09 | 0.92 | 0.09 | 2692.45 | 0.92 |
| **dv_sc_age_5.L** | 0.09 | 0.13 | 0.68 | 1254.11 | 0.50 |
| **dv_sc_age_5.Q** | 0.01 | 0.05 | 0.25 | 3756.50 | 0.80 |
| **e216aNone** | 0.08 | 0.07 | 1.13 | 2139.69 | 0.26 |
| **dv_cog_abil_5** | -1.43 | 0.44 | -3.27 | 1114.56 | 0.00 |
| **dv_med_51** | -0.02 | 0.10 | -0.22 | 1497.78 | 0.82 |
| **d016a** | 0.11 | 0.07 | 1.72 | 2762.44 | 0.09 |
| **dv_bas_g** | -0.57 | 0.31 | -1.82 | 1581.74 | 0.07 |
| **dv_med_101** | 0.04 | 0.06 | 0.69 | 2899.96 | 0.49 |
| **bmi** | -0.47 | 0.29 | -1.63 | 2903.07 | 0.10 |
| **fclrg90.L** | 0.22 | 0.10 | 2.30 | 2914.48 | 0.02 |
| **fclrg90.Q** | 0.05 | 0.07 | 0.70 | 3598.02 | 0.49 |
| **fclrg90.C** | -0.04 | 0.10 | -0.40 | 1896.76 | 0.69 |
| **fclrg90^4** | -0.02 | 0.08 | -0.23 | 1984.10 | 0.82 |
| **tenure.L** | 0.06 | 0.07 | 0.82 | 1684.90 | 0.41 |
| **tenure.Q** | -0.05 | 0.09 | -0.53 | 1970.24 | 0.60 |
| **divorce1** | 0.04 | 0.07 | 0.60 | 3382.85 | 0.55 |
| **sepmumbcs1** | 0.34 | 0.13 | 2.59 | 2851.31 | 0.01 |
| **prmnh1** | 0.36 | 0.15 | 2.39 | 1677.06 | 0.02 |
| **crowdUp to 1** | -0.11 | 0.07 | -1.65 | 3506.37 | 0.10 |
| **ameniNo occasions** | 0.02 | 0.11 | 0.17 | 4051.50 | 0.87 |
| **brfed.L** | -0.01 | 0.06 | -0.13 | 2169.66 | 0.90 |
| **brfed.Q** | 0.00 | 0.11 | -0.01 | 1324.63 | 0.99 |
| **resmove.L** | 0.20 | 0.09 | 2.35 | 1760.75 | 0.02 |
| **resmove.Q** | 0.06 | 0.04 | 1.38 | 3662.39 | 0.17 |

*Table S33.* Raw output from modified Poisson regression (with log link) of internalising behaviour at age 16 (intbcsz) against median sleep duration (binarised to normal vs abnormal) derived from the sleep diary at age 46, adjusted for potential confounders (see main text Table 3). The parameter estimates in the “Estimate” column and the associated standard errors have not been exponentiated.

| Term | Estimate | Standard error | Statistic | Degrees of freedom | p-value |
|---|---|---|---|---|---|
| **(Intercept)** | -0.36 | 0.26 | -1.41 | 1944.57 | 0.16 |
| **intbcsz** | 0.02 | 0.02 | 1.18 | 2058.03 | 0.24 |
| **a0005a** | -0.35 | 0.10 | -3.50 | 4803.89 | 0.00 |
| **a0043b.L** | -0.05 | 0.05 | -1.03 | 1840.42 | 0.30 |
| **a0043b.Q** | -0.02 | 0.06 | -0.38 | 2165.32 | 0.71 |
| **a0043b.C** | 0.00 | 0.06 | 0.01 | 1672.71 | 0.99 |
| **a0043b^4** | -0.06 | 0.04 | -1.31 | 3561.61 | 0.19 |
| **a0043b^5** | -0.01 | 0.07 | -0.19 | 2142.63 | 0.85 |
| **a0195a** | -0.38 | 0.46 | -0.83 | 1690.35 | 0.41 |
| **a0278** | -0.21 | 0.20 | -1.04 | 2382.68 | 0.30 |
| **a0014.L** | 0.02 | 0.07 | 0.21 | 1972.82 | 0.83 |
| **a0014.Q** | 0.03 | 0.05 | 0.59 | 1981.61 | 0.55 |
| **a0014.C** | 0.00 | 0.05 | 0.06 | 2529.94 | 0.95 |
| **a0014^4** | -0.01 | 0.04 | -0.25 | 1791.70 | 0.80 |
| **dv_par_edu_birth** | -0.39 | 0.46 | -0.84 | 2735.09 | 0.40 |
| **dv_sc_age_5.L** | -0.02 | 0.06 | -0.24 | 1504.61 | 0.81 |
| **dv_sc_age_5.Q** | 0.00 | 0.03 | -0.15 | 2917.00 | 0.88 |
| **e216aNone** | -0.01 | 0.05 | -0.13 | 1558.50 | 0.90 |
| **dv_cog_abil_5** | 0.04 | 0.28 | 0.16 | 879.45 | 0.88 |
| **dv_med_51** | 0.00 | 0.04 | 0.09 | 3167.47 | 0.93 |
| **d016a** | 0.06 | 0.03 | 1.66 | 3602.01 | 0.10 |
| **dv_bas_g** | -0.22 | 0.16 | -1.39 | 1765.17 | 0.17 |
| **dv_med_101** | 0.01 | 0.03 | 0.25 | 2675.35 | 0.80 |
| **bmi** | 0.20 | 0.17 | 1.14 | 2144.12 | 0.25 |
| **fclrg90.L** | 0.02 | 0.06 | 0.40 | 2472.90 | 0.69 |
| **fclrg90.Q** | 0.05 | 0.04 | 1.06 | 2455.66 | 0.29 |
| **fclrg90.C** | 0.06 | 0.04 | 1.50 | 4133.14 | 0.13 |
| **fclrg90^4** | 0.01 | 0.05 | 0.27 | 1752.99 | 0.79 |
| **tenure.L** | 0.01 | 0.03 | 0.46 | 2369.25 | 0.65 |
| **tenure.Q** | -0.03 | 0.05 | -0.69 | 2274.87 | 0.49 |
| **divorce1** | 0.03 | 0.04 | 0.70 | 3654.67 | 0.48 |
| **sepmumbcs1** | -0.06 | 0.09 | -0.70 | 3028.44 | 0.48 |
| **prmnh1** | 0.17 | 0.09 | 1.86 | 1874.80 | 0.06 |
| **crowdUp to 1** | -0.05 | 0.05 | -0.88 | 1883.25 | 0.38 |
| **ameniNo occasions** | -0.08 | 0.07 | -1.04 | 2158.48 | 0.30 |
| **brfed.L** | 0.03 | 0.03 | 0.81 | 2078.67 | 0.42 |
| **brfed.Q** | -0.01 | 0.03 | -0.39 | 3477.39 | 0.70 |
| **resmove.L** | -0.03 | 0.04 | -0.73 | 2744.46 | 0.47 |
| **resmove.Q** | 0.06 | 0.02 | 2.51 | 4355.78 | 0.01 |

*Table S34.* Raw output from modified Poisson regression (with log link) of internalising behaviour at age 16 (intbcsz) against median sleep duration (binarised to normal vs abnormal) derived from the activPAL algorithm at age 46, adjusted for potential confounders (see main text Table 3). The parameter estimates in the “Estimate” column and the associated standard errors have not been exponentiated.

| Term | Estimate | Standard error | Statistic | Degrees of freedom | p-value |
|---|---|---|---|---|---|
| **(Intercept)** | -0.89 | 0.35 | -2.56 | 1460.51 | 0.01 |
| **intbcsz** | 0.01 | 0.03 | 0.39 | 1239.14 | 0.70 |
| **a0005a** | -0.01 | 0.17 | -0.07 | 1788.82 | 0.94 |
| **a0043b.L** | 0.01 | 0.06 | 0.11 | 1596.81 | 0.91 |
| **a0043b.Q** | -0.06 | 0.07 | -0.92 | 1795.19 | 0.36 |
| **a0043b.C** | -0.18 | 0.07 | -2.79 | 1728.62 | 0.01 |
| **a0043b^4** | 0.10 | 0.06 | 1.60 | 2048.95 | 0.11 |
| **a0043b^5** | 0.03 | 0.08 | 0.45 | 2123.95 | 0.65 |
| **a0195a** | -0.26 | 0.47 | -0.55 | 1994.30 | 0.58 |
| **a0278** | 0.36 | 0.18 | 2.00 | 3808.65 | 0.05 |
| **a0014.L** | 0.03 | 0.09 | 0.33 | 1544.89 | 0.74 |
| **a0014.Q** | 0.04 | 0.06 | 0.66 | 1568.70 | 0.51 |
| **a0014.C** | -0.01 | 0.05 | -0.16 | 2602.03 | 0.87 |
| **a0014^4** | -0.02 | 0.04 | -0.49 | 2395.42 | 0.63 |
| **dv_par_edu_birth** | -0.12 | 0.41 | -0.29 | 3829.98 | 0.77 |
| **dv_sc_age_5.L** | 0.15 | 0.08 | 1.86 | 1157.80 | 0.06 |
| **dv_sc_age_5.Q** | -0.03 | 0.04 | -0.94 | 2190.06 | 0.35 |
| **e216aNone** | 0.06 | 0.05 | 1.30 | 1804.11 | 0.19 |
| **dv_cog_abil_5** | -0.01 | 0.26 | -0.04 | 1093.85 | 0.97 |
| **dv_med_51** | 0.04 | 0.05 | 0.82 | 1839.16 | 0.41 |
| **d016a** | 0.09 | 0.04 | 1.95 | 2338.77 | 0.05 |
| **dv_bas_g** | -0.41 | 0.22 | -1.89 | 1263.61 | 0.06 |
| **dv_med_101** | 0.04 | 0.05 | 0.88 | 1637.90 | 0.38 |
| **bmi** | -0.25 | 0.24 | -1.04 | 1564.35 | 0.30 |
| **fclrg90.L** | -0.01 | 0.07 | -0.14 | 2528.92 | 0.89 |
| **fclrg90.Q** | -0.03 | 0.06 | -0.44 | 1662.06 | 0.66 |
| **fclrg90.C** | -0.10 | 0.06 | -1.75 | 2120.46 | 0.08 |
| **fclrg90^4** | 0.01 | 0.05 | 0.21 | 2112.37 | 0.83 |
| **tenure.L** | 0.03 | 0.05 | 0.70 | 1413.32 | 0.48 |
| **tenure.Q** | 0.09 | 0.05 | 1.64 | 2283.91 | 0.10 |
| **divorce1** | 0.14 | 0.05 | 2.99 | 2569.77 | 0.00 |
| **sepmumbcs1** | -0.11 | 0.14 | -0.80 | 1567.02 | 0.43 |
| **prmnh1** | -0.22 | 0.18 | -1.17 | 1131.87 | 0.24 |
| **crowdUp to 1** | 0.03 | 0.06 | 0.47 | 1864.89 | 0.64 |
| **ameniNo occasions** | -0.08 | 0.08 | -1.08 | 2366.38 | 0.28 |
| **brfed.L** | 0.01 | 0.04 | 0.23 | 1613.16 | 0.82 |
| **brfed.Q** | 0.04 | 0.05 | 0.94 | 2261.24 | 0.35 |
| **resmove.L** | 0.00 | 0.04 | 0.04 | 3467.49 | 0.97 |
| **resmove.Q** | -0.06 | 0.03 | -2.22 | 3830.31 | 0.03 |

*Table S35.* Raw output from modified Poisson regression (with log link) of internalising behaviour at age 16 (intbcsz) against median sleep duration (binarised to normal vs abnormal) derived from the van der Berg et al. algorithm at age 46, adjusted for potential confounders (see main text Table 3). The parameter estimates in the “Estimate” column and the associated standard errors have not been exponentiated.

| Term | Estimate | Standard error | Statistic | Degrees of freedom | p-value |
|---|---|---|---|---|---|
| **(Intercept)** | -0.89 | 0.29 | -3.08 | 1720.72 | 0.00 |
| **intbcsz** | 0.02 | 0.02 | 1.00 | 1376.36 | 0.32 |
| **a0005a** | 0.14 | 0.20 | 0.71 | 1241.74 | 0.48 |
| **a0043b.L** | -0.04 | 0.04 | -1.11 | 2945.66 | 0.27 |
| **a0043b.Q** | 0.02 | 0.06 | 0.39 | 2157.74 | 0.70 |
| **a0043b.C** | 0.06 | 0.04 | 1.57 | 4310.07 | 0.12 |
| **a0043b^4** | 0.02 | 0.05 | 0.47 | 2663.53 | 0.64 |
| **a0043b^5** | 0.04 | 0.07 | 0.57 | 2492.72 | 0.57 |
| **a0195a** | -0.06 | 0.33 | -0.19 | 3940.98 | 0.85 |
| **a0278** | -0.10 | 0.27 | -0.37 | 1590.73 | 0.71 |
| **a0014.L** | 0.08 | 0.07 | 1.10 | 2310.74 | 0.27 |
| **a0014.Q** | 0.01 | 0.06 | 0.21 | 1580.27 | 0.84 |
| **a0014.C** | -0.03 | 0.09 | -0.40 | 1090.92 | 0.69 |
| **a0014^4** | 0.03 | 0.05 | 0.65 | 1572.70 | 0.51 |
| **dv_par_edu_birth** | 0.44 | 0.43 | 1.02 | 1777.58 | 0.31 |
| **dv_sc_age_5.L** | 0.02 | 0.09 | 0.25 | 1008.39 | 0.80 |
| **dv_sc_age_5.Q** | -0.01 | 0.05 | -0.17 | 1417.64 | 0.87 |
| **e216aNone** | -0.13 | 0.05 | -2.39 | 1396.08 | 0.02 |
| **dv_cog_abil_5** | -0.26 | 0.27 | -0.98 | 1013.21 | 0.33 |
| **dv_med_51** | 0.03 | 0.04 | 0.67 | 2580.96 | 0.50 |
| **d016a** | 0.19 | 0.05 | 3.64 | 1497.38 | 0.00 |
| **dv_bas_g** | -0.27 | 0.18 | -1.50 | 1586.10 | 0.13 |
| **dv_med_101** | 0.03 | 0.05 | 0.72 | 1635.86 | 0.47 |
| **bmi** | 0.54 | 0.20 | 2.64 | 1699.38 | 0.01 |
| **fclrg90.L** | -0.13 | 0.08 | -1.48 | 1558.48 | 0.14 |
| **fclrg90.Q** | -0.05 | 0.06 | -0.90 | 1802.40 | 0.37 |
| **fclrg90.C** | 0.02 | 0.07 | 0.35 | 1473.36 | 0.72 |
| **fclrg90^4** | 0.01 | 0.05 | 0.13 | 1765.98 | 0.90 |
| **tenure.L** | 0.04 | 0.04 | 0.88 | 1507.23 | 0.38 |
| **tenure.Q** | -0.04 | 0.06 | -0.76 | 1630.87 | 0.45 |
| **divorce1** | -0.01 | 0.07 | -0.18 | 1431.22 | 0.86 |
| **sepmumbcs1** | 0.21 | 0.10 | 2.13 | 2017.30 | 0.03 |
| **prmnh1** | -0.05 | 0.11 | -0.41 | 1928.68 | 0.68 |
| **crowdUp to 1** | 0.07 | 0.05 | 1.34 | 2061.00 | 0.18 |
| **ameniNo occasions** | -0.16 | 0.10 | -1.67 | 1506.01 | 0.10 |
| **brfed.L** | 0.03 | 0.03 | 0.89 | 2285.21 | 0.37 |
| **brfed.Q** | -0.05 | 0.05 | -0.89 | 1600.92 | 0.38 |
| **resmove.L** | 0.02 | 0.06 | 0.33 | 1434.88 | 0.74 |
| **resmove.Q** | 0.02 | 0.02 | 1.04 | 5301.10 | 0.30 |

*Table S36.* Raw output from modified Poisson regression (with log link) of internalising behaviour at age 16 (intbcsz) against median sleep duration (binarised to normal vs abnormal) derived from the Winkler et al. algorithm at age 46, adjusted for potential confounders (see main text Table 3). The parameter estimates in the "Estimate" column and the associated standard errors have not been exponentiated.

| Term | Estimate | Standard error | Statistic | Degrees of freedom | p-value |
|---|---|---|---|---|---|
| **(Intercept)** | -0.72 | 0.41 | -1.76 | 2637.60 | 0.08 |
| **extbcsz** | 0.11 | 0.03 | 3.42 | 1445.80 | 0.00 |
| **a0005a** | -0.18 | 0.15 | -1.18 | 9222.01 | 0.24 |
| **a0043b.L** | 0.17 | 0.07 | 2.52 | 2961.01 | 0.01 |
| **a0043b.Q** | 0.10 | 0.09 | 1.10 | 2742.81 | 0.27 |
| **a0043b.C** | 0.03 | 0.08 | 0.43 | 3125.35 | 0.67 |
| **a0043b^4** | -0.04 | 0.10 | -0.34 | 2112.27 | 0.73 |
| **a0043b^5** | -0.07 | 0.12 | -0.56 | 2556.20 | 0.58 |
| **a0195a** | -0.04 | 0.52 | -0.08 | 3932.92 | 0.94 |
| **a0278** | -0.58 | 0.25 | -2.34 | 7702.67 | 0.02 |
| **a0014.L** | 0.13 | 0.10 | 1.34 | 3894.36 | 0.18 |
| **a0014.Q** | -0.01 | 0.06 | -0.14 | 7576.20 | 0.89 |
| **a0014.C** | 0.12 | 0.08 | 1.56 | 3590.03 | 0.12 |
| **a0014^4** | 0.04 | 0.07 | 0.61 | 2743.20 | 0.54 |
| **dv_par_edu_birth** | -1.20 | 1.24 | -0.97 | 3036.29 | 0.33 |
| **dv_sc_age_5.L** | 0.17 | 0.10 | 1.75 | 1760.65 | 0.08 |
| **dv_sc_age_5.Q** | -0.05 | 0.05 | -1.05 | 3559.16 | 0.30 |
| **e216aNone** | -0.01 | 0.06 | -0.23 | 3470.46 | 0.81 |
| **dv_cog_abil_5** | -0.60 | 0.28 | -2.09 | 1930.70 | 0.04 |
| **dv_med_51** | 0.03 | 0.06 | 0.52 | 3967.80 | 0.60 |
| **d016a** | 0.20 | 0.06 | 3.54 | 3932.83 | 0.00 |
| **dv_bas_g** | -0.57 | 0.22 | -2.63 | 2716.70 | 0.01 |
| **dv_med_101** | 0.02 | 0.05 | 0.43 | 6724.20 | 0.67 |
| **bmi** | 0.04 | 0.26 | 0.14 | 3111.04 | 0.89 |
| **fclrg90.L** | 0.02 | 0.09 | 0.26 | 3137.29 | 0.80 |
| **fclrg90.Q** | 0.03 | 0.06 | 0.45 | 6768.47 | 0.65 |
| **fclrg90.C** | -0.08 | 0.08 | -1.01 | 3688.68 | 0.31 |
| **fclrg90^4** | 0.02 | 0.06 | 0.41 | 4700.24 | 0.68 |
| **tenure.L** | 0.11 | 0.05 | 2.15 | 2629.85 | 0.03 |
| **tenure.Q** | -0.06 | 0.06 | -0.90 | 3860.22 | 0.37 |
| **divorce1** | 0.08 | 0.06 | 1.35 | 4660.64 | 0.18 |
| **sepmumbcs1** | 0.09 | 0.11 | 0.78 | 5431.19 | 0.44 |
| **prmnh1** | 0.23 | 0.15 | 1.55 | 1773.40 | 0.12 |
| **crowdUp to 1** | -0.15 | 0.06 | -2.60 | 4617.53 | 0.01 |
| **ameniNo occasions** | -0.05 | 0.11 | -0.45 | 3053.72 | 0.65 |
| **brfed.L** | 0.07 | 0.05 | 1.44 | 3895.95 | 0.15 |
| **brfed.Q** | -0.05 | 0.06 | -0.74 | 3276.90 | 0.46 |
| **resmove.L** | 0.13 | 0.06 | 2.11 | 3979.94 | 0.03 |
| **resmove.Q** | -0.03 | 0.03 | -0.83 | 9230.10 | 0.41 |

*Table S37.* Raw output from modified Poisson regression (with log link) of externalising behaviour at age 16 (extbcsz) against self-reported average sleep duration (binarised to normal vs abnormal) at age 46, adjusted for potential confounders (see main text Table 3). The parameter estimates in the “Estimate” column and the associated standard errors have not been exponentiated.

| Term | Estimate | Standard error | Statistic | Degrees of freedom | p-value |
|---|---|---|---|---|---|
| **(Intercept)** | -1.17 | 0.46 | -2.53 | 1991.59 | 0.01 |
| **extbcsz** | 0.07 | 0.04 | 1.78 | 1173.82 | 0.08 |
| **a0005a** | 0.51 | 0.22 | 2.35 | 2621.65 | 0.02 |
| **a0043b.L** | 0.17 | 0.07 | 2.34 | 2653.14 | 0.02 |
| **a0043b.Q** | 0.19 | 0.10 | 1.87 | 2579.38 | 0.06 |
| **a0043b.C** | 0.05 | 0.09 | 0.49 | 2286.52 | 0.63 |
| **a0043b^4** | -0.11 | 0.09 | -1.16 | 2962.56 | 0.25 |
| **a0043b^5** | -0.07 | 0.12 | -0.58 | 2924.24 | 0.56 |
| **a0195a** | -0.02 | 0.66 | -0.04 | 2460.46 | 0.97 |
| **a0278** | 0.28 | 0.28 | 1.01 | 5126.02 | 0.31 |
| **a0014.L** | 0.17 | 0.15 | 1.12 | 1685.51 | 0.26 |
| **a0014.Q** | 0.05 | 0.08 | 0.63 | 2833.01 | 0.53 |
| **a0014.C** | 0.16 | 0.09 | 1.70 | 2442.28 | 0.09 |
| **a0014^4** | 0.09 | 0.06 | 1.46 | 4032.15 | 0.14 |
| **dv_par_edu_birth** | 0.11 | 0.91 | 0.12 | 2687.67 | 0.91 |
| **dv_sc_age_5.L** | 0.08 | 0.13 | 0.62 | 1240.57 | 0.54 |
| **dv_sc_age_5.Q** | 0.01 | 0.05 | 0.26 | 3895.62 | 0.80 |
| **e216aNone** | 0.09 | 0.07 | 1.18 | 2130.93 | 0.24 |
| **dv_cog_abil_5** | -1.41 | 0.44 | -3.22 | 1119.15 | 0.00 |
| **dv_med_51** | -0.02 | 0.10 | -0.25 | 1559.91 | 0.80 |
| **d016a** | 0.11 | 0.07 | 1.59 | 2739.28 | 0.11 |
| **dv_bas_g** | -0.55 | 0.31 | -1.78 | 1604.31 | 0.07 |
| **dv_med_101** | 0.05 | 0.06 | 0.77 | 2970.74 | 0.44 |
| **bmi** | -0.46 | 0.28 | -1.63 | 2963.68 | 0.10 |
| **fclrg90.L** | 0.23 | 0.10 | 2.33 | 2942.37 | 0.02 |
| **fclrg90.Q** | 0.05 | 0.07 | 0.68 | 3535.98 | 0.50 |
| **fclrg90.C** | -0.04 | 0.10 | -0.39 | 1864.58 | 0.70 |
| **fclrg90^4** | -0.02 | 0.08 | -0.24 | 1957.18 | 0.81 |
| **tenure.L** | 0.06 | 0.07 | 0.81 | 1718.24 | 0.42 |
| **tenure.Q** | -0.05 | 0.09 | -0.55 | 2029.77 | 0.58 |
| **divorce1** | 0.03 | 0.07 | 0.43 | 3138.77 | 0.67 |
| **sepmumbcs1** | 0.33 | 0.13 | 2.53 | 2985.67 | 0.01 |
| **prmnh1** | 0.36 | 0.15 | 2.41 | 1683.05 | 0.02 |
| **crowdUp to 1** | -0.10 | 0.07 | -1.40 | 3035.10 | 0.16 |
| **ameniNo occasions** | 0.01 | 0.11 | 0.13 | 3930.36 | 0.90 |
| **brfed.L** | -0.01 | 0.06 | -0.12 | 2171.13 | 0.91 |
| **brfed.Q** | 0.00 | 0.11 | -0.04 | 1322.56 | 0.97 |
| **resmove.L** | 0.20 | 0.09 | 2.32 | 1758.76 | 0.02 |
| **resmove.Q** | 0.06 | 0.04 | 1.35 | 3793.21 | 0.18 |

*Table S38.* Raw output from modified Poisson regression (with log link) of externalising behaviour at age 16 (extbcsz) against median sleep duration (binarised to normal vs abnormal) derived from the sleep diary at age 46, adjusted for potential confounders (see main text Table 3). The parameter estimates in the “Estimate” column and the associated standard errors have not been exponentiated.

| Term | Estimate | Standard error | Statistic | Degrees of freedom | p-value |
|---|---|---|---|---|---|
| **(Intercept)** | -0.36 | 0.25 | -1.42 | 1975.89 | 0.16 |
| **extbcsz** | 0.01 | 0.01 | 0.76 | 3728.88 | 0.45 |
| **a0005a** | -0.35 | 0.10 | -3.53 | 4908.23 | 0.00 |
| **a0043b.L** | -0.05 | 0.05 | -1.01 | 1814.31 | 0.31 |
| **a0043b.Q** | -0.02 | 0.06 | -0.37 | 2128.21 | 0.71 |
| **a0043b.C** | 0.00 | 0.06 | 0.00 | 1661.53 | 1.00 |
| **a0043b^4** | -0.06 | 0.04 | -1.29 | 3547.67 | 0.20 |
| **a0043b^5** | -0.01 | 0.07 | -0.19 | 2116.12 | 0.85 |
| **a0195a** | -0.37 | 0.46 | -0.82 | 1705.41 | 0.41 |
| **a0278** | -0.21 | 0.20 | -1.08 | 2441.54 | 0.28 |
| **a0014.L** | 0.02 | 0.07 | 0.21 | 1962.36 | 0.83 |
| **a0014.Q** | 0.03 | 0.05 | 0.57 | 1959.23 | 0.57 |
| **a0014.C** | 0.00 | 0.05 | 0.06 | 2546.76 | 0.95 |
| **a0014^4** | -0.01 | 0.04 | -0.27 | 1773.74 | 0.79 |
| **dv_par_edu_birth** | -0.38 | 0.46 | -0.83 | 2706.33 | 0.40 |
| **dv_sc_age_5.L** | -0.02 | 0.06 | -0.25 | 1493.09 | 0.80 |
| **dv_sc_age_5.Q** | 0.00 | 0.03 | -0.15 | 2910.64 | 0.88 |
| **e216aNone** | -0.01 | 0.05 | -0.14 | 1562.08 | 0.89 |
| **dv_cog_abil_5** | 0.04 | 0.28 | 0.16 | 887.63 | 0.87 |
| **dv_med_51** | 0.00 | 0.04 | 0.13 | 2942.36 | 0.90 |
| **d016a** | 0.06 | 0.03 | 1.66 | 3544.85 | 0.10 |
| **dv_bas_g** | -0.23 | 0.16 | -1.41 | 1756.09 | 0.16 |
| **dv_med_101** | 0.01 | 0.03 | 0.33 | 2813.54 | 0.74 |
| **bmi** | 0.20 | 0.17 | 1.14 | 2167.86 | 0.26 |
| **fclrg90.L** | 0.02 | 0.06 | 0.42 | 2486.63 | 0.68 |
| **fclrg90.Q** | 0.05 | 0.04 | 1.06 | 2486.15 | 0.29 |
| **fclrg90.C** | 0.06 | 0.04 | 1.50 | 4043.76 | 0.13 |
| **fclrg90^4** | 0.01 | 0.05 | 0.27 | 1749.36 | 0.79 |
| **tenure.L** | 0.01 | 0.03 | 0.45 | 2349.90 | 0.65 |
| **tenure.Q** | -0.03 | 0.05 | -0.68 | 2249.37 | 0.49 |
| **divorce1** | 0.03 | 0.04 | 0.74 | 4081.30 | 0.46 |
| **sepmumbcs1** | -0.06 | 0.09 | -0.72 | 2989.58 | 0.47 |
| **prmnh1** | 0.18 | 0.09 | 1.88 | 1785.66 | 0.06 |
| **crowdUp to 1** | -0.05 | 0.05 | -0.87 | 1881.13 | 0.39 |
| **ameniNo occasions** | -0.08 | 0.07 | -1.06 | 2166.96 | 0.29 |
| **brfed.L** | 0.03 | 0.03 | 0.81 | 2105.31 | 0.42 |
| **brfed.Q** | -0.01 | 0.03 | -0.41 | 3457.78 | 0.68 |
| **resmove.L** | -0.03 | 0.04 | -0.74 | 2754.74 | 0.46 |
| **resmove.Q** | 0.06 | 0.02 | 2.49 | 4355.18 | 0.01 |

*Table S39.* Raw output from modified Poisson regression (with log link) of externalising behaviour at age 16 (extbcsz) against median sleep duration (binarised to normal vs abnormal) derived from the activPAL algorithm at age 46, adjusted for potential confounders (see main text Table 3). The parameter estimates in the “Estimate” column and the associated standard errors have not been exponentiated.

| Term | Estimate | Standard error | Statistic | Degrees of freedom | p-value |
|---|---|---|---|---|---|
| **(Intercept)** | -0.92 | 0.36 | -2.57 | 1403.79 | 0.01 |
| **extbcsz** | 0.03 | 0.03 | 1.31 | 1241.36 | 0.19 |
| **a0005a** | 0.00 | 0.17 | -0.03 | 1771.67 | 0.98 |
| **a0043b.L** | 0.01 | 0.06 | 0.09 | 1600.74 | 0.93 |
| **a0043b.Q** | -0.06 | 0.07 | -0.92 | 1797.51 | 0.36 |
| **a0043b.C** | -0.18 | 0.06 | -2.80 | 1746.51 | 0.01 |
| **a0043b^4** | 0.10 | 0.06 | 1.63 | 2069.66 | 0.10 |
| **a0043b^5** | 0.03 | 0.08 | 0.42 | 2096.46 | 0.67 |
| **a0195a** | -0.24 | 0.47 | -0.51 | 1963.72 | 0.61 |
| **a0278** | 0.36 | 0.18 | 2.02 | 4031.41 | 0.04 |
| **a0014.L** | 0.03 | 0.09 | 0.32 | 1540.44 | 0.75 |
| **a0014.Q** | 0.04 | 0.07 | 0.62 | 1556.28 | 0.54 |
| **a0014.C** | -0.01 | 0.05 | -0.15 | 2618.34 | 0.88 |
| **a0014^4** | -0.02 | 0.04 | -0.50 | 2385.42 | 0.62 |
| **dv_par_edu_birth** | -0.11 | 0.41 | -0.27 | 3881.13 | 0.78 |
| **dv_sc_age_5.L** | 0.15 | 0.08 | 1.82 | 1158.27 | 0.07 |
| **dv_sc_age_5.Q** | -0.03 | 0.04 | -0.94 | 2176.06 | 0.35 |
| **e216aNone** | 0.07 | 0.05 | 1.34 | 1785.72 | 0.18 |
| **dv_cog_abil_5** | 0.00 | 0.27 | 0.01 | 1060.59 | 0.99 |
| **dv_med_51** | 0.04 | 0.05 | 0.77 | 1871.70 | 0.44 |
| **d016a** | 0.08 | 0.04 | 1.85 | 2372.62 | 0.06 |
| **dv_bas_g** | -0.40 | 0.22 | -1.80 | 1261.19 | 0.07 |
| **dv_med_101** | 0.04 | 0.05 | 0.84 | 1577.23 | 0.40 |
| **bmi** | -0.25 | 0.24 | -1.03 | 1564.65 | 0.30 |
| **fclrg90.L** | -0.01 | 0.06 | -0.14 | 2564.92 | 0.89 |
| **fclrg90.Q** | -0.03 | 0.06 | -0.45 | 1666.26 | 0.66 |
| **fclrg90.C** | -0.10 | 0.06 | -1.77 | 2152.42 | 0.08 |
| **fclrg90^4** | 0.01 | 0.05 | 0.20 | 2089.36 | 0.84 |
| **tenure.L** | 0.03 | 0.05 | 0.68 | 1401.17 | 0.50 |
| **tenure.Q** | 0.09 | 0.06 | 1.62 | 2250.13 | 0.10 |
| **divorce1** | 0.14 | 0.05 | 2.81 | 2553.05 | 0.01 |
| **sepmumbcs1** | -0.12 | 0.14 | -0.84 | 1581.54 | 0.40 |
| **prmnh1** | -0.23 | 0.19 | -1.23 | 1122.74 | 0.22 |
| **crowdUp to 1** | 0.03 | 0.06 | 0.55 | 1862.86 | 0.58 |
| **ameniNo occasions** | -0.09 | 0.08 | -1.09 | 2366.70 | 0.27 |
| **brfed.L** | 0.01 | 0.04 | 0.23 | 1602.79 | 0.82 |
| **brfed.Q** | 0.04 | 0.05 | 0.93 | 2253.47 | 0.35 |
| **resmove.L** | 0.00 | 0.04 | 0.02 | 3594.22 | 0.99 |
| **resmove.Q** | -0.06 | 0.03 | -2.28 | 3920.65 | 0.02 |

*Table S40.* Raw output from modified Poisson regression (with log link) of externalising behaviour at age 16 (extbcsz) against median sleep duration (binarised to normal vs abnormal) derived from the van der Berg et al. algorithm at age 46, adjusted for potential confounders (see main text Table 3). The parameter estimates in the “Estimate” column and the associated standard errors have not been exponentiated.

| Term | Estimate | Standard error | Statistic | Degrees of freedom | p-value |
|---|---|---|---|---|---|
| **(Intercept)** | -0.96 | 0.29 | -3.27 | 1685.86 | 0.00 |
| **extbcsz** | 0.07 | 0.02 | 3.13 | 1223.42 | 0.00 |
| **a0005a** | 0.16 | 0.20 | 0.80 | 1259.74 | 0.42 |
| **a0043b.L** | -0.05 | 0.04 | -1.17 | 3022.74 | 0.24 |
| **a0043b.Q** | 0.02 | 0.06 | 0.37 | 2184.55 | 0.71 |
| **a0043b.C** | 0.06 | 0.04 | 1.63 | 4506.30 | 0.10 |
| **a0043b^4** | 0.03 | 0.05 | 0.53 | 2696.09 | 0.60 |
| **a0043b^5** | 0.03 | 0.07 | 0.50 | 2435.77 | 0.61 |
| **a0195a** | -0.03 | 0.33 | -0.09 | 3966.55 | 0.93 |
| **a0278** | -0.11 | 0.27 | -0.39 | 1578.32 | 0.70 |
| **a0014.L** | 0.08 | 0.07 | 1.06 | 2289.44 | 0.29 |
| **a0014.Q** | 0.01 | 0.06 | 0.15 | 1597.24 | 0.88 |
| **a0014.C** | -0.03 | 0.09 | -0.38 | 1093.88 | 0.70 |
| **a0014^4** | 0.03 | 0.05 | 0.62 | 1548.53 | 0.54 |
| **dv_par_edu_birth** | 0.46 | 0.43 | 1.09 | 1831.72 | 0.28 |
| **dv_sc_age_5.L** | 0.02 | 0.09 | 0.18 | 1021.15 | 0.85 |
| **dv_sc_age_5.Q** | -0.01 | 0.05 | -0.16 | 1431.49 | 0.87 |
| **e216aNone** | -0.12 | 0.05 | -2.26 | 1375.56 | 0.02 |
| **dv_cog_abil_5** | -0.23 | 0.27 | -0.87 | 1004.58 | 0.38 |
| **dv_med_51** | 0.02 | 0.04 | 0.53 | 2725.68 | 0.60 |
| **d016a** | 0.18 | 0.06 | 3.26 | 1383.22 | 0.00 |
| **dv_bas_g** | -0.23 | 0.18 | -1.27 | 1538.07 | 0.21 |
| **dv_med_101** | 0.03 | 0.05 | 0.68 | 1618.84 | 0.50 |
| **bmi** | 0.55 | 0.20 | 2.68 | 1687.20 | 0.01 |
| **fclrg90.L** | -0.13 | 0.08 | -1.52 | 1600.85 | 0.13 |
| **fclrg90.Q** | -0.05 | 0.06 | -0.94 | 1814.62 | 0.35 |
| **fclrg90.C** | 0.02 | 0.07 | 0.35 | 1502.37 | 0.72 |
| **fclrg90^4** | 0.01 | 0.05 | 0.10 | 1733.89 | 0.92 |
| **tenure.L** | 0.04 | 0.04 | 0.84 | 1520.33 | 0.40 |
| **tenure.Q** | -0.04 | 0.06 | -0.74 | 1580.86 | 0.46 |
| **divorce1** | -0.03 | 0.06 | -0.45 | 1575.69 | 0.65 |
| **sepmumbcs1** | 0.19 | 0.10 | 1.98 | 1991.06 | 0.05 |
| **prmnh1** | -0.07 | 0.10 | -0.63 | 2152.32 | 0.53 |
| **crowdUp to 1** | 0.08 | 0.05 | 1.62 | 2260.17 | 0.10 |
| **ameniNo occasions** | -0.16 | 0.10 | -1.69 | 1497.12 | 0.09 |
| **brfed.L** | 0.03 | 0.03 | 0.89 | 2279.01 | 0.38 |
| **brfed.Q** | -0.05 | 0.05 | -0.91 | 1609.68 | 0.36 |
| **resmove.L** | 0.02 | 0.06 | 0.29 | 1429.28 | 0.77 |
| **resmove.Q** | 0.02 | 0.02 | 0.97 | 5312.09 | 0.33 |

*Table S41.* Raw output from modified Poisson regression (with log link) of externalising behaviour at age 16 (extbcsz) against median sleep duration (binarised to normal vs abnormal) derived from the Winkler et al. algorithm at age 46, adjusted for potential confounders (see main text Table 3). The parameter estimates in the “Estimate” column and the associated standard errors have not been exponentiated.

**Poisson regression outputs (mediation, part 1): Table S42**

| Dependent variable | Independent variable | Risk ratio (95% CI) | p-value | E-value |
|---|---|---|---|---|
| **Malaise (age 42)*** | **Rutter (age 5)** | 1.106 (1.082, 1.130) | 0.000 | 1.380 |
| **WEMWBS (age 42)*** | **Rutter (age 5)** | 0.973 (0.967, 0.979) | 0.000 | 1.169 |
| **Malaise (age 42)*** | **Rutter (age 10)** | 1.115 (1.092, 1.139) | 0.000 | 1.409 |
| **WEMWBS (age 42)*** | **Rutter (age 10)** | 0.969 (0.963, 0.975) | 0.000 | 1.189 |
| **Malaise (age 42)*** | **Child Development Scale (age 10)** | 1.072 (1.044, 1.099) | 0.000 | 1.260 |
| **WEMWBS (age 42)*** | **Child Development Scale (age 10)** | 0.980 (0.970, 0.990) | 0.000 | 1.111 |
| **Malaise (age 42)*** | **Malaise (age 16)*** | 1.291 (1.263, 1.319) | 0.000 | 1.840 |
| **WEMWBS (age 42)*** | **Malaise (age 16)*** | 0.949 (0.942, 0.956) | 0.000 | 1.264 |
| **Malaise (age 42)*** | **Behavioural & emotional problems (age 16)** | 1.045 (1.018, 1.072) | 0.001 | 1.155 |
| **WEMWBS (age 42)*** | **Behavioural & emotional problems (age 16)** | 0.988 (0.980, 0.995) | 0.002 | 1.074 |
| **Malaise (age 42)*** | **Internalising behaviour (age 16)** | 1.185 (1.164, 1.206) | 0.000 | 1.601 |
| **WEMWBS (age 42)*** | **Internalising behaviour (age 16)** | 0.960 (0.955, 0.965) | 0.000 | 1.230 |
| **Malaise (age 42)*** | **Externalising behaviour (age 16)** | 1.117 (1.091, 1.144) | 0.000 | 1.406 |
| **WEMWBS (age 42)*** | **Externalising behaviour (age 16)** | 0.966 (0.959, 0.973) | 0.000 | 1.196 |

*Table S42.* Estimated risk ratios quantifying the associations between mental health variables in childhood and at age 42. The E-value, quantifying the minimum unmeasured confounding risk ratio that would be needed to nullify each association estimated here, is also shown. Each exposure-outcome relationship was assessed in a separate model. All models were adjusted for a range of socioeconomic, perinatal and health-related covariates. Missing data were handled by multiple imputation. RR = risk ratio. * = self-reported.

**Raw Poisson regression outputs (mediation, part 2): Tables S43–77**

The following pages contain raw model outputs for the modified Poisson regressions adjusting for adult mental health used to produce main text Figure 2. The tables are as follows:

| **Sleep measure ⇒**<br><br>**Mental health measure ⇓** | **Self-report** | **Sleep diary** | **activPAL algorithm** | **van der Berg et al. algorithm** | **Winkler et al. algorithm** |
|---|---|---|---|---|---|
| **Rutter score (age 5)** | Table S43 | Table S44 | Table S45 | Table S46 | Table S47 |
| **Rutter score (age 10)** | Table S48 | Table S49 | Table S50 | Table S51 | Table S52 |
| **Child Development Scale (age 10)** | Table S53 | Table S54 | Table S55 | Table S56 | Table S57 |
| **Malaise Inventory (age 16)** | Table S58 | Table S59 | Table S60 | Table S61 | Table S62 |
| **Behavioural & emotional problems (age 16)** | Table S63 | Table S64 | Table S65 | Table S66 | Table S67 |
| **Internalising behaviour (age 16)** | Table S68 | Table S69 | Table S70 | Table S71 | Table S72 |
| **Externalising behaviour (age 16)** | Table S73 | Table S74 | Table S75 | Table S76 | Table S77 |

| Term | Estimate | Standard error | Statistic | Degrees of freedom | p-value |
|---|---|---|---|---|---|
| **(Intercept)** | -0.63 | 0.39 | -1.60 | 2917.77 | 0.11 |
| **d119** | 0.05 | 0.02 | 2.00 | 3474.84 | 0.05 |
| **BD9MAL** | 1.04 | 0.15 | 7.00 | 2199.02 | 0.00 |
| **BD9WEMWB** | -1.24 | 0.23 | -5.45 | 2751.70 | 0.00 |
| **a0005a** | -0.39 | 0.14 | -2.70 | 9606.77 | 0.01 |
| **a0043b.L** | 0.21 | 0.06 | 3.37 | 3291.11 | 0.00 |
| **a0043b.Q** | 0.08 | 0.09 | 0.92 | 2775.23 | 0.36 |
| **a0043b.C** | 0.03 | 0.08 | 0.42 | 2825.95 | 0.68 |
| **a0043b^4** | -0.03 | 0.11 | -0.29 | 2023.23 | 0.78 |
| **a0043b^5** | -0.06 | 0.11 | -0.53 | 2873.71 | 0.60 |
| **a0195a** | -0.24 | 0.50 | -0.48 | 4470.22 | 0.63 |
| **a0278** | -0.76 | 0.24 | -3.15 | 8594.72 | 0.00 |
| **a0014.L** | 0.44 | 0.08 | 5.60 | 4316.32 | 0.00 |
| **a0014.Q** | 0.02 | 0.06 | 0.30 | 5767.65 | 0.77 |
| **a0014.C** | 0.08 | 0.08 | 1.01 | 3117.60 | 0.31 |
| **a0014^4** | 0.09 | 0.06 | 1.41 | 2900.71 | 0.16 |
| **dv_par_edu_birth** | -1.70 | 1.44 | -1.18 | 2675.12 | 0.24 |
| **brfed.L** | 0.11 | 0.05 | 2.13 | 3678.13 | 0.03 |
| **brfed.Q** | -0.04 | 0.06 | -0.63 | 3969.92 | 0.53 |

*Table S43.* Raw output from modified Poisson regression (with log link) of Rutter score at age 5 (d119) against self-reported average sleep duration (binarised to normal vs abnormal) at age 46, adjusted for potential confounders (see main text Table 3) and mental health variables at age 42 (see Table S42). The parameter estimates in the “Estimate” column and the associated standard errors have not been exponentiated.

| Term | Estimate | Standard error | Statistic | Degrees of freedom | p-value |
|---|---|---|---|---|---|
| **(Intercept)** | -1.31 | 0.51 | -2.55 | 1820.01 | 0.01 |
| **d119** | 0.06 | 0.04 | 1.81 | 1709.45 | 0.07 |
| **BD9MAL** | 0.44 | 0.13 | 3.34 | 3967.19 | 0.00 |
| **BD9WEMWB** | -1.13 | 0.24 | -4.62 | 2746.01 | 0.00 |
| **a0005a** | 0.34 | 0.21 | 1.62 | 2567.69 | 0.10 |
| **a0043b.L** | 0.20 | 0.08 | 2.65 | 2282.32 | 0.01 |
| **a0043b.Q** | 0.18 | 0.10 | 1.76 | 2428.65 | 0.08 |
| **a0043b.C** | 0.05 | 0.09 | 0.58 | 2342.49 | 0.56 |
| **a0043b^4** | -0.11 | 0.09 | -1.16 | 3041.74 | 0.25 |
| **a0043b^5** | -0.06 | 0.12 | -0.53 | 3127.88 | 0.59 |
| **a0195a** | -0.13 | 0.63 | -0.21 | 3016.97 | 0.83 |
| **a0278** | -0.11 | 0.30 | -0.36 | 4097.70 | 0.72 |
| **a0014.L** | 0.49 | 0.10 | 4.96 | 2267.02 | 0.00 |
| **a0014.Q** | 0.09 | 0.08 | 1.20 | 2549.99 | 0.23 |
| **a0014.C** | 0.14 | 0.09 | 1.66 | 2647.96 | 0.10 |
| **a0014^4** | 0.12 | 0.05 | 2.26 | 5157.90 | 0.02 |
| **dv_par_edu_birth** | -0.50 | 1.22 | -0.41 | 2450.43 | 0.68 |
| **brfed.L** | 0.04 | 0.07 | 0.61 | 2098.80 | 0.54 |
| **brfed.Q** | 0.00 | 0.10 | 0.04 | 1534.07 | 0.97 |

*Table S44.* Raw output from modified Poisson regression (with log link) of Rutter score at age 5 (d119) against median sleep duration (binarised to normal vs abnormal) derived from the sleep diary at age 46, adjusted for potential confounders (see main text Table 3) and mental health variables at age 42 (see Table S42). The parameter estimates in the "Estimate" column and the associated standard errors have not been exponentiated.

| Term | Estimate | Standard error | Statistic | Degrees of freedom | p-value |
|---|---|---|---|---|---|
| **(Intercept)** | -0.53 | 0.25 | -2.12 | 2025.12 | 0.03 |
| **d119** | -0.01 | 0.02 | -0.66 | 1777.81 | 0.51 |
| **BD9MAL** | 0.15 | 0.10 | 1.57 | 2147.30 | 0.12 |
| **BD9WEMWB** | 0.02 | 0.17 | 0.14 | 1787.55 | 0.89 |
| **a0005a** | -0.32 | 0.10 | -3.27 | 4173.19 | 0.00 |
| **a0043b.L** | -0.03 | 0.05 | -0.61 | 1771.68 | 0.54 |
| **a0043b.Q** | -0.02 | 0.06 | -0.31 | 2123.83 | 0.76 |
| **a0043b.C** | 0.01 | 0.06 | 0.09 | 1638.42 | 0.93 |
| **a0043b^4** | -0.06 | 0.04 | -1.30 | 3763.51 | 0.19 |
| **a0043b^5** | -0.02 | 0.07 | -0.25 | 2265.49 | 0.80 |
| **a0195a** | -0.43 | 0.47 | -0.91 | 1667.92 | 0.37 |
| **a0278** | -0.22 | 0.21 | -1.02 | 2029.47 | 0.31 |
| **a0014.L** | 0.07 | 0.06 | 1.14 | 2025.50 | 0.25 |
| **a0014.Q** | 0.03 | 0.05 | 0.69 | 1852.95 | 0.49 |
| **a0014.C** | 0.00 | 0.05 | 0.08 | 2533.40 | 0.94 |
| **a0014^4** | -0.01 | 0.04 | -0.16 | 1816.28 | 0.88 |
| **dv_par_edu_birth** | -0.49 | 0.50 | -0.99 | 2650.59 | 0.32 |
| **brfed.L** | 0.04 | 0.03 | 1.10 | 2226.93 | 0.27 |
| **brfed.Q** | -0.01 | 0.03 | -0.43 | 3359.49 | 0.67 |

*Table S45.* Raw output from modified Poisson regression (with log link) of Rutter score at age 5 (d119) against median sleep duration (binarised to normal vs abnormal) derived from the activPAL algorithm at age 46, adjusted for potential confounders (see main text Table 3) and mental health variables at age 42 (see Table S42). The parameter estimates in the "Estimate" column and the associated standard errors have not been exponentiated.

| Term | Estimate | Standard error | Statistic | Degrees of freedom | p-value |
|---|---|---|---|---|---|
| **(Intercept)** | -0.92 | 0.30 | -3.09 | 1817.55 | 0.00 |
| **d119** | 0.02 | 0.02 | 0.89 | 2816.93 | 0.37 |
| **BD9MAL** | 0.18 | 0.12 | 1.52 | 1855.09 | 0.13 |
| **BD9WEMWB** | -0.22 | 0.19 | -1.15 | 1724.69 | 0.25 |
| **a0005a** | -0.10 | 0.15 | -0.67 | 1871.80 | 0.50 |
| **a0043b.L** | 0.02 | 0.06 | 0.30 | 1693.56 | 0.76 |
| **a0043b.Q** | -0.07 | 0.07 | -1.05 | 1730.52 | 0.30 |
| **a0043b.C** | -0.18 | 0.07 | -2.62 | 1648.37 | 0.01 |
| **a0043b^4** | 0.10 | 0.06 | 1.60 | 1999.08 | 0.11 |
| **a0043b^5** | 0.03 | 0.08 | 0.42 | 2167.72 | 0.67 |
| **a0195a** | -0.28 | 0.45 | -0.63 | 2212.57 | 0.53 |
| **a0278** | 0.25 | 0.18 | 1.40 | 3798.04 | 0.16 |
| **a0014.L** | 0.21 | 0.09 | 2.33 | 1197.46 | 0.02 |
| **a0014.Q** | 0.06 | 0.08 | 0.74 | 1189.89 | 0.46 |
| **a0014.C** | -0.04 | 0.05 | -0.87 | 2827.50 | 0.39 |
| **a0014^4** | 0.01 | 0.04 | 0.29 | 2639.30 | 0.77 |
| **dv_par_edu_birth** | -0.31 | 0.42 | -0.74 | 4302.92 | 0.46 |
| **brfed.L** | 0.02 | 0.04 | 0.57 | 1671.96 | 0.57 |
| **brfed.Q** | 0.05 | 0.05 | 1.04 | 2284.06 | 0.30 |

*Table S46.* Raw output from modified Poisson regression (with log link) of Rutter score at age 5 (d119) against median sleep duration (binarised to normal vs abnormal) derived from the van der Berg et al. algorithm at age 46, adjusted for potential confounders (see main text Table 3) and mental health variables at age 42 (see Table S42). The parameter estimates in the “Estimate” column and the associated standard errors have not been exponentiated.

| Term | Estimate | Standard error | Statistic | Degrees of freedom | p-value |
|---|---|---|---|---|---|
| **(Intercept)** | -1.05 | 0.25 | -4.14 | 2094.40 | 0.00 |
| **d119** | 0.08 | 0.02 | 3.94 | 1830.43 | 0.00 |
| **BD9MAL** | 0.27 | 0.13 | 2.04 | 1379.47 | 0.04 |
| **BD9WEMWB** | -0.13 | 0.24 | -0.54 | 1159.47 | 0.59 |
| **a0005a** | 0.06 | 0.18 | 0.31 | 1328.14 | 0.76 |
| **a0043b.L** | -0.03 | 0.04 | -0.72 | 2794.57 | 0.47 |
| **a0043b.Q** | 0.02 | 0.06 | 0.26 | 2158.16 | 0.80 |
| **a0043b.C** | 0.07 | 0.04 | 1.74 | 5412.71 | 0.08 |
| **a0043b^4** | 0.03 | 0.05 | 0.53 | 2640.14 | 0.60 |
| **a0043b^5** | 0.04 | 0.07 | 0.60 | 2469.54 | 0.55 |
| **a0195a** | -0.10 | 0.34 | -0.29 | 3862.40 | 0.77 |
| **a0278** | -0.09 | 0.27 | -0.35 | 1593.43 | 0.73 |
| **a0014.L** | 0.11 | 0.07 | 1.65 | 1741.13 | 0.10 |
| **a0014.Q** | 0.01 | 0.06 | 0.09 | 1653.22 | 0.93 |
| **a0014.C** | -0.04 | 0.08 | -0.48 | 1131.32 | 0.63 |
| **a0014^4** | 0.04 | 0.05 | 0.76 | 1572.97 | 0.45 |
| **dv_par_edu_birth** | 0.48 | 0.45 | 1.08 | 1737.02 | 0.28 |
| **brfed.L** | 0.04 | 0.04 | 1.03 | 2141.12 | 0.30 |
| **brfed.Q** | -0.04 | 0.05 | -0.82 | 1589.73 | 0.41 |

*Table S47.* Raw output from modified Poisson regression (with log link) of Rutter score at age 5 (d119) against median sleep duration (binarised to normal vs abnormal) derived from the Winkler et al. algorithm at age 46, adjusted for potential confounders (see main text Table 3) and mental health variables at age 42 (see Table S42). The parameter estimates in the “Estimate” column and the associated standard errors have not been exponentiated.

| Term | Estimate | Standard error | Statistic | Degrees of freedom | p-value |
|---|---|---|---|---|---|
| **(Intercept)** | -0.49 | 0.40 | -1.23 | 2738.35 | 0.22 |
| **BD3MRUTT** | 0.02 | 0.02 | 0.94 | 3855.52 | 0.35 |
| **BD9MAL** | 1.03 | 0.14 | 7.14 | 2283.11 | 0.00 |
| **BD9WEMWB** | -1.12 | 0.23 | -4.90 | 2664.18 | 0.00 |
| **a0005a** | -0.26 | 0.15 | -1.75 | 9115.03 | 0.08 |
| **a0043b.L** | 0.16 | 0.06 | 2.58 | 3347.91 | 0.01 |
| **a0043b.Q** | 0.06 | 0.09 | 0.71 | 2821.31 | 0.48 |
| **a0043b.C** | 0.02 | 0.08 | 0.30 | 2945.30 | 0.76 |
| **a0043b^4** | -0.02 | 0.11 | -0.16 | 2013.12 | 0.87 |
| **a0043b^5** | -0.07 | 0.11 | -0.59 | 2854.80 | 0.56 |
| **a0195a** | -0.18 | 0.49 | -0.36 | 4566.51 | 0.72 |
| **a0278** | -0.57 | 0.24 | -2.34 | 7485.38 | 0.02 |
| **a0014.L** | 0.16 | 0.11 | 1.48 | 2625.76 | 0.14 |
| **a0014.Q** | 0.00 | 0.06 | -0.05 | 6280.16 | 0.96 |
| **a0014.C** | 0.11 | 0.08 | 1.39 | 2908.04 | 0.16 |
| **a0014^4** | 0.06 | 0.07 | 0.87 | 2557.33 | 0.38 |
| **dv_par_edu_birth** | -1.18 | 1.25 | -0.95 | 2826.63 | 0.34 |
| **dv_sc_age_5.L** | 0.19 | 0.10 | 1.94 | 1628.11 | 0.05 |
| **dv_sc_age_5.Q** | -0.06 | 0.04 | -1.47 | 5444.40 | 0.14 |
| **e216aNone** | 0.00 | 0.06 | 0.02 | 3026.51 | 0.99 |
| **dv_cog_abil_5** | -0.69 | 0.29 | -2.33 | 1650.38 | 0.02 |
| **dv_med_51** | 0.03 | 0.06 | 0.59 | 4371.40 | 0.56 |
| **d016a** | 0.21 | 0.05 | 4.11 | 4486.17 | 0.00 |
| **sepmumbcs1** | 0.13 | 0.11 | 1.16 | 3914.42 | 0.25 |
| **crowdUp to 1** | -0.21 | 0.06 | -3.39 | 3283.58 | 0.00 |
| **brfed.L** | 0.09 | 0.05 | 1.73 | 3858.85 | 0.08 |
| **brfed.Q** | -0.05 | 0.06 | -0.79 | 3875.85 | 0.43 |
| **resmove.L** | 0.17 | 0.06 | 3.10 | 4438.11 | 0.00 |
| **resmove.Q** | -0.02 | 0.03 | -0.62 | 9499.22 | 0.54 |

*Table S48.* Raw output from modified Poisson regression (with log link) of Rutter score at age 10 (BD3MRUTT) against self-reported average sleep duration (binarised to normal vs abnormal) at age 46, adjusted for potential confounders (see main text Table 3) and mental health variables at age 42 (see Table S42). The parameter estimates in the “Estimate” column and the associated standard errors have not been exponentiated.

| Term | Estimate | Standard error | Statistic | Degrees of freedom | p-value |
|---|---|---|---|---|---|
| **(Intercept)** | -0.96 | 0.54 | -1.78 | 1602.02 | 0.07 |
| **BD3MRUTT** | 0.08 | 0.03 | 3.05 | 3198.75 | 0.00 |
| **BD9MAL** | 0.41 | 0.13 | 3.13 | 3665.93 | 0.00 |
| **BD9WEMWB** | -0.95 | 0.23 | -4.08 | 2991.86 | 0.00 |
| **a0005a** | 0.50 | 0.21 | 2.32 | 2609.40 | 0.02 |
| **a0043b.L** | 0.16 | 0.07 | 2.23 | 2505.52 | 0.03 |
| **a0043b.Q** | 0.17 | 0.10 | 1.66 | 2461.85 | 0.10 |
| **a0043b.C** | 0.04 | 0.09 | 0.42 | 2245.05 | 0.68 |
| **a0043b^4** | -0.10 | 0.09 | -1.05 | 2863.92 | 0.29 |
| **a0043b^5** | -0.07 | 0.12 | -0.59 | 3361.72 | 0.55 |
| **a0195a** | -0.06 | 0.66 | -0.09 | 2502.33 | 0.93 |
| **a0278** | 0.18 | 0.28 | 0.63 | 4872.59 | 0.53 |
| **a0014.L** | 0.24 | 0.14 | 1.70 | 1693.63 | 0.09 |
| **a0014.Q** | 0.06 | 0.08 | 0.73 | 2517.20 | 0.47 |
| **a0014.C** | 0.16 | 0.09 | 1.84 | 2774.25 | 0.07 |
| **a0014^4** | 0.10 | 0.06 | 1.69 | 4078.58 | 0.09 |
| **dv_par_edu_birth** | -0.05 | 0.98 | -0.05 | 2611.03 | 0.96 |
| **dv_sc_age_5.L** | 0.13 | 0.11 | 1.19 | 1366.18 | 0.23 |
| **dv_sc_age_5.Q** | 0.00 | 0.05 | -0.02 | 3486.87 | 0.98 |
| **e216aNone** | 0.10 | 0.07 | 1.39 | 2170.99 | 0.17 |
| **dv_cog_abil_5** | -1.53 | 0.37 | -4.09 | 1264.84 | 0.00 |
| **dv_med_51** | -0.02 | 0.10 | -0.20 | 1498.01 | 0.84 |
| **d016a** | 0.11 | 0.07 | 1.63 | 2707.40 | 0.10 |
| **sepmumbcs1** | 0.33 | 0.12 | 2.68 | 2857.73 | 0.01 |
| **crowdUp to 1** | -0.14 | 0.06 | -2.23 | 3399.98 | 0.03 |
| **brfed.L** | 0.00 | 0.06 | 0.07 | 2146.54 | 0.94 |
| **brfed.Q** | -0.01 | 0.10 | -0.05 | 1454.52 | 0.96 |
| **resmove.L** | 0.22 | 0.08 | 2.61 | 1756.55 | 0.01 |
| **resmove.Q** | 0.06 | 0.04 | 1.35 | 3669.88 | 0.18 |

*Table S49.* Raw output from modified Poisson regression (with log link) of Rutter score at age 10 (BD3MRUTT) against median sleep duration (binarised to normal vs abnormal) derived from the sleep diary at age 46, adjusted for potential confounders (see main text Table 3) and mental health variables at age 42 (see Table S42). The parameter estimates in the “Estimate” column and the associated standard errors have not been exponentiated.

| Term | Estimate | Standard error | Statistic | Degrees of freedom | p-value |
|---|---|---|---|---|---|
| **(Intercept)** | -0.48 | 0.24 | -2.02 | 2376.62 | 0.04 |
| **BD3MRUTT** | 0.01 | 0.02 | 0.53 | 2711.88 | 0.59 |
| **BD9MAL** | 0.15 | 0.10 | 1.54 | 2174.09 | 0.12 |
| **BD9WEMWB** | 0.05 | 0.18 | 0.28 | 1630.79 | 0.78 |
| **a0005a** | -0.37 | 0.10 | -3.86 | 5716.43 | 0.00 |
| **a0043b.L** | -0.04 | 0.05 | -0.76 | 1668.00 | 0.45 |
| **a0043b.Q** | -0.02 | 0.06 | -0.38 | 2129.18 | 0.70 |
| **a0043b.C** | 0.00 | 0.06 | 0.03 | 1645.16 | 0.98 |
| **a0043b^4** | -0.05 | 0.04 | -1.24 | 3639.35 | 0.21 |
| **a0043b^5** | -0.02 | 0.07 | -0.24 | 2205.67 | 0.81 |
| **a0195a** | -0.40 | 0.47 | -0.84 | 1656.00 | 0.40 |
| **a0278** | -0.21 | 0.21 | -1.00 | 2184.54 | 0.32 |
| **a0014.L** | 0.04 | 0.07 | 0.50 | 1966.99 | 0.62 |
| **a0014.Q** | 0.04 | 0.05 | 0.69 | 1940.13 | 0.49 |
| **a0014.C** | 0.01 | 0.04 | 0.19 | 2779.10 | 0.85 |
| **a0014^4** | -0.01 | 0.04 | -0.27 | 1953.63 | 0.79 |
| **dv_par_edu_birth** | -0.45 | 0.48 | -0.94 | 2662.75 | 0.35 |
| **dv_sc_age_5.L** | 0.00 | 0.05 | -0.01 | 2027.66 | 0.99 |
| **dv_sc_age_5.Q** | 0.00 | 0.03 | -0.05 | 3877.58 | 0.96 |
| **e216aNone** | -0.01 | 0.05 | -0.20 | 1545.10 | 0.85 |
| **dv_cog_abil_5** | -0.05 | 0.25 | -0.18 | 930.73 | 0.86 |
| **dv_med_51** | 0.01 | 0.04 | 0.28 | 2945.47 | 0.78 |
| **d016a** | 0.06 | 0.03 | 1.83 | 3424.70 | 0.07 |
| **sepmumbcs1** | -0.04 | 0.09 | -0.48 | 2622.58 | 0.63 |
| **crowdUp to 1** | -0.07 | 0.05 | -1.42 | 2190.31 | 0.15 |
| **brfed.L** | 0.03 | 0.03 | 0.99 | 2102.10 | 0.32 |
| **brfed.Q** | -0.01 | 0.03 | -0.43 | 3364.28 | 0.67 |
| **resmove.L** | -0.02 | 0.04 | -0.50 | 2637.76 | 0.61 |
| **resmove.Q** | 0.06 | 0.02 | 2.58 | 4302.26 | 0.01 |

*Table S50.* Raw output from modified Poisson regression (with log link) of Rutter score at age 10 (BD3MRUTT) against median sleep duration (binarised to normal vs abnormal) derived from the activPAL algorithm at age 46, adjusted for potential confounders (see main text Table 3) and mental health variables at age 42 (see Table S42). The parameter estimates in the “Estimate” column and the associated standard errors have not been exponentiated.

| Term | Estimate | Standard error | Statistic | Degrees of freedom | p-value |
|---|---|---|---|---|---|
| **(Intercept)** | -1.04 | 0.34 | -3.10 | 1577.42 | 0.00 |
| **BD3MRUTT** | 0.02 | 0.02 | 0.73 | 1654.88 | 0.46 |
| **BD9MAL** | 0.18 | 0.12 | 1.51 | 1803.22 | 0.13 |
| **BD9WEMWB** | -0.16 | 0.19 | -0.85 | 1735.56 | 0.39 |
| **a0005a** | -0.06 | 0.17 | -0.35 | 1743.10 | 0.72 |
| **a0043b.L** | 0.00 | 0.06 | 0.08 | 1724.35 | 0.93 |
| **a0043b.Q** | -0.08 | 0.07 | -1.16 | 1823.74 | 0.25 |
| **a0043b.C** | -0.18 | 0.07 | -2.79 | 1719.91 | 0.01 |
| **a0043b^4** | 0.11 | 0.06 | 1.76 | 2056.92 | 0.08 |
| **a0043b^5** | 0.03 | 0.07 | 0.42 | 2194.13 | 0.68 |
| **a0195a** | -0.24 | 0.46 | -0.53 | 2114.61 | 0.60 |
| **a0278** | 0.32 | 0.17 | 1.81 | 4295.17 | 0.07 |
| **a0014.L** | 0.05 | 0.10 | 0.50 | 1403.11 | 0.62 |
| **a0014.Q** | 0.04 | 0.07 | 0.66 | 1457.32 | 0.51 |
| **a0014.C** | -0.02 | 0.05 | -0.39 | 2896.15 | 0.70 |
| **a0014^4** | -0.01 | 0.04 | -0.20 | 2729.22 | 0.84 |
| **dv_par_edu_birth** | -0.16 | 0.40 | -0.39 | 4169.70 | 0.70 |
| **dv_sc_age_5.L** | 0.17 | 0.07 | 2.28 | 1263.12 | 0.02 |
| **dv_sc_age_5.Q** | -0.04 | 0.04 | -1.11 | 1883.36 | 0.27 |
| **e216aNone** | 0.06 | 0.05 | 1.30 | 1849.05 | 0.19 |
| **dv_cog_abil_5** | -0.15 | 0.25 | -0.59 | 1100.46 | 0.55 |
| **dv_med_51** | 0.04 | 0.05 | 0.88 | 1976.16 | 0.38 |
| **d016a** | 0.09 | 0.04 | 2.05 | 2427.05 | 0.04 |
| **sepmumbcs1** | -0.04 | 0.13 | -0.28 | 1685.84 | 0.78 |
| **crowdUp to 1** | 0.01 | 0.06 | 0.18 | 1746.97 | 0.86 |
| **brfed.L** | 0.02 | 0.04 | 0.35 | 1596.09 | 0.72 |
| **brfed.Q** | 0.04 | 0.05 | 0.90 | 2219.26 | 0.37 |
| **resmove.L** | 0.02 | 0.04 | 0.41 | 3102.85 | 0.68 |
| **resmove.Q** | -0.06 | 0.03 | -1.99 | 3369.58 | 0.05 |

*Table S51.* Raw output from modified Poisson regression (with log link) of Rutter score at age 10 (BD3MRUTT) against median sleep duration (binarised to normal vs abnormal) derived from the van der Berg et al. algorithm at age 46, adjusted for potential confounders (see main text Table 3) and mental health variables at age 42 (see Table S42). The parameter estimates in the “Estimate” column and the associated standard errors have not been exponentiated.

| Term | Estimate | Standard error | Statistic | Degrees of freedom | p-value |
|---|---|---|---|---|---|
| **(Intercept)** | -0.96 | 0.30 | -3.18 | 1654.16 | 0.00 |
| **BD3MRUTT** | 0.02 | 0.02 | 0.97 | 2239.54 | 0.33 |
| **BD9MAL** | 0.28 | 0.13 | 2.06 | 1360.75 | 0.04 |
| **BD9WEMWB** | -0.12 | 0.23 | -0.51 | 1210.77 | 0.61 |
| **a0005a** | 0.12 | 0.20 | 0.62 | 1265.46 | 0.53 |
| **a0043b.L** | -0.04 | 0.04 | -0.94 | 2705.64 | 0.35 |
| **a0043b.Q** | 0.01 | 0.06 | 0.21 | 2107.63 | 0.83 |
| **a0043b.C** | 0.06 | 0.04 | 1.56 | 4652.36 | 0.12 |
| **a0043b^4** | 0.03 | 0.05 | 0.56 | 2560.57 | 0.58 |
| **a0043b^5** | 0.04 | 0.07 | 0.54 | 2509.35 | 0.59 |
| **a0195a** | -0.10 | 0.34 | -0.31 | 3657.88 | 0.76 |
| **a0278** | -0.05 | 0.28 | -0.18 | 1503.16 | 0.86 |
| **a0014.L** | 0.08 | 0.08 | 0.91 | 1676.78 | 0.36 |
| **a0014.Q** | 0.00 | 0.07 | 0.02 | 1423.78 | 0.98 |
| **a0014.C** | -0.04 | 0.09 | -0.43 | 1098.10 | 0.67 |
| **a0014^4** | 0.04 | 0.05 | 0.71 | 1582.18 | 0.47 |
| **dv_par_edu_birth** | 0.43 | 0.44 | 0.98 | 1813.50 | 0.33 |
| **dv_sc_age_5.L** | 0.01 | 0.08 | 0.07 | 1047.02 | 0.94 |
| **dv_sc_age_5.Q** | -0.01 | 0.05 | -0.21 | 1468.61 | 0.83 |
| **e216aNone** | -0.13 | 0.05 | -2.31 | 1345.22 | 0.02 |
| **dv_cog_abil_5** | -0.30 | 0.25 | -1.19 | 1006.33 | 0.23 |
| **dv_med_51** | 0.03 | 0.04 | 0.78 | 2553.97 | 0.43 |
| **d016a** | 0.19 | 0.05 | 3.51 | 1429.23 | 0.00 |
| **sepmumbcs1** | 0.20 | 0.10 | 2.09 | 1845.28 | 0.04 |
| **crowdUp to 1** | 0.06 | 0.05 | 1.24 | 2633.50 | 0.21 |
| **brfed.L** | 0.04 | 0.04 | 1.01 | 2185.86 | 0.31 |
| **brfed.Q** | -0.05 | 0.05 | -0.88 | 1600.89 | 0.38 |
| **resmove.L** | 0.03 | 0.06 | 0.46 | 1436.51 | 0.64 |
| **resmove.Q** | 0.02 | 0.02 | 1.07 | 5413.72 | 0.29 |

*Table S52.* Raw output from modified Poisson regression (with log link) of Rutter score at age 10 (BD3MRUTT) against median sleep duration (binarised to normal vs abnormal) derived from the Winkler et al. algorithm at age 46, adjusted for potential confounders (see main text Table 3) and mental health variables at age 42 (see Table S42). The parameter estimates in the “Estimate” column and the associated standard errors have not been exponentiated.

| Term | Estimate | Standard error | Statistic | Degrees of freedom | p-value |
|---|---|---|---|---|---|
| **(Intercept)** | -0.63 | 0.41 | -1.52 | 2467.96 | 0.13 |
| **dv_cds_10** | 0.12 | 0.03 | 3.71 | 1975.76 | 0.00 |
| **BD9MAL** | 1.02 | 0.15 | 6.92 | 2187.11 | 0.00 |
| **BD9WEMWB** | -1.08 | 0.22 | -4.90 | 2861.16 | 0.00 |
| **a0005a** | -0.27 | 0.15 | -1.85 | 9157.55 | 0.07 |
| **a0043b.L** | 0.15 | 0.06 | 2.45 | 3232.10 | 0.01 |
| **a0043b.Q** | 0.06 | 0.09 | 0.71 | 2713.63 | 0.48 |
| **a0043b.C** | 0.02 | 0.08 | 0.24 | 2968.21 | 0.81 |
| **a0043b^4** | -0.02 | 0.11 | -0.14 | 2012.56 | 0.89 |
| **a0043b^5** | -0.06 | 0.11 | -0.55 | 2894.27 | 0.58 |
| **a0195a** | -0.13 | 0.50 | -0.25 | 4148.09 | 0.80 |
| **a0278** | -0.55 | 0.24 | -2.29 | 8143.25 | 0.02 |
| **a0014.L** | 0.16 | 0.11 | 1.46 | 2616.62 | 0.14 |
| **a0014.Q** | 0.00 | 0.06 | -0.01 | 6307.43 | 0.99 |
| **a0014.C** | 0.11 | 0.08 | 1.36 | 2933.89 | 0.17 |
| **a0014^4** | 0.05 | 0.07 | 0.77 | 2541.76 | 0.44 |
| **dv_par_edu_birth** | -1.10 | 1.23 | -0.90 | 2802.56 | 0.37 |
| **dv_sc_age_5.L** | 0.18 | 0.09 | 1.84 | 1630.02 | 0.07 |
| **dv_sc_age_5.Q** | -0.06 | 0.04 | -1.41 | 5502.48 | 0.16 |
| **e216aNone** | 0.00 | 0.06 | 0.03 | 2993.19 | 0.98 |
| **dv_cog_abil_5** | -0.54 | 0.30 | -1.83 | 1633.95 | 0.07 |
| **dv_med_51** | 0.03 | 0.06 | 0.51 | 4728.50 | 0.61 |
| **d016a** | 0.20 | 0.06 | 3.55 | 3497.97 | 0.00 |
| **sepmumbcs1** | 0.11 | 0.11 | 0.96 | 3842.71 | 0.34 |
| **crowdUp to 1** | -0.20 | 0.06 | -3.32 | 3381.69 | 0.00 |
| **brfed.L** | 0.09 | 0.05 | 1.78 | 3924.24 | 0.08 |
| **brfed.Q** | -0.04 | 0.06 | -0.75 | 3926.53 | 0.46 |
| **resmove.L** | 0.16 | 0.05 | 3.01 | 5118.61 | 0.00 |
| **resmove.Q** | -0.02 | 0.03 | -0.61 | 8989.54 | 0.54 |

*Table S53.* Raw output from modified Poisson regression (with log link) of Child Development Scale score at age 10 (dv_cds_10) against self-reported average sleep duration (binarised to normal vs abnormal) at age 46, adjusted for potential confounders (see main text Table 3) and mental health variables at age 42 (see Table S42). The parameter estimates in the “Estimate” column and the associated standard errors have not been exponentiated.

| Term | Estimate | Standard error | Statistic | Degrees of freedom | p-value |
|---|---|---|---|---|---|
| **(Intercept)** | -1.06 | 0.55 | -1.91 | 1548.55 | 0.06 |
| **dv_cds_10** | 0.13 | 0.03 | 3.64 | 1826.43 | 0.00 |
| **BD9MAL** | 0.42 | 0.13 | 3.16 | 3675.50 | 0.00 |
| **BD9WEMWB** | -0.94 | 0.24 | -3.91 | 2728.80 | 0.00 |
| **a0005a** | 0.46 | 0.22 | 2.15 | 2597.00 | 0.03 |
| **a0043b.L** | 0.16 | 0.07 | 2.17 | 2454.08 | 0.03 |
| **a0043b.Q** | 0.17 | 0.10 | 1.66 | 2403.64 | 0.10 |
| **a0043b.C** | 0.04 | 0.09 | 0.39 | 2353.03 | 0.70 |
| **a0043b^4** | -0.10 | 0.10 | -1.03 | 2781.33 | 0.30 |
| **a0043b^5** | -0.06 | 0.12 | -0.56 | 3329.30 | 0.58 |
| **a0195a** | -0.01 | 0.65 | -0.02 | 2540.26 | 0.98 |
| **a0278** | 0.19 | 0.27 | 0.71 | 5200.16 | 0.48 |
| **a0014.L** | 0.24 | 0.14 | 1.71 | 1684.17 | 0.09 |
| **a0014.Q** | 0.06 | 0.08 | 0.74 | 2517.44 | 0.46 |
| **a0014.C** | 0.15 | 0.09 | 1.79 | 2746.69 | 0.07 |
| **a0014^4** | 0.09 | 0.06 | 1.55 | 3914.58 | 0.12 |
| **dv_par_edu_birth** | 0.01 | 0.96 | 0.01 | 2616.01 | 0.99 |
| **dv_sc_age_5.L** | 0.13 | 0.11 | 1.16 | 1381.47 | 0.24 |
| **dv_sc_age_5.Q** | 0.00 | 0.05 | -0.01 | 3520.85 | 0.99 |
| **e216aNone** | 0.10 | 0.07 | 1.36 | 2215.56 | 0.17 |
| **dv_cog_abil_5** | -1.41 | 0.37 | -3.79 | 1291.00 | 0.00 |
| **dv_med_51** | -0.02 | 0.10 | -0.18 | 1478.49 | 0.86 |
| **d016a** | 0.11 | 0.07 | 1.63 | 2800.35 | 0.10 |
| **sepmumbcs1** | 0.31 | 0.13 | 2.48 | 2742.84 | 0.01 |
| **crowdUp to 1** | -0.14 | 0.06 | -2.21 | 3444.97 | 0.03 |
| **brfed.L** | 0.01 | 0.06 | 0.12 | 2166.81 | 0.91 |
| **brfed.Q** | -0.01 | 0.10 | -0.06 | 1480.10 | 0.95 |
| **resmove.L** | 0.21 | 0.08 | 2.60 | 1819.37 | 0.01 |
| **resmove.Q** | 0.06 | 0.04 | 1.43 | 3695.10 | 0.15 |

*Table S54.* Raw output from modified Poisson regression (with log link) of Child Development Scale score at age 10 (dv_cds_10) against median sleep duration (binarised to normal vs abnormal) derived from the sleep diary at age 46, adjusted for potential confounders (see main text Table 3) and mental health variables at age 42 (see Table S42). The parameter estimates in the “Estimate” column and the associated standard errors have not been exponentiated.

| Term | Estimate | Standard error | Statistic | Degrees of freedom | p-value |
|---|---|---|---|---|---|
| **(Intercept)** | -0.45 | 0.24 | -1.91 | 2381.54 | 0.06 |
| **dv_cds_10** | -0.02 | 0.02 | -1.00 | 2988.24 | 0.32 |
| **BD9MAL** | 0.15 | 0.10 | 1.55 | 2107.83 | 0.12 |
| **BD9WEMWB** | 0.04 | 0.18 | 0.21 | 1631.74 | 0.83 |
| **a0005a** | -0.37 | 0.10 | -3.89 | 5722.99 | 0.00 |
| **a0043b.L** | -0.04 | 0.05 | -0.74 | 1704.39 | 0.46 |
| **a0043b.Q** | -0.02 | 0.06 | -0.38 | 2121.81 | 0.70 |
| **a0043b.C** | 0.00 | 0.06 | 0.05 | 1636.99 | 0.96 |
| **a0043b^4** | -0.05 | 0.04 | -1.24 | 3597.77 | 0.22 |
| **a0043b^5** | -0.02 | 0.07 | -0.25 | 2206.73 | 0.80 |
| **a0195a** | -0.40 | 0.47 | -0.85 | 1646.43 | 0.40 |
| **a0278** | -0.21 | 0.21 | -1.02 | 2167.87 | 0.31 |
| **a0014.L** | 0.04 | 0.07 | 0.51 | 1961.55 | 0.61 |
| **a0014.Q** | 0.03 | 0.05 | 0.67 | 1948.13 | 0.50 |
| **a0014.C** | 0.01 | 0.04 | 0.19 | 2779.68 | 0.85 |
| **a0014^4** | -0.01 | 0.04 | -0.25 | 1970.36 | 0.80 |
| **dv_par_edu_birth** | -0.46 | 0.48 | -0.95 | 2645.44 | 0.34 |
| **dv_sc_age_5.L** | 0.00 | 0.05 | 0.03 | 1997.25 | 0.97 |
| **dv_sc_age_5.Q** | 0.00 | 0.03 | -0.07 | 3858.05 | 0.94 |
| **e216aNone** | -0.01 | 0.05 | -0.21 | 1533.22 | 0.83 |
| **dv_cog_abil_5** | -0.07 | 0.25 | -0.28 | 957.05 | 0.78 |
| **dv_med_51** | 0.01 | 0.04 | 0.31 | 2881.52 | 0.75 |
| **d016a** | 0.07 | 0.03 | 2.00 | 3679.25 | 0.05 |
| **sepmumbcs1** | -0.04 | 0.09 | -0.43 | 2558.19 | 0.66 |
| **crowdUp to 1** | -0.07 | 0.05 | -1.46 | 2162.41 | 0.14 |
| **brfed.L** | 0.03 | 0.03 | 0.99 | 2094.04 | 0.32 |
| **brfed.Q** | -0.02 | 0.03 | -0.46 | 3446.53 | 0.65 |
| **resmove.L** | -0.02 | 0.04 | -0.44 | 2717.11 | 0.66 |
| **resmove.Q** | 0.06 | 0.02 | 2.58 | 4186.50 | 0.01 |

*Table S55.* Raw output from modified Poisson regression (with log link) of Child Development Scale score at age 10 (dv_cds_10) against median sleep duration (binarised to normal vs abnormal) derived from the activPAL algorithm at age 46, adjusted for potential confounders (see main text Table 3) and mental health variables at age 42 (see Table S42). The parameter estimates in the “Estimate” column and the associated standard errors have not been exponentiated.

| Term | Estimate | Standard error | Statistic | Degrees of freedom | p-value |
|---|---|---|---|---|---|
| **(Intercept)** | -1.04 | 0.36 | -2.93 | 1463.72 | 0.00 |
| **dv_cds_10** | 0.01 | 0.03 | 0.44 | 1260.39 | 0.66 |
| **BD9MAL** | 0.18 | 0.12 | 1.55 | 1824.05 | 0.12 |
| **BD9WEMWB** | -0.17 | 0.19 | -0.86 | 1667.27 | 0.39 |
| **a0005a** | -0.07 | 0.17 | -0.40 | 1760.83 | 0.69 |
| **a0043b.L** | 0.01 | 0.06 | 0.09 | 1746.96 | 0.93 |
| **a0043b.Q** | -0.08 | 0.07 | -1.16 | 1837.55 | 0.24 |
| **a0043b.C** | -0.18 | 0.07 | -2.77 | 1699.23 | 0.01 |
| **a0043b^4** | 0.11 | 0.06 | 1.75 | 2057.38 | 0.08 |
| **a0043b^5** | 0.03 | 0.07 | 0.42 | 2212.02 | 0.67 |
| **a0195a** | -0.24 | 0.46 | -0.51 | 2056.90 | 0.61 |
| **a0278** | 0.32 | 0.17 | 1.82 | 4333.17 | 0.07 |
| **a0014.L** | 0.05 | 0.10 | 0.50 | 1400.08 | 0.62 |
| **a0014.Q** | 0.04 | 0.07 | 0.66 | 1440.82 | 0.51 |
| **a0014.C** | -0.02 | 0.05 | -0.39 | 2867.91 | 0.70 |
| **a0014^4** | -0.01 | 0.04 | -0.21 | 2686.29 | 0.83 |
| **dv_par_edu_birth** | -0.15 | 0.41 | -0.38 | 4045.87 | 0.71 |
| **dv_sc_age_5.L** | 0.17 | 0.07 | 2.34 | 1297.07 | 0.02 |
| **dv_sc_age_5.Q** | -0.04 | 0.04 | -1.11 | 1868.13 | 0.27 |
| **e216aNone** | 0.06 | 0.05 | 1.28 | 1837.38 | 0.20 |
| **dv_cog_abil_5** | -0.14 | 0.25 | -0.55 | 1090.44 | 0.58 |
| **dv_med_51** | 0.04 | 0.05 | 0.89 | 1966.03 | 0.37 |
| **d016a** | 0.09 | 0.04 | 2.03 | 2293.73 | 0.04 |
| **sepmumbcs1** | -0.04 | 0.13 | -0.29 | 1666.67 | 0.77 |
| **crowdUp to 1** | 0.01 | 0.06 | 0.17 | 1817.20 | 0.87 |
| **brfed.L** | 0.02 | 0.04 | 0.37 | 1605.08 | 0.71 |
| **brfed.Q** | 0.04 | 0.05 | 0.88 | 2207.90 | 0.38 |
| **resmove.L** | 0.02 | 0.04 | 0.43 | 3060.00 | 0.67 |
| **resmove.Q** | -0.05 | 0.03 | -1.98 | 3434.96 | 0.05 |

*Table S56.* Raw output from modified Poisson regression (with log link) of Child Development Scale score at age 10 (dv_cds_10) against median sleep duration (binarised to normal vs abnormal) derived from the van der Berg et al. algorithm at age 46, adjusted for potential confounders (see main text Table 3) and mental health variables at age 42 (see Table S42). The parameter estimates in the "Estimate" column and the associated standard errors have not been exponentiated.

| Term | Estimate | Standard error | Statistic | Degrees of freedom | p-value |
|---|---|---|---|---|---|
| **(Intercept)** | -1.07 | 0.31 | -3.41 | 1571.97 | 0.00 |
| **dv_cds_10** | 0.09 | 0.03 | 3.31 | 1194.95 | 0.00 |
| **BD9MAL** | 0.27 | 0.13 | 2.03 | 1373.62 | 0.04 |
| **BD9WEMWB** | -0.09 | 0.23 | -0.39 | 1204.81 | 0.70 |
| **a0005a** | 0.11 | 0.20 | 0.57 | 1257.94 | 0.57 |
| **a0043b.L** | -0.04 | 0.04 | -1.08 | 2801.81 | 0.28 |
| **a0043b.Q** | 0.01 | 0.06 | 0.22 | 2106.88 | 0.83 |
| **a0043b.C** | 0.06 | 0.04 | 1.42 | 4302.59 | 0.16 |
| **a0043b^4** | 0.03 | 0.05 | 0.57 | 2550.81 | 0.57 |
| **a0043b^5** | 0.04 | 0.07 | 0.59 | 2532.20 | 0.55 |
| **a0195a** | -0.07 | 0.34 | -0.19 | 3519.79 | 0.85 |
| **a0278** | -0.04 | 0.28 | -0.14 | 1482.15 | 0.89 |
| **a0014.L** | 0.07 | 0.08 | 0.89 | 1678.66 | 0.37 |
| **a0014.Q** | 0.00 | 0.07 | 0.04 | 1392.73 | 0.96 |
| **a0014.C** | -0.04 | 0.09 | -0.45 | 1099.57 | 0.65 |
| **a0014^4** | 0.03 | 0.05 | 0.62 | 1621.90 | 0.53 |
| **dv_par_edu_birth** | 0.47 | 0.44 | 1.07 | 1775.30 | 0.28 |
| **dv_sc_age_5.L** | 0.00 | 0.08 | -0.01 | 1041.92 | 0.99 |
| **dv_sc_age_5.Q** | -0.01 | 0.04 | -0.18 | 1469.25 | 0.86 |
| **e216aNone** | -0.13 | 0.05 | -2.34 | 1364.79 | 0.02 |
| **dv_cog_abil_5** | -0.19 | 0.25 | -0.74 | 1017.45 | 0.46 |
| **dv_med_51** | 0.03 | 0.04 | 0.76 | 2822.08 | 0.44 |
| **d016a** | 0.18 | 0.05 | 3.38 | 1444.18 | 0.00 |
| **sepmumbcs1** | 0.18 | 0.10 | 1.90 | 1845.71 | 0.06 |
| **crowdUp to 1** | 0.06 | 0.05 | 1.37 | 2629.68 | 0.17 |
| **brfed.L** | 0.04 | 0.04 | 1.05 | 2144.50 | 0.29 |
| **brfed.Q** | -0.04 | 0.05 | -0.87 | 1605.01 | 0.39 |
| **resmove.L** | 0.02 | 0.06 | 0.34 | 1466.60 | 0.73 |
| **resmove.Q** | 0.02 | 0.02 | 1.07 | 5178.93 | 0.29 |

*Table S57.* Raw output from modified Poisson regression (with log link) of Child Development Scale score at age 10 (dv_cds_10) against median sleep duration (binarised to normal vs abnormal) derived from the Winkler et al. algorithm at age 46, adjusted for potential confounders (see main text Table 3) and mental health variables at age 42 (see Table S42). The parameter estimates in the “Estimate” column and the associated standard errors have not been exponentiated.

| Term | Estimate | Standard error | Statistic | Degrees of freedom | p-value |
|---|---|---|---|---|---|
| **(Intercept)** | -0.40 | 0.45 | -0.89 | 2324.20 | 0.37 |
| **BD4MAL** | 0.06 | 0.04 | 1.74 | 1519.90 | 0.08 |
| **BD9MAL** | 0.94 | 0.15 | 6.38 | 2359.61 | 0.00 |
| **BD9WEMWB** | -1.09 | 0.23 | -4.79 | 2656.14 | 0.00 |
| **a0005a** | -0.22 | 0.15 | -1.44 | 8409.36 | 0.15 |
| **a0043b.L** | 0.14 | 0.06 | 2.17 | 3322.15 | 0.03 |
| **a0043b.Q** | 0.08 | 0.09 | 0.85 | 2603.06 | 0.40 |
| **a0043b.C** | 0.01 | 0.08 | 0.17 | 2672.03 | 0.86 |
| **a0043b^4** | -0.03 | 0.11 | -0.27 | 2006.02 | 0.78 |
| **a0043b^5** | -0.05 | 0.12 | -0.47 | 2786.16 | 0.64 |
| **a0195a** | -0.19 | 0.49 | -0.38 | 4475.29 | 0.70 |
| **a0278** | -0.53 | 0.24 | -2.17 | 7551.44 | 0.03 |
| **a0014.L** | 0.13 | 0.11 | 1.22 | 3095.20 | 0.22 |
| **a0014.Q** | -0.01 | 0.06 | -0.12 | 6532.90 | 0.90 |
| **a0014.C** | 0.12 | 0.08 | 1.53 | 3206.08 | 0.13 |
| **a0014^4** | 0.05 | 0.07 | 0.71 | 2594.38 | 0.48 |
| **dv_par_edu_birth** | -1.01 | 1.19 | -0.85 | 2887.72 | 0.39 |
| **dv_sc_age_5.L** | 0.14 | 0.10 | 1.38 | 1621.27 | 0.17 |
| **dv_sc_age_5.Q** | -0.05 | 0.04 | -1.07 | 4084.51 | 0.28 |
| **e216aNone** | 0.01 | 0.06 | 0.14 | 2853.18 | 0.89 |
| **dv_cog_abil_5** | -0.51 | 0.31 | -1.64 | 1634.72 | 0.10 |
| **dv_med_51** | 0.02 | 0.06 | 0.40 | 4015.90 | 0.69 |
| **d016a** | 0.20 | 0.05 | 3.87 | 4313.68 | 0.00 |
| **dv_bas_g** | -0.37 | 0.23 | -1.60 | 2327.38 | 0.11 |
| **dv_med_101** | -0.01 | 0.04 | -0.18 | 6691.34 | 0.86 |
| **bmi** | -0.04 | 0.25 | -0.16 | 3550.82 | 0.87 |
| **fclrg90.L** | 0.00 | 0.09 | 0.04 | 3330.42 | 0.96 |
| **fclrg90.Q** | 0.02 | 0.06 | 0.36 | 5650.77 | 0.72 |
| **fclrg90.C** | -0.08 | 0.08 | -1.06 | 3501.85 | 0.29 |
| **fclrg90^4** | 0.02 | 0.05 | 0.45 | 6013.62 | 0.66 |
| **tenure.L** | 0.11 | 0.05 | 2.09 | 2894.62 | 0.04 |
| **tenure.Q** | -0.06 | 0.07 | -0.99 | 3577.01 | 0.32 |
| **divorce1** | 0.06 | 0.06 | 1.05 | 4585.18 | 0.29 |
| **sepmumbcs1** | 0.09 | 0.11 | 0.85 | 4981.26 | 0.40 |
| **prmnh1** | 0.15 | 0.12 | 1.24 | 2310.59 | 0.22 |
| **crowdUp to 1** | -0.15 | 0.06 | -2.78 | 4990.21 | 0.01 |
| **ameniNo occasions** | -0.06 | 0.12 | -0.48 | 2381.20 | 0.63 |
| **brfed.L** | 0.08 | 0.05 | 1.46 | 3793.21 | 0.14 |
| **brfed.Q** | -0.04 | 0.06 | -0.76 | 3934.43 | 0.44 |
| **resmove.L** | 0.14 | 0.06 | 2.43 | 4015.56 | 0.01 |
| **resmove.Q** | -0.02 | 0.03 | -0.63 | 8521.47 | 0.53 |

*Table S58.* Raw output from modified Poisson regression (with log link) of Malaise Inventory score at age 16 (BD4MAL) against self-reported average sleep duration (binarised to normal vs abnormal) at age 46, adjusted for potential confounders (see main text Table 3) and mental health variables at age 42 (see Table S42). The parameter estimates in the "Estimate" column and the associated standard errors have not been exponentiated.

| Term | Estimate | Standard error | Statistic | Degrees of freedom | p-value |
|---|---|---|---|---|---|
| **(Intercept)** | -0.69 | 0.51 | -1.35 | 1920.42 | 0.18 |
| **BD4MAL** | 0.01 | 0.04 | 0.14 | 1737.64 | 0.89 |
| **BD9MAL** | 0.37 | 0.11 | 3.19 | 6770.06 | 0.00 |
| **BD9WEMWB** | -0.99 | 0.24 | -4.22 | 2910.46 | 0.00 |
| **a0005a** | 0.48 | 0.22 | 2.21 | 2624.88 | 0.03 |
| **a0043b.L** | 0.15 | 0.07 | 2.05 | 2549.45 | 0.04 |
| **a0043b.Q** | 0.17 | 0.10 | 1.70 | 2517.22 | 0.09 |
| **a0043b.C** | 0.04 | 0.10 | 0.39 | 2153.01 | 0.69 |
| **a0043b^4** | -0.10 | 0.09 | -1.10 | 2906.16 | 0.27 |
| **a0043b^5** | -0.07 | 0.12 | -0.56 | 3178.92 | 0.58 |
| **a0195a** | -0.09 | 0.65 | -0.14 | 2518.79 | 0.89 |
| **a0278** | 0.27 | 0.29 | 0.94 | 4538.95 | 0.35 |
| **a0014.L** | 0.16 | 0.14 | 1.14 | 1733.00 | 0.26 |
| **a0014.Q** | 0.05 | 0.08 | 0.67 | 2888.24 | 0.50 |
| **a0014.C** | 0.16 | 0.09 | 1.72 | 2446.98 | 0.09 |
| **a0014^4** | 0.09 | 0.06 | 1.55 | 4043.06 | 0.12 |
| **dv_par_edu_birth** | 0.16 | 0.89 | 0.18 | 2735.17 | 0.86 |
| **dv_sc_age_5.L** | 0.06 | 0.13 | 0.50 | 1269.11 | 0.62 |
| **dv_sc_age_5.Q** | 0.01 | 0.05 | 0.28 | 4279.59 | 0.78 |
| **e216aNone** | 0.10 | 0.07 | 1.36 | 2307.61 | 0.18 |
| **dv_cog_abil_5** | -1.35 | 0.42 | -3.19 | 1156.39 | 0.00 |
| **dv_med_51** | -0.03 | 0.10 | -0.25 | 1448.93 | 0.80 |
| **d016a** | 0.11 | 0.07 | 1.68 | 2633.51 | 0.09 |
| **dv_bas_g** | -0.45 | 0.31 | -1.48 | 1624.31 | 0.14 |
| **dv_med_101** | 0.04 | 0.06 | 0.63 | 3007.04 | 0.53 |
| **bmi** | -0.50 | 0.28 | -1.80 | 3151.03 | 0.07 |
| **fclrg90.L** | 0.21 | 0.10 | 2.19 | 2929.51 | 0.03 |
| **fclrg90.Q** | 0.04 | 0.07 | 0.66 | 3549.82 | 0.51 |
| **fclrg90.C** | -0.04 | 0.11 | -0.42 | 1813.40 | 0.67 |
| **fclrg90^4** | -0.02 | 0.08 | -0.25 | 1947.28 | 0.80 |
| **tenure.L** | 0.05 | 0.07 | 0.75 | 1795.39 | 0.45 |
| **tenure.Q** | -0.05 | 0.09 | -0.54 | 1975.13 | 0.59 |
| **divorce1** | 0.03 | 0.07 | 0.42 | 3673.76 | 0.68 |
| **sepmumbcs1** | 0.33 | 0.14 | 2.40 | 2544.68 | 0.02 |
| **prmnh1** | 0.34 | 0.14 | 2.40 | 1847.19 | 0.02 |
| **crowdUp to 1** | -0.10 | 0.07 | -1.59 | 3509.76 | 0.11 |
| **ameniNo occasions** | 0.01 | 0.11 | 0.13 | 4047.44 | 0.90 |
| **brfed.L** | -0.01 | 0.06 | -0.14 | 2222.43 | 0.89 |
| **brfed.Q** | 0.00 | 0.10 | -0.03 | 1386.62 | 0.97 |
| **resmove.L** | 0.21 | 0.09 | 2.44 | 1737.60 | 0.01 |
| **resmove.Q** | 0.06 | 0.04 | 1.41 | 3579.83 | 0.16 |

*Table S59.* Raw output from modified Poisson regression (with log link) of Malaise Inventory score at age 16 (BD4MAL) against median sleep duration (binarised to normal vs abnormal) derived from the sleep diary at age 46, adjusted for potential confounders (see main text Table 3) and mental health variables at age 42 (see Table S42). The parameter estimates in the “Estimate” column and the associated standard errors have not been exponentiated.

| Term | Estimate | Standard error | Statistic | Degrees of freedom | p-value |
|---|---|---|---|---|---|
| **(Intercept)** | -0.42 | 0.25 | -1.66 | 2345.22 | 0.10 |
| **BD4MAL** | 0.00 | 0.02 | 0.25 | 1914.06 | 0.80 |
| **BD9MAL** | 0.12 | 0.09 | 1.27 | 2356.80 | 0.20 |
| **BD9WEMWB** | 0.05 | 0.18 | 0.29 | 1634.99 | 0.77 |
| **a0005a** | -0.35 | 0.10 | -3.56 | 4908.31 | 0.00 |
| **a0043b.L** | -0.05 | 0.05 | -1.02 | 1776.05 | 0.31 |
| **a0043b.Q** | -0.02 | 0.06 | -0.39 | 2100.73 | 0.70 |
| **a0043b.C** | 0.00 | 0.06 | -0.02 | 1672.60 | 0.98 |
| **a0043b^4** | -0.06 | 0.04 | -1.32 | 3654.36 | 0.19 |
| **a0043b^5** | -0.01 | 0.07 | -0.17 | 2101.53 | 0.86 |
| **a0195a** | -0.39 | 0.46 | -0.83 | 1666.99 | 0.40 |
| **a0278** | -0.20 | 0.20 | -1.02 | 2397.58 | 0.31 |
| **a0014.L** | 0.01 | 0.07 | 0.20 | 1948.68 | 0.84 |
| **a0014.Q** | 0.03 | 0.05 | 0.57 | 1979.66 | 0.57 |
| **a0014.C** | 0.00 | 0.05 | 0.06 | 2575.81 | 0.95 |
| **a0014^4** | -0.01 | 0.04 | -0.26 | 1759.97 | 0.79 |
| **dv_par_edu_birth** | -0.38 | 0.46 | -0.83 | 2684.72 | 0.41 |
| **dv_sc_age_5.L** | -0.02 | 0.06 | -0.26 | 1498.89 | 0.79 |
| **dv_sc_age_5.Q** | 0.00 | 0.03 | -0.16 | 2920.73 | 0.87 |
| **e216aNone** | -0.01 | 0.05 | -0.12 | 1539.83 | 0.90 |
| **dv_cog_abil_5** | 0.04 | 0.28 | 0.15 | 863.33 | 0.88 |
| **dv_med_51** | 0.00 | 0.04 | 0.12 | 2958.76 | 0.91 |
| **d016a** | 0.06 | 0.03 | 1.70 | 3473.91 | 0.09 |
| **dv_bas_g** | -0.22 | 0.15 | -1.41 | 1852.52 | 0.16 |
| **dv_med_101** | 0.01 | 0.03 | 0.27 | 2886.59 | 0.79 |
| **bmi** | 0.19 | 0.17 | 1.08 | 2171.61 | 0.28 |
| **fclrg90.L** | 0.02 | 0.06 | 0.39 | 2489.46 | 0.70 |
| **fclrg90.Q** | 0.05 | 0.04 | 1.04 | 2465.91 | 0.30 |
| **fclrg90.C** | 0.06 | 0.04 | 1.52 | 3982.99 | 0.13 |
| **fclrg90^4** | 0.01 | 0.05 | 0.27 | 1775.74 | 0.79 |
| **tenure.L** | 0.01 | 0.03 | 0.46 | 2429.25 | 0.64 |
| **tenure.Q** | -0.03 | 0.05 | -0.70 | 2277.40 | 0.48 |
| **divorce1** | 0.03 | 0.04 | 0.73 | 4015.22 | 0.47 |
| **sepmumbcs1** | -0.06 | 0.09 | -0.70 | 3004.60 | 0.48 |
| **prmnh1** | 0.17 | 0.10 | 1.80 | 1741.33 | 0.07 |
| **crowdUp to 1** | -0.05 | 0.05 | -0.88 | 1848.76 | 0.38 |
| **ameniNo occasions** | -0.08 | 0.08 | -1.06 | 2136.76 | 0.29 |
| **brfed.L** | 0.03 | 0.03 | 0.82 | 2105.10 | 0.41 |
| **brfed.Q** | -0.01 | 0.03 | -0.40 | 3442.13 | 0.69 |
| **resmove.L** | -0.03 | 0.04 | -0.74 | 2834.89 | 0.46 |
| **resmove.Q** | 0.06 | 0.02 | 2.50 | 4232.59 | 0.01 |

*Table S60.* Raw output from modified Poisson regression (with log link) of Malaise Inventory score at age 16 (BD4MAL) against median sleep duration (binarised to normal vs abnormal) derived from the activPAL algorithm at age 46, adjusted for potential confounders (see main text Table 3) and mental health variables at age 42 (see Table S42). The parameter estimates in the “Estimate” column and the associated standard errors have not been exponentiated.

| Term | Estimate | Standard error | Statistic | Degrees of freedom | p-value |
|---|---|---|---|---|---|
| **(Intercept)** | -0.86 | 0.36 | -2.40 | 1572.59 | 0.02 |
| **BD4MAL** | -0.01 | 0.03 | -0.56 | 1501.76 | 0.57 |
| **BD9MAL** | 0.19 | 0.12 | 1.56 | 1749.40 | 0.12 |
| **BD9WEMWB** | -0.15 | 0.18 | -0.83 | 1807.03 | 0.41 |
| **a0005a** | -0.01 | 0.17 | -0.09 | 1756.57 | 0.93 |
| **a0043b.L** | 0.00 | 0.06 | 0.01 | 1595.23 | 0.99 |
| **a0043b.Q** | -0.07 | 0.07 | -0.99 | 1783.98 | 0.32 |
| **a0043b.C** | -0.18 | 0.07 | -2.78 | 1701.34 | 0.01 |
| **a0043b^4** | 0.10 | 0.06 | 1.64 | 2033.84 | 0.10 |
| **a0043b^5** | 0.03 | 0.07 | 0.43 | 2182.89 | 0.66 |
| **a0195a** | -0.28 | 0.47 | -0.59 | 1997.84 | 0.56 |
| **a0278** | 0.36 | 0.18 | 2.00 | 3879.25 | 0.05 |
| **a0014.L** | 0.03 | 0.09 | 0.31 | 1544.61 | 0.75 |
| **a0014.Q** | 0.04 | 0.07 | 0.62 | 1542.75 | 0.54 |
| **a0014.C** | -0.01 | 0.05 | -0.16 | 2652.28 | 0.87 |
| **a0014^4** | -0.02 | 0.04 | -0.46 | 2363.14 | 0.65 |
| **dv_par_edu_birth** | -0.11 | 0.41 | -0.26 | 3809.88 | 0.80 |
| **dv_sc_age_5.L** | 0.15 | 0.08 | 1.85 | 1176.68 | 0.07 |
| **dv_sc_age_5.Q** | -0.03 | 0.04 | -0.94 | 2194.20 | 0.35 |
| **e216aNone** | 0.07 | 0.05 | 1.40 | 1903.74 | 0.16 |
| **dv_cog_abil_5** | 0.00 | 0.27 | 0.01 | 1040.01 | 0.99 |
| **dv_med_51** | 0.04 | 0.05 | 0.82 | 1871.66 | 0.41 |
| **d016a** | 0.09 | 0.04 | 2.00 | 2412.97 | 0.05 |
| **dv_bas_g** | -0.38 | 0.22 | -1.74 | 1267.16 | 0.08 |
| **dv_med_101** | 0.04 | 0.05 | 0.78 | 1500.85 | 0.43 |
| **bmi** | -0.26 | 0.24 | -1.08 | 1568.12 | 0.28 |
| **fclrg90.L** | -0.02 | 0.06 | -0.24 | 2729.17 | 0.81 |
| **fclrg90.Q** | -0.03 | 0.06 | -0.46 | 1675.58 | 0.64 |
| **fclrg90.C** | -0.10 | 0.06 | -1.79 | 2162.68 | 0.07 |
| **fclrg90^4** | 0.01 | 0.05 | 0.22 | 2080.47 | 0.83 |
| **tenure.L** | 0.03 | 0.05 | 0.65 | 1382.08 | 0.52 |
| **tenure.Q** | 0.09 | 0.06 | 1.59 | 2225.47 | 0.11 |
| **divorce1** | 0.14 | 0.05 | 3.12 | 2946.47 | 0.00 |
| **sepmumbcs1** | -0.11 | 0.14 | -0.81 | 1572.54 | 0.42 |
| **prmnh1** | -0.22 | 0.18 | -1.20 | 1127.46 | 0.23 |
| **crowdUp to 1** | 0.03 | 0.06 | 0.44 | 1835.24 | 0.66 |
| **ameniNo occasions** | -0.08 | 0.08 | -1.09 | 2484.87 | 0.28 |
| **brfed.L** | 0.01 | 0.04 | 0.23 | 1629.27 | 0.82 |
| **brfed.Q** | 0.04 | 0.05 | 0.92 | 2245.13 | 0.36 |
| **resmove.L** | 0.00 | 0.04 | 0.08 | 3452.72 | 0.93 |
| **resmove.Q** | -0.06 | 0.03 | -2.14 | 3541.08 | 0.03 |

*Table S61.* Raw output from modified Poisson regression (with log link) of Malaise Inventory score at age 16 (BD4MAL) against median sleep duration (binarised to normal vs abnormal) derived from the van der Berg et al. algorithm at age 46, adjusted for potential confounders (see main text Table 3) and mental health variables at age 42 (see Table S42). The parameter estimates in the “Estimate” column and the associated standard errors have not been exponentiated.

| Term | Estimate | Standard error | Statistic | Degrees of freedom | p-value |
|---|---|---|---|---|---|
| **(Intercept)** | -0.91 | 0.37 | -2.48 | 1366.65 | 0.01 |
| **BD4MAL** | 0.04 | 0.02 | 1.99 | 1722.49 | 0.05 |
| **BD9MAL** | 0.23 | 0.14 | 1.67 | 1384.30 | 0.10 |
| **BD9WEMWB** | -0.10 | 0.24 | -0.43 | 1177.25 | 0.66 |
| **a0005a** | 0.15 | 0.20 | 0.72 | 1252.15 | 0.47 |
| **a0043b.L** | -0.05 | 0.04 | -1.23 | 2997.28 | 0.22 |
| **a0043b.Q** | 0.02 | 0.06 | 0.37 | 2143.06 | 0.71 |
| **a0043b.C** | 0.06 | 0.04 | 1.43 | 4349.15 | 0.15 |
| **a0043b^4** | 0.02 | 0.05 | 0.46 | 2541.34 | 0.64 |
| **a0043b^5** | 0.05 | 0.07 | 0.67 | 2587.53 | 0.50 |
| **a0195a** | -0.08 | 0.33 | -0.24 | 3890.87 | 0.81 |
| **a0278** | -0.08 | 0.27 | -0.31 | 1601.22 | 0.76 |
| **a0014.L** | 0.08 | 0.08 | 1.02 | 2091.42 | 0.31 |
| **a0014.Q** | 0.01 | 0.06 | 0.18 | 1536.58 | 0.86 |
| **a0014.C** | -0.04 | 0.09 | -0.42 | 1088.33 | 0.67 |
| **a0014^4** | 0.03 | 0.05 | 0.62 | 1550.83 | 0.54 |
| **dv_par_edu_birth** | 0.47 | 0.42 | 1.12 | 1835.00 | 0.26 |
| **dv_sc_age_5.L** | 0.01 | 0.09 | 0.14 | 998.98 | 0.89 |
| **dv_sc_age_5.Q** | -0.01 | 0.05 | -0.16 | 1420.84 | 0.87 |
| **e216aNone** | -0.12 | 0.05 | -2.29 | 1396.77 | 0.02 |
| **dv_cog_abil_5** | -0.24 | 0.27 | -0.89 | 988.69 | 0.37 |
| **dv_med_51** | 0.02 | 0.04 | 0.58 | 2570.30 | 0.57 |
| **d016a** | 0.19 | 0.05 | 3.52 | 1425.43 | 0.00 |
| **dv_bas_g** | -0.23 | 0.18 | -1.26 | 1580.23 | 0.21 |
| **dv_med_101** | 0.03 | 0.05 | 0.58 | 1553.85 | 0.56 |
| **bmi** | 0.52 | 0.20 | 2.54 | 1686.34 | 0.01 |
| **fclrg90.L** | -0.12 | 0.08 | -1.46 | 1558.19 | 0.15 |
| **fclrg90.Q** | -0.05 | 0.06 | -0.93 | 1776.50 | 0.35 |
| **fclrg90.C** | 0.03 | 0.07 | 0.38 | 1490.12 | 0.70 |
| **fclrg90^4** | 0.01 | 0.05 | 0.12 | 1738.82 | 0.91 |
| **tenure.L** | 0.04 | 0.04 | 0.81 | 1440.83 | 0.42 |
| **tenure.Q** | -0.05 | 0.06 | -0.80 | 1641.49 | 0.43 |
| **divorce1** | -0.02 | 0.07 | -0.33 | 1480.27 | 0.74 |
| **sepmumbcs1** | 0.21 | 0.10 | 2.13 | 1993.15 | 0.03 |
| **prmnh1** | -0.07 | 0.11 | -0.66 | 2064.93 | 0.51 |
| **crowdUp to 1** | 0.08 | 0.05 | 1.46 | 2056.63 | 0.14 |
| **ameniNo occasions** | -0.16 | 0.09 | -1.78 | 1564.38 | 0.08 |
| **brfed.L** | 0.03 | 0.03 | 0.91 | 2292.60 | 0.36 |
| **brfed.Q** | -0.05 | 0.05 | -0.89 | 1581.83 | 0.37 |
| **resmove.L** | 0.02 | 0.06 | 0.37 | 1511.49 | 0.71 |
| **resmove.Q** | 0.03 | 0.02 | 1.10 | 4863.58 | 0.27 |

*Table S62.* Raw output from modified Poisson regression (with log link) of Malaise Inventory score at age 16 (BD4MAL) against median sleep duration (binarised to normal vs abnormal) derived from the Winkler et al. algorithm at age 46, adjusted for potential confounders (see main text Table 3) and mental health variables at age 42 (see Table S42). The parameter estimates in the "Estimate" column and the associated standard errors have not been exponentiated.

| Term | Estimate | Standard error | Statistic | Degrees of freedom | p-value |
|---|---|---|---|---|---|
| **(Intercept)** | -0.40 | 0.44 | -0.90 | 2391.09 | 0.37 |
| **rd6m_1** | 0.05 | 0.02 | 2.88 | 2606.60 | 0.00 |
| **BD9MAL** | 0.99 | 0.14 | 6.87 | 2309.02 | 0.00 |
| **BD9WEMWB** | -1.10 | 0.22 | -4.92 | 2802.30 | 0.00 |
| **a0005a** | -0.22 | 0.15 | -1.47 | 8315.03 | 0.14 |
| **a0043b.L** | 0.14 | 0.06 | 2.21 | 3248.05 | 0.03 |
| **a0043b.Q** | 0.07 | 0.09 | 0.81 | 2752.22 | 0.42 |
| **a0043b.C** | 0.02 | 0.08 | 0.23 | 2865.65 | 0.81 |
| **a0043b^4** | -0.03 | 0.11 | -0.25 | 2007.51 | 0.81 |
| **a0043b^5** | -0.06 | 0.11 | -0.56 | 2862.10 | 0.57 |
| **a0195a** | -0.21 | 0.49 | -0.43 | 4417.21 | 0.67 |
| **a0278** | -0.54 | 0.25 | -2.18 | 7292.80 | 0.03 |
| **a0014.L** | 0.12 | 0.10 | 1.20 | 3211.33 | 0.23 |
| **a0014.Q** | -0.01 | 0.06 | -0.19 | 6991.39 | 0.85 |
| **a0014.C** | 0.12 | 0.08 | 1.48 | 3082.92 | 0.14 |
| **a0014^4** | 0.05 | 0.07 | 0.74 | 2627.29 | 0.46 |
| **dv_par_edu_birth** | -1.07 | 1.21 | -0.88 | 2893.35 | 0.38 |
| **dv_sc_age_5.L** | 0.15 | 0.10 | 1.40 | 1581.17 | 0.16 |
| **dv_sc_age_5.Q** | -0.05 | 0.04 | -1.10 | 4176.52 | 0.27 |
| **e216aNone** | 0.01 | 0.06 | 0.15 | 2925.64 | 0.88 |
| **dv_cog_abil_5** | -0.51 | 0.30 | -1.68 | 1688.71 | 0.09 |
| **dv_med_51** | 0.02 | 0.06 | 0.41 | 4230.11 | 0.68 |
| **d016a** | 0.20 | 0.05 | 3.71 | 4315.16 | 0.00 |
| **dv_bas_g** | -0.36 | 0.23 | -1.59 | 2434.24 | 0.11 |
| **dv_med_101** | -0.01 | 0.05 | -0.14 | 6025.56 | 0.89 |
| **bmi** | -0.04 | 0.24 | -0.17 | 3765.07 | 0.86 |
| **fclrg90.L** | 0.00 | 0.09 | 0.01 | 3275.74 | 0.99 |
| **fclrg90.Q** | 0.02 | 0.06 | 0.42 | 5397.13 | 0.67 |
| **fclrg90.C** | -0.08 | 0.08 | -1.08 | 3613.83 | 0.28 |
| **fclrg90^4** | 0.03 | 0.05 | 0.49 | 5781.35 | 0.62 |
| **tenure.L** | 0.10 | 0.05 | 2.10 | 2976.04 | 0.04 |
| **tenure.Q** | -0.06 | 0.06 | -1.02 | 3851.56 | 0.31 |
| **divorce1** | 0.07 | 0.06 | 1.12 | 4252.16 | 0.26 |
| **sepmumbcs1** | 0.07 | 0.11 | 0.59 | 4936.28 | 0.55 |
| **prmnh1** | 0.18 | 0.13 | 1.32 | 1979.91 | 0.19 |
| **crowdUp to 1** | -0.16 | 0.06 | -2.85 | 4671.09 | 0.00 |
| **ameniNo occasions** | -0.04 | 0.11 | -0.33 | 2662.46 | 0.74 |
| **brfed.L** | 0.07 | 0.05 | 1.42 | 3866.01 | 0.16 |
| **brfed.Q** | -0.05 | 0.06 | -0.83 | 3815.04 | 0.41 |
| **resmove.L** | 0.14 | 0.06 | 2.54 | 4331.82 | 0.01 |
| **resmove.Q** | -0.02 | 0.03 | -0.63 | 8501.88 | 0.53 |

*Table S63.* Raw output from modified Poisson regression (with log link) of behavioural and emotional problems at age 16 (rd6m_1) against self-reported average sleep duration (binarised to normal vs abnormal) at age 46, adjusted for potential confounders (see main text Table 3) and mental health variables at age 42 (see Table S42). The parameter estimates in the “Estimate” column and the associated standard errors have not been exponentiated.

| Term | Estimate | Standard error | Statistic | Degrees of freedom | p-value |
|---|---|---|---|---|---|
| **(Intercept)** | -0.70 | 0.50 | -1.40 | 1950.75 | 0.16 |
| **rd6m_1** | 0.05 | 0.03 | 1.68 | 1319.22 | 0.09 |
| **BD9MAL** | 0.36 | 0.13 | 2.81 | 4275.96 | 0.00 |
| **BD9WEMWB** | -0.98 | 0.22 | -4.40 | 3322.81 | 0.00 |
| **a0005a** | 0.47 | 0.21 | 2.20 | 2640.09 | 0.03 |
| **a0043b.L** | 0.15 | 0.07 | 2.17 | 2673.07 | 0.03 |
| **a0043b.Q** | 0.17 | 0.10 | 1.73 | 2576.10 | 0.08 |
| **a0043b.C** | 0.04 | 0.10 | 0.41 | 2173.02 | 0.68 |
| **a0043b^4** | -0.10 | 0.09 | -1.14 | 3049.77 | 0.26 |
| **a0043b^5** | -0.07 | 0.12 | -0.58 | 3082.57 | 0.56 |
| **a0195a** | -0.11 | 0.65 | -0.17 | 2489.80 | 0.87 |
| **a0278** | 0.28 | 0.28 | 0.98 | 4658.24 | 0.32 |
| **a0014.L** | 0.16 | 0.14 | 1.13 | 1747.10 | 0.26 |
| **a0014.Q** | 0.05 | 0.08 | 0.65 | 2837.96 | 0.51 |
| **a0014.C** | 0.15 | 0.09 | 1.72 | 2480.26 | 0.09 |
| **a0014^4** | 0.09 | 0.06 | 1.57 | 4128.02 | 0.12 |
| **dv_par_edu_birth** | 0.15 | 0.90 | 0.16 | 2678.69 | 0.87 |
| **dv_sc_age_5.L** | 0.06 | 0.12 | 0.49 | 1293.69 | 0.62 |
| **dv_sc_age_5.Q** | 0.01 | 0.05 | 0.28 | 4273.98 | 0.78 |
| **e216aNone** | 0.10 | 0.07 | 1.37 | 2346.04 | 0.17 |
| **dv_cog_abil_5** | -1.35 | 0.42 | -3.20 | 1163.31 | 0.00 |
| **dv_med_51** | -0.03 | 0.10 | -0.29 | 1454.31 | 0.77 |
| **d016a** | 0.10 | 0.07 | 1.49 | 2527.66 | 0.14 |
| **dv_bas_g** | -0.44 | 0.30 | -1.45 | 1664.04 | 0.15 |
| **dv_med_101** | 0.04 | 0.06 | 0.62 | 3089.14 | 0.53 |
| **bmi** | -0.50 | 0.27 | -1.82 | 3192.94 | 0.07 |
| **fclrg90.L** | 0.22 | 0.10 | 2.24 | 2917.50 | 0.03 |
| **fclrg90.Q** | 0.05 | 0.07 | 0.73 | 3664.23 | 0.47 |
| **fclrg90.C** | -0.04 | 0.10 | -0.43 | 1850.05 | 0.67 |
| **fclrg90^4** | -0.02 | 0.08 | -0.23 | 1941.42 | 0.82 |
| **tenure.L** | 0.05 | 0.07 | 0.75 | 1820.73 | 0.45 |
| **tenure.Q** | -0.05 | 0.09 | -0.56 | 1994.04 | 0.58 |
| **divorce1** | 0.03 | 0.07 | 0.37 | 3707.47 | 0.71 |
| **sepmumbcs1** | 0.31 | 0.14 | 2.21 | 2426.60 | 0.03 |
| **prmnh1** | 0.35 | 0.15 | 2.38 | 1732.19 | 0.02 |
| **crowdUp to 1** | -0.11 | 0.07 | -1.59 | 3341.30 | 0.11 |
| **ameniNo occasions** | 0.03 | 0.11 | 0.27 | 3905.79 | 0.79 |
| **brfed.L** | -0.01 | 0.06 | -0.17 | 2177.80 | 0.86 |
| **brfed.Q** | -0.01 | 0.10 | -0.05 | 1405.77 | 0.96 |
| **resmove.L** | 0.21 | 0.09 | 2.47 | 1741.05 | 0.01 |
| **resmove.Q** | 0.06 | 0.04 | 1.42 | 3585.45 | 0.16 |

*Table S64.* Raw output from modified Poisson regression (with log link) of behavioural and emotional problems at age 16 (rd6m_1) against median sleep duration (binarised to normal vs abnormal) derived from the sleep diary at age 46, adjusted for potential confounders (see main text Table 3) and mental health variables at age 42 (see Table S42). The parameter estimates in the “Estimate” column and the associated standard errors have not been exponentiated.

| Term | Estimate | Standard error | Statistic | Degrees of freedom | p-value |
|---|---|---|---|---|---|
| **(Intercept)** | -0.42 | 0.25 | -1.69 | 2359.86 | 0.09 |
| **rd6m_1** | 0.02 | 0.02 | 1.06 | 1406.77 | 0.29 |
| **BD9MAL** | 0.12 | 0.10 | 1.27 | 2196.11 | 0.20 |
| **BD9WEMWB** | 0.06 | 0.18 | 0.32 | 1660.34 | 0.75 |
| **a0005a** | -0.36 | 0.10 | -3.59 | 5050.87 | 0.00 |
| **a0043b.L** | -0.05 | 0.05 | -1.00 | 1785.97 | 0.32 |
| **a0043b.Q** | -0.02 | 0.06 | -0.40 | 2105.50 | 0.69 |
| **a0043b.C** | 0.00 | 0.06 | -0.01 | 1667.71 | 0.99 |
| **a0043b^4** | -0.06 | 0.04 | -1.31 | 3653.98 | 0.19 |
| **a0043b^5** | -0.01 | 0.07 | -0.19 | 2142.66 | 0.85 |
| **a0195a** | -0.39 | 0.47 | -0.85 | 1665.36 | 0.40 |
| **a0278** | -0.20 | 0.20 | -1.02 | 2431.90 | 0.31 |
| **a0014.L** | 0.01 | 0.07 | 0.20 | 1952.43 | 0.84 |
| **a0014.Q** | 0.03 | 0.05 | 0.56 | 1949.65 | 0.58 |
| **a0014.C** | 0.00 | 0.05 | 0.05 | 2588.02 | 0.96 |
| **a0014^4** | -0.01 | 0.04 | -0.26 | 1793.61 | 0.79 |
| **dv_par_edu_birth** | -0.38 | 0.46 | -0.83 | 2654.29 | 0.41 |
| **dv_sc_age_5.L** | -0.02 | 0.06 | -0.28 | 1541.98 | 0.78 |
| **dv_sc_age_5.Q** | 0.00 | 0.03 | -0.15 | 2928.62 | 0.88 |
| **e216aNone** | -0.01 | 0.05 | -0.10 | 1531.11 | 0.92 |
| **dv_cog_abil_5** | 0.05 | 0.28 | 0.16 | 857.08 | 0.87 |
| **dv_med_51** | 0.00 | 0.04 | 0.10 | 2943.71 | 0.92 |
| **d016a** | 0.05 | 0.03 | 1.70 | 4059.12 | 0.09 |
| **dv_bas_g** | -0.22 | 0.15 | -1.42 | 1927.60 | 0.15 |
| **dv_med_101** | 0.01 | 0.03 | 0.26 | 2805.75 | 0.79 |
| **bmi** | 0.19 | 0.17 | 1.07 | 2130.65 | 0.28 |
| **fclrg90.L** | 0.02 | 0.06 | 0.42 | 2510.38 | 0.68 |
| **fclrg90.Q** | 0.05 | 0.04 | 1.06 | 2441.75 | 0.29 |
| **fclrg90.C** | 0.06 | 0.04 | 1.52 | 3946.51 | 0.13 |
| **fclrg90^4** | 0.01 | 0.05 | 0.30 | 1818.04 | 0.77 |
| **tenure.L** | 0.01 | 0.03 | 0.45 | 2381.02 | 0.66 |
| **tenure.Q** | -0.03 | 0.05 | -0.71 | 2289.78 | 0.48 |
| **divorce1** | 0.03 | 0.04 | 0.70 | 3661.91 | 0.48 |
| **sepmumbcs1** | -0.07 | 0.09 | -0.78 | 2948.11 | 0.44 |
| **prmnh1** | 0.17 | 0.10 | 1.84 | 1755.93 | 0.07 |
| **crowdUp to 1** | -0.05 | 0.05 | -0.90 | 1878.78 | 0.37 |
| **ameniNo occasions** | -0.08 | 0.07 | -1.02 | 2190.83 | 0.31 |
| **brfed.L** | 0.03 | 0.03 | 0.80 | 2109.04 | 0.43 |
| **brfed.Q** | -0.01 | 0.03 | -0.43 | 3455.03 | 0.67 |
| **resmove.L** | -0.03 | 0.04 | -0.72 | 2804.79 | 0.47 |
| **resmove.Q** | 0.06 | 0.02 | 2.53 | 4306.53 | 0.01 |

*Table S65.* Raw output from modified Poisson regression (with log link) of behavioural and emotional problems at age 16 (rd6m_1) against median sleep duration (binarised to normal vs abnormal) derived from the activPAL algorithm at age 46, adjusted for potential confounders (see main text Table 3) and mental health variables at age 42 (see Table S42). The parameter estimates in the “Estimate” column and the associated standard errors have not been exponentiated.

| Term | Estimate | Standard error | Statistic | Degrees of freedom | p-value |
|---|---|---|---|---|---|
| **(Intercept)** | -0.85 | 0.36 | -2.37 | 1567.55 | 0.02 |
| **rd6m_1** | -0.04 | 0.03 | -1.60 | 1550.87 | 0.11 |
| **BD9MAL** | 0.18 | 0.11 | 1.58 | 1875.44 | 0.11 |
| **BD9WEMWB** | -0.15 | 0.19 | -0.83 | 1770.76 | 0.41 |
| **a0005a** | -0.01 | 0.17 | -0.07 | 1780.89 | 0.94 |
| **a0043b.L** | 0.00 | 0.06 | -0.02 | 1600.15 | 0.98 |
| **a0043b.Q** | -0.07 | 0.07 | -0.98 | 1778.57 | 0.33 |
| **a0043b.C** | -0.18 | 0.07 | -2.80 | 1705.56 | 0.01 |
| **a0043b^4** | 0.10 | 0.06 | 1.64 | 2049.06 | 0.10 |
| **a0043b^5** | 0.04 | 0.07 | 0.47 | 2188.75 | 0.64 |
| **a0195a** | -0.27 | 0.47 | -0.57 | 1991.11 | 0.57 |
| **a0278** | 0.36 | 0.18 | 2.00 | 3852.42 | 0.05 |
| **a0014.L** | 0.03 | 0.09 | 0.32 | 1540.40 | 0.75 |
| **a0014.Q** | 0.04 | 0.07 | 0.64 | 1545.16 | 0.53 |
| **a0014.C** | -0.01 | 0.05 | -0.15 | 2720.82 | 0.88 |
| **a0014^4** | -0.02 | 0.04 | -0.46 | 2385.05 | 0.64 |
| **dv_par_edu_birth** | -0.10 | 0.40 | -0.24 | 3901.26 | 0.81 |
| **dv_sc_age_5.L** | 0.15 | 0.08 | 1.80 | 1135.96 | 0.07 |
| **dv_sc_age_5.Q** | -0.03 | 0.04 | -0.95 | 2229.40 | 0.34 |
| **e216aNone** | 0.07 | 0.05 | 1.40 | 1937.27 | 0.16 |
| **dv_cog_abil_5** | 0.00 | 0.27 | 0.00 | 1037.57 | 1.00 |
| **dv_med_51** | 0.04 | 0.05 | 0.85 | 1905.82 | 0.40 |
| **d016a** | 0.09 | 0.04 | 2.08 | 2383.89 | 0.04 |
| **dv_bas_g** | -0.39 | 0.22 | -1.77 | 1266.03 | 0.08 |
| **dv_med_101** | 0.04 | 0.05 | 0.80 | 1541.13 | 0.42 |
| **bmi** | -0.26 | 0.24 | -1.06 | 1545.35 | 0.29 |
| **fclrg90.L** | -0.02 | 0.06 | -0.25 | 2566.70 | 0.80 |
| **fclrg90.Q** | -0.03 | 0.06 | -0.49 | 1677.89 | 0.62 |
| **fclrg90.C** | -0.10 | 0.06 | -1.81 | 2193.83 | 0.07 |
| **fclrg90^4** | 0.01 | 0.05 | 0.18 | 2081.00 | 0.85 |
| **tenure.L** | 0.03 | 0.05 | 0.65 | 1361.15 | 0.52 |
| **tenure.Q** | 0.09 | 0.05 | 1.66 | 2318.30 | 0.10 |
| **divorce1** | 0.14 | 0.04 | 3.16 | 3040.60 | 0.00 |
| **sepmumbcs1** | -0.10 | 0.14 | -0.72 | 1570.49 | 0.47 |
| **prmnh1** | -0.23 | 0.18 | -1.26 | 1147.68 | 0.21 |
| **crowdUp to 1** | 0.03 | 0.06 | 0.46 | 1807.19 | 0.65 |
| **ameniNo occasions** | -0.09 | 0.08 | -1.19 | 2458.34 | 0.24 |
| **brfed.L** | 0.01 | 0.04 | 0.26 | 1629.53 | 0.79 |
| **brfed.Q** | 0.04 | 0.05 | 0.94 | 2222.14 | 0.35 |
| **resmove.L** | 0.00 | 0.04 | 0.06 | 3421.75 | 0.95 |
| **resmove.Q** | -0.06 | 0.03 | -2.15 | 3527.61 | 0.03 |

*Table S66.* Raw output from modified Poisson regression (with log link) of behavioural and emotional problems at age 16 (rd6m_1) against median sleep duration (binarised to normal vs abnormal) derived from the van der Berg et al. algorithm at age 46, adjusted for potential confounders (see main text Table 3) and mental health variables at age 42 (see Table S42). The parameter estimates in the "Estimate" column and the associated standard errors have not been exponentiated.

| Term | Estimate | Standard error | Statistic | Degrees of freedom | p-value |
|---|---|---|---|---|---|
| **(Intercept)** | -0.90 | 0.37 | -2.42 | 1348.52 | 0.02 |
| **rd6m_1** | 0.01 | 0.02 | 0.50 | 1700.09 | 0.61 |
| **BD9MAL** | 0.27 | 0.14 | 2.01 | 1339.17 | 0.04 |
| **BD9WEMWB** | -0.12 | 0.24 | -0.49 | 1184.67 | 0.62 |
| **a0005a** | 0.14 | 0.20 | 0.69 | 1237.11 | 0.49 |
| **a0043b.L** | -0.05 | 0.04 | -1.24 | 2918.71 | 0.21 |
| **a0043b.Q** | 0.02 | 0.06 | 0.30 | 2148.42 | 0.76 |
| **a0043b.C** | 0.06 | 0.04 | 1.52 | 4640.26 | 0.13 |
| **a0043b^4** | 0.03 | 0.05 | 0.50 | 2591.87 | 0.62 |
| **a0043b^5** | 0.04 | 0.07 | 0.58 | 2505.34 | 0.56 |
| **a0195a** | -0.09 | 0.33 | -0.26 | 3925.90 | 0.79 |
| **a0278** | -0.09 | 0.27 | -0.35 | 1608.61 | 0.73 |
| **a0014.L** | 0.08 | 0.07 | 1.02 | 2131.69 | 0.31 |
| **a0014.Q** | 0.01 | 0.06 | 0.16 | 1547.23 | 0.87 |
| **a0014.C** | -0.04 | 0.09 | -0.41 | 1080.31 | 0.68 |
| **a0014^4** | 0.03 | 0.05 | 0.64 | 1527.21 | 0.52 |
| **dv_par_edu_birth** | 0.46 | 0.43 | 1.07 | 1818.91 | 0.29 |
| **dv_sc_age_5.L** | 0.02 | 0.09 | 0.19 | 989.27 | 0.85 |
| **dv_sc_age_5.Q** | -0.01 | 0.05 | -0.17 | 1428.72 | 0.87 |
| **e216aNone** | -0.12 | 0.05 | -2.32 | 1416.66 | 0.02 |
| **dv_cog_abil_5** | -0.25 | 0.27 | -0.91 | 993.57 | 0.36 |
| **dv_med_51** | 0.03 | 0.04 | 0.64 | 2621.18 | 0.52 |
| **d016a** | 0.19 | 0.05 | 3.53 | 1424.59 | 0.00 |
| **dv_bas_g** | -0.23 | 0.18 | -1.26 | 1541.58 | 0.21 |
| **dv_med_101** | 0.03 | 0.05 | 0.62 | 1539.24 | 0.54 |
| **bmi** | 0.52 | 0.21 | 2.52 | 1652.31 | 0.01 |
| **fclrg90.L** | -0.13 | 0.08 | -1.53 | 1558.53 | 0.13 |
| **fclrg90.Q** | -0.05 | 0.06 | -0.93 | 1805.47 | 0.35 |
| **fclrg90.C** | 0.03 | 0.07 | 0.36 | 1492.43 | 0.72 |
| **fclrg90^4** | 0.01 | 0.05 | 0.12 | 1735.27 | 0.90 |
| **tenure.L** | 0.03 | 0.04 | 0.80 | 1454.77 | 0.42 |
| **tenure.Q** | -0.05 | 0.06 | -0.79 | 1619.45 | 0.43 |
| **divorce1** | -0.02 | 0.07 | -0.25 | 1525.18 | 0.80 |
| **sepmumbcs1** | 0.20 | 0.10 | 2.05 | 1968.83 | 0.04 |
| **prmnh1** | -0.06 | 0.11 | -0.50 | 1921.33 | 0.61 |
| **crowdUp to 1** | 0.07 | 0.05 | 1.37 | 2098.87 | 0.17 |
| **ameniNo occasions** | -0.16 | 0.10 | -1.65 | 1483.46 | 0.10 |
| **brfed.L** | 0.03 | 0.04 | 0.89 | 2252.31 | 0.38 |
| **brfed.Q** | -0.05 | 0.05 | -0.91 | 1597.59 | 0.36 |
| **resmove.L** | 0.02 | 0.06 | 0.38 | 1497.18 | 0.70 |
| **resmove.Q** | 0.03 | 0.02 | 1.11 | 4910.43 | 0.27 |

*Table S67.* Raw output from modified Poisson regression (with log link) of behavioural and emotional problems at age 16 (rd6m_1) against median sleep duration (binarised to normal vs abnormal) derived from the Winkler et al. algorithm at age 46, adjusted for potential confounders (see main text Table 3) and mental health variables at age 42 (see Table S42). The parameter estimates in the “Estimate” column and the associated standard errors have not been exponentiated.

| Term | Estimate | Standard error | Statistic | Degrees of freedom | p-value |
|---|---|---|---|---|---|
| **(Intercept)** | -0.37 | 0.45 | -0.82 | 2337.89 | 0.41 |
| **intbcsz** | -0.01 | 0.03 | -0.24 | 1957.42 | 0.81 |
| **BD9MAL** | 1.01 | 0.14 | 7.12 | 2418.61 | 0.00 |
| **BD9WEMWB** | -1.12 | 0.23 | -4.95 | 2711.12 | 0.00 |
| **a0005a** | -0.22 | 0.15 | -1.45 | 8040.11 | 0.15 |
| **a0043b.L** | 0.13 | 0.06 | 2.15 | 3426.21 | 0.03 |
| **a0043b.Q** | 0.07 | 0.09 | 0.79 | 2735.09 | 0.43 |
| **a0043b.C** | 0.02 | 0.08 | 0.22 | 2847.39 | 0.82 |
| **a0043b^4** | -0.02 | 0.11 | -0.23 | 2006.30 | 0.82 |
| **a0043b^5** | -0.06 | 0.11 | -0.54 | 2875.54 | 0.59 |
| **a0195a** | -0.20 | 0.49 | -0.40 | 4301.85 | 0.69 |
| **a0278** | -0.55 | 0.25 | -2.24 | 7269.57 | 0.03 |
| **a0014.L** | 0.13 | 0.10 | 1.23 | 3209.18 | 0.22 |
| **a0014.Q** | -0.01 | 0.06 | -0.17 | 7251.17 | 0.87 |
| **a0014.C** | 0.12 | 0.08 | 1.56 | 3277.93 | 0.12 |
| **a0014^4** | 0.05 | 0.07 | 0.73 | 2605.83 | 0.47 |
| **dv_par_edu_birth** | -1.06 | 1.20 | -0.88 | 2914.75 | 0.38 |
| **dv_sc_age_5.L** | 0.15 | 0.10 | 1.47 | 1638.73 | 0.14 |
| **dv_sc_age_5.Q** | -0.05 | 0.04 | -1.11 | 4277.36 | 0.27 |
| **e216aNone** | 0.01 | 0.06 | 0.11 | 2798.29 | 0.91 |
| **dv_cog_abil_5** | -0.52 | 0.31 | -1.68 | 1643.35 | 0.09 |
| **dv_med_51** | 0.03 | 0.06 | 0.48 | 4014.40 | 0.63 |
| **d016a** | 0.21 | 0.05 | 3.98 | 4386.41 | 0.00 |
| **dv_bas_g** | -0.38 | 0.23 | -1.68 | 2431.35 | 0.09 |
| **dv_med_101** | 0.00 | 0.05 | -0.09 | 6396.35 | 0.93 |
| **bmi** | -0.04 | 0.25 | -0.16 | 3557.15 | 0.87 |
| **fclrg90.L** | -0.01 | 0.09 | -0.06 | 3504.30 | 0.95 |
| **fclrg90.Q** | 0.02 | 0.06 | 0.36 | 5579.24 | 0.72 |
| **fclrg90.C** | -0.08 | 0.08 | -1.08 | 3410.89 | 0.28 |
| **fclrg90^4** | 0.02 | 0.05 | 0.45 | 5831.61 | 0.65 |
| **tenure.L** | 0.10 | 0.05 | 2.10 | 2956.38 | 0.04 |
| **tenure.Q** | -0.06 | 0.06 | -1.01 | 3695.82 | 0.31 |
| **divorce1** | 0.07 | 0.06 | 1.20 | 4324.10 | 0.23 |
| **sepmumbcs1** | 0.09 | 0.11 | 0.79 | 4856.68 | 0.43 |
| **prmnh1** | 0.18 | 0.13 | 1.33 | 2012.75 | 0.18 |
| **crowdUp to 1** | -0.16 | 0.06 | -2.87 | 4728.31 | 0.00 |
| **ameniNo occasions** | -0.05 | 0.12 | -0.43 | 2439.35 | 0.67 |
| **brfed.L** | 0.08 | 0.05 | 1.48 | 3948.51 | 0.14 |
| **brfed.Q** | -0.05 | 0.06 | -0.81 | 3797.46 | 0.42 |
| **resmove.L** | 0.14 | 0.06 | 2.46 | 4098.96 | 0.01 |
| **resmove.Q** | -0.02 | 0.03 | -0.65 | 8923.34 | 0.52 |

*Table S68.* Raw output from modified Poisson regression (with log link) of internalising behaviour at age 16 (intbcsz) against self-reported average sleep duration (binarised to normal vs abnormal) at age 46, adjusted for potential confounders (see main text Table 3) and mental health variables at age 42 (see Table S42). The parameter estimates in the "Estimate" column and the associated standard errors have not been exponentiated.

| Term | Estimate | Standard error | Statistic | Degrees of freedom | p-value |
|---|---|---|---|---|---|
| **(Intercept)** | -0.70 | 0.50 | -1.39 | 1948.31 | 0.17 |
| **intbcsz** | 0.02 | 0.02 | 0.99 | 4936.50 | 0.32 |
| **BD9MAL** | 0.36 | 0.13 | 2.77 | 4140.31 | 0.01 |
| **BD9WEMWB** | -0.98 | 0.23 | -4.32 | 3175.50 | 0.00 |
| **a0005a** | 0.48 | 0.22 | 2.22 | 2604.17 | 0.03 |
| **a0043b.L** | 0.15 | 0.07 | 2.05 | 2601.17 | 0.04 |
| **a0043b.Q** | 0.17 | 0.10 | 1.70 | 2521.90 | 0.09 |
| **a0043b.C** | 0.04 | 0.10 | 0.40 | 2174.06 | 0.69 |
| **a0043b^4** | -0.10 | 0.09 | -1.11 | 2933.40 | 0.27 |
| **a0043b^5** | -0.07 | 0.12 | -0.58 | 3252.35 | 0.56 |
| **a0195a** | -0.10 | 0.65 | -0.15 | 2516.73 | 0.88 |
| **a0278** | 0.28 | 0.28 | 0.97 | 4708.59 | 0.33 |
| **a0014.L** | 0.16 | 0.14 | 1.14 | 1736.17 | 0.26 |
| **a0014.Q** | 0.05 | 0.08 | 0.67 | 2841.02 | 0.50 |
| **a0014.C** | 0.16 | 0.09 | 1.73 | 2481.66 | 0.08 |
| **a0014^4** | 0.09 | 0.06 | 1.57 | 4054.49 | 0.12 |
| **dv_par_edu_birth** | 0.16 | 0.89 | 0.18 | 2738.98 | 0.86 |
| **dv_sc_age_5.L** | 0.06 | 0.13 | 0.50 | 1257.09 | 0.61 |
| **dv_sc_age_5.Q** | 0.01 | 0.05 | 0.28 | 4234.87 | 0.78 |
| **e216aNone** | 0.10 | 0.07 | 1.37 | 2319.74 | 0.17 |
| **dv_cog_abil_5** | -1.36 | 0.42 | -3.20 | 1161.03 | 0.00 |
| **dv_med_51** | -0.03 | 0.10 | -0.28 | 1489.03 | 0.78 |
| **d016a** | 0.11 | 0.07 | 1.65 | 2637.20 | 0.10 |
| **dv_bas_g** | -0.45 | 0.31 | -1.46 | 1638.92 | 0.15 |
| **dv_med_101** | 0.03 | 0.06 | 0.59 | 2984.83 | 0.56 |
| **bmi** | -0.49 | 0.28 | -1.78 | 3151.96 | 0.08 |
| **fclrg90.L** | 0.21 | 0.10 | 2.16 | 2885.83 | 0.03 |
| **fclrg90.Q** | 0.04 | 0.07 | 0.66 | 3548.60 | 0.51 |
| **fclrg90.C** | -0.05 | 0.10 | -0.44 | 1853.66 | 0.66 |
| **fclrg90^4** | -0.02 | 0.08 | -0.25 | 1952.14 | 0.80 |
| **tenure.L** | 0.05 | 0.07 | 0.75 | 1810.80 | 0.45 |
| **tenure.Q** | -0.05 | 0.09 | -0.54 | 1965.54 | 0.59 |
| **divorce1** | 0.03 | 0.07 | 0.37 | 3668.84 | 0.71 |
| **sepmumbcs1** | 0.33 | 0.14 | 2.43 | 2598.16 | 0.02 |
| **prmnh1** | 0.33 | 0.15 | 2.24 | 1738.99 | 0.02 |
| **crowdUp to 1** | -0.11 | 0.07 | -1.57 | 3333.16 | 0.12 |
| **ameniNo occasions** | 0.02 | 0.11 | 0.15 | 3971.23 | 0.88 |
| **brfed.L** | -0.01 | 0.06 | -0.14 | 2216.47 | 0.89 |
| **brfed.Q** | 0.00 | 0.10 | -0.03 | 1376.53 | 0.98 |
| **resmove.L** | 0.21 | 0.09 | 2.43 | 1733.03 | 0.02 |
| **resmove.Q** | 0.06 | 0.04 | 1.41 | 3603.00 | 0.16 |

*Table S69.* Raw output from modified Poisson regression (with log link) of internalising behaviour at age 16 (intbcsz) against median sleep duration (binarised to normal vs abnormal) derived from the sleep diary at age 46, adjusted for potential confounders (see main text Table 3) and mental health variables at age 42 (see Table S42). The parameter estimates in the “Estimate” column and the associated standard errors have not been exponentiated.

| Term | Estimate | Standard error | Statistic | Degrees of freedom | p-value |
|---|---|---|---|---|---|
| **(Intercept)** | -0.43 | 0.25 | -1.70 | 2363.81 | 0.09 |
| **intbcsz** | 0.02 | 0.02 | 0.90 | 1835.50 | 0.37 |
| **BD9MAL** | 0.12 | 0.10 | 1.11 | 1935.15 | 0.27 |
| **BD9WEMWB** | 0.06 | 0.18 | 0.34 | 1673.71 | 0.73 |
| **a0005a** | -0.35 | 0.10 | -3.50 | 4817.80 | 0.00 |
| **a0043b.L** | -0.05 | 0.05 | -1.04 | 1796.41 | 0.30 |
| **a0043b.Q** | -0.02 | 0.06 | -0.40 | 2117.40 | 0.69 |
| **a0043b.C** | 0.00 | 0.06 | -0.01 | 1670.98 | 1.00 |
| **a0043b^4** | -0.06 | 0.04 | -1.32 | 3620.35 | 0.19 |
| **a0043b^5** | -0.01 | 0.07 | -0.19 | 2153.54 | 0.85 |
| **a0195a** | -0.39 | 0.46 | -0.85 | 1676.36 | 0.39 |
| **a0278** | -0.20 | 0.20 | -0.99 | 2375.91 | 0.32 |
| **a0014.L** | 0.01 | 0.07 | 0.20 | 1959.49 | 0.84 |
| **a0014.Q** | 0.03 | 0.05 | 0.57 | 1966.91 | 0.57 |
| **a0014.C** | 0.00 | 0.05 | 0.06 | 2553.50 | 0.95 |
| **a0014^4** | -0.01 | 0.04 | -0.26 | 1798.84 | 0.80 |
| **dv_par_edu_birth** | -0.38 | 0.46 | -0.83 | 2688.37 | 0.41 |
| **dv_sc_age_5.L** | -0.02 | 0.06 | -0.24 | 1546.63 | 0.81 |
| **dv_sc_age_5.Q** | 0.00 | 0.03 | -0.16 | 2955.08 | 0.87 |
| **e216aNone** | -0.01 | 0.05 | -0.11 | 1545.88 | 0.91 |
| **dv_cog_abil_5** | 0.04 | 0.28 | 0.15 | 853.57 | 0.88 |
| **dv_med_51** | 0.00 | 0.04 | 0.08 | 3058.87 | 0.94 |
| **d016a** | 0.06 | 0.03 | 1.68 | 3545.98 | 0.09 |
| **dv_bas_g** | -0.21 | 0.15 | -1.38 | 1863.45 | 0.17 |
| **dv_med_101** | 0.01 | 0.03 | 0.20 | 2708.93 | 0.84 |
| **bmi** | 0.19 | 0.17 | 1.09 | 2139.77 | 0.28 |
| **fclrg90.L** | 0.02 | 0.06 | 0.36 | 2516.22 | 0.72 |
| **fclrg90.Q** | 0.05 | 0.04 | 1.04 | 2439.64 | 0.30 |
| **fclrg90.C** | 0.06 | 0.04 | 1.50 | 4033.73 | 0.13 |
| **fclrg90^4** | 0.01 | 0.05 | 0.27 | 1787.42 | 0.78 |
| **tenure.L** | 0.01 | 0.03 | 0.45 | 2421.05 | 0.65 |
| **tenure.Q** | -0.03 | 0.05 | -0.70 | 2277.66 | 0.48 |
| **divorce1** | 0.03 | 0.04 | 0.67 | 3744.75 | 0.50 |
| **sepmumbcs1** | -0.06 | 0.09 | -0.71 | 3091.68 | 0.48 |
| **prmnh1** | 0.17 | 0.09 | 1.80 | 1871.89 | 0.07 |
| **crowdUp to 1** | -0.05 | 0.05 | -0.88 | 1880.07 | 0.38 |
| **ameniNo occasions** | -0.08 | 0.07 | -1.04 | 2159.58 | 0.30 |
| **brfed.L** | 0.03 | 0.03 | 0.82 | 2097.62 | 0.41 |
| **brfed.Q** | -0.01 | 0.03 | -0.39 | 3529.37 | 0.69 |
| **resmove.L** | -0.03 | 0.04 | -0.73 | 2816.99 | 0.47 |
| **resmove.Q** | 0.06 | 0.02 | 2.53 | 4357.79 | 0.01 |

*Table S70.* Raw output from modified Poisson regression (with log link) of internalising behaviour at age 16 (intbcsz) against median sleep duration (binarised to normal vs abnormal) derived from the activPAL algorithm at age 46, adjusted for potential confounders (see main text Table 3) and mental health variables at age 42 (see Table S42). The parameter estimates in the "Estimate" column and the associated standard errors have not been exponentiated.

| Term | Estimate | Standard error | Statistic | Degrees of freedom | p-value |
|---|---|---|---|---|---|
| **(Intercept)** | -0.86 | 0.37 | -2.35 | 1514.66 | 0.02 |
| **intbcsz** | 0.00 | 0.03 | -0.03 | 1176.12 | 0.98 |
| **BD9MAL** | 0.17 | 0.12 | 1.47 | 1783.56 | 0.14 |
| **BD9WEMWB** | -0.15 | 0.19 | -0.79 | 1778.44 | 0.43 |
| **a0005a** | -0.01 | 0.17 | -0.08 | 1780.13 | 0.94 |
| **a0043b.L** | 0.00 | 0.06 | 0.02 | 1604.65 | 0.98 |
| **a0043b.Q** | -0.07 | 0.07 | -0.99 | 1816.13 | 0.32 |
| **a0043b.C** | -0.18 | 0.07 | -2.80 | 1703.67 | 0.01 |
| **a0043b^4** | 0.10 | 0.06 | 1.62 | 2023.56 | 0.10 |
| **a0043b^5** | 0.03 | 0.07 | 0.46 | 2172.11 | 0.65 |
| **a0195a** | -0.27 | 0.47 | -0.59 | 2013.38 | 0.56 |
| **a0278** | 0.37 | 0.18 | 2.00 | 3765.82 | 0.05 |
| **a0014.L** | 0.03 | 0.09 | 0.31 | 1529.74 | 0.76 |
| **a0014.Q** | 0.04 | 0.07 | 0.63 | 1560.30 | 0.53 |
| **a0014.C** | -0.01 | 0.05 | -0.17 | 2637.68 | 0.86 |
| **a0014^4** | -0.02 | 0.04 | -0.47 | 2379.92 | 0.64 |
| **dv_par_edu_birth** | -0.10 | 0.41 | -0.25 | 3817.69 | 0.80 |
| **dv_sc_age_5.L** | 0.15 | 0.08 | 1.81 | 1160.99 | 0.07 |
| **dv_sc_age_5.Q** | -0.03 | 0.04 | -0.93 | 2198.27 | 0.35 |
| **e216aNone** | 0.07 | 0.05 | 1.39 | 1854.45 | 0.17 |
| **dv_cog_abil_5** | 0.00 | 0.27 | 0.02 | 1040.80 | 0.99 |
| **dv_med_51** | 0.04 | 0.05 | 0.80 | 1843.93 | 0.43 |
| **d016a** | 0.08 | 0.04 | 1.96 | 2372.73 | 0.05 |
| **dv_bas_g** | -0.38 | 0.22 | -1.74 | 1270.77 | 0.08 |
| **dv_med_101** | 0.04 | 0.05 | 0.80 | 1603.17 | 0.42 |
| **bmi** | -0.26 | 0.24 | -1.06 | 1541.52 | 0.29 |
| **fclrg90.L** | -0.01 | 0.07 | -0.20 | 2506.42 | 0.84 |
| **fclrg90.Q** | -0.03 | 0.06 | -0.46 | 1667.41 | 0.64 |
| **fclrg90.C** | -0.10 | 0.06 | -1.77 | 2142.17 | 0.08 |
| **fclrg90^4** | 0.01 | 0.05 | 0.21 | 2101.77 | 0.83 |
| **tenure.L** | 0.03 | 0.05 | 0.66 | 1380.73 | 0.51 |
| **tenure.Q** | 0.09 | 0.06 | 1.62 | 2262.50 | 0.11 |
| **divorce1** | 0.14 | 0.05 | 2.95 | 2667.68 | 0.00 |
| **sepmumbcs1** | -0.11 | 0.14 | -0.80 | 1563.93 | 0.42 |
| **prmnh1** | -0.23 | 0.19 | -1.22 | 1126.47 | 0.22 |
| **crowdUp to 1** | 0.03 | 0.06 | 0.47 | 1827.56 | 0.64 |
| **ameniNo occasions** | -0.08 | 0.08 | -1.09 | 2418.77 | 0.28 |
| **brfed.L** | 0.01 | 0.04 | 0.24 | 1626.81 | 0.81 |
| **brfed.Q** | 0.04 | 0.05 | 0.93 | 2270.66 | 0.35 |
| **resmove.L** | 0.00 | 0.04 | 0.09 | 3500.09 | 0.93 |
| **resmove.Q** | -0.06 | 0.03 | -2.14 | 3568.23 | 0.03 |

*Table S71.* Raw output from modified Poisson regression (with log link) of internalising behaviour at age 16 (intbcsz) against median sleep duration (binarised to normal vs abnormal) derived from the van der Berg et al. algorithm at age 46, adjusted for potential confounders (see main text Table 3) and mental health variables at age 42 (see Table S42). The parameter estimates in the “Estimate” column and the associated standard errors have not been exponentiated.

| Term | Estimate | Standard error | Statistic | Degrees of freedom | p-value |
|---|---|---|---|---|---|
| **(Intercept)** | -0.90 | 0.37 | -2.46 | 1377.37 | 0.01 |
| **intbcsz** | 0.01 | 0.03 | 0.36 | 1257.73 | 0.72 |
| **BD9MAL** | 0.27 | 0.14 | 1.90 | 1280.51 | 0.06 |
| **BD9WEMWB** | -0.11 | 0.24 | -0.48 | 1189.49 | 0.63 |
| **a0005a** | 0.14 | 0.20 | 0.71 | 1238.66 | 0.48 |
| **a0043b.L** | -0.05 | 0.04 | -1.28 | 2917.77 | 0.20 |
| **a0043b.Q** | 0.02 | 0.06 | 0.29 | 2136.13 | 0.77 |
| **a0043b.C** | 0.06 | 0.04 | 1.52 | 4592.30 | 0.13 |
| **a0043b^4** | 0.03 | 0.05 | 0.50 | 2642.40 | 0.62 |
| **a0043b^5** | 0.04 | 0.07 | 0.58 | 2510.47 | 0.56 |
| **a0195a** | -0.09 | 0.33 | -0.27 | 3917.86 | 0.79 |
| **a0278** | -0.09 | 0.27 | -0.34 | 1591.75 | 0.74 |
| **a0014.L** | 0.08 | 0.07 | 1.01 | 2117.84 | 0.31 |
| **a0014.Q** | 0.01 | 0.06 | 0.16 | 1534.16 | 0.87 |
| **a0014.C** | -0.04 | 0.09 | -0.40 | 1081.00 | 0.69 |
| **a0014^4** | 0.03 | 0.05 | 0.65 | 1528.62 | 0.52 |
| **dv_par_edu_birth** | 0.46 | 0.42 | 1.08 | 1837.60 | 0.28 |
| **dv_sc_age_5.L** | 0.02 | 0.09 | 0.20 | 990.00 | 0.84 |
| **dv_sc_age_5.Q** | -0.01 | 0.05 | -0.18 | 1428.32 | 0.86 |
| **e216aNone** | -0.12 | 0.05 | -2.33 | 1422.69 | 0.02 |
| **dv_cog_abil_5** | -0.25 | 0.27 | -0.91 | 993.34 | 0.36 |
| **dv_med_51** | 0.03 | 0.04 | 0.61 | 2488.11 | 0.54 |
| **d016a** | 0.19 | 0.05 | 3.63 | 1478.47 | 0.00 |
| **dv_bas_g** | -0.23 | 0.18 | -1.28 | 1598.43 | 0.20 |
| **dv_med_101** | 0.03 | 0.05 | 0.61 | 1607.62 | 0.54 |
| **bmi** | 0.52 | 0.21 | 2.52 | 1644.12 | 0.01 |
| **fclrg90.L** | -0.13 | 0.08 | -1.56 | 1565.13 | 0.12 |
| **fclrg90.Q** | -0.05 | 0.06 | -0.94 | 1801.70 | 0.35 |
| **fclrg90.C** | 0.02 | 0.07 | 0.35 | 1480.86 | 0.72 |
| **fclrg90^4** | 0.01 | 0.05 | 0.12 | 1746.63 | 0.91 |
| **tenure.L** | 0.03 | 0.04 | 0.81 | 1457.55 | 0.42 |
| **tenure.Q** | -0.05 | 0.06 | -0.78 | 1634.15 | 0.43 |
| **divorce1** | -0.02 | 0.07 | -0.26 | 1435.53 | 0.79 |
| **sepmumbcs1** | 0.20 | 0.10 | 2.07 | 1943.89 | 0.04 |
| **prmnh1** | -0.06 | 0.11 | -0.54 | 1909.72 | 0.59 |
| **crowdUp to 1** | 0.07 | 0.05 | 1.37 | 2074.60 | 0.17 |
| **ameniNo occasions** | -0.16 | 0.09 | -1.70 | 1526.18 | 0.09 |
| **brfed.L** | 0.03 | 0.04 | 0.90 | 2241.90 | 0.37 |
| **brfed.Q** | -0.05 | 0.05 | -0.90 | 1593.85 | 0.37 |
| **resmove.L** | 0.02 | 0.06 | 0.38 | 1509.22 | 0.70 |
| **resmove.Q** | 0.02 | 0.02 | 1.08 | 4911.55 | 0.28 |

*Table S72.* Raw output from modified Poisson regression (with log link) of internalising behaviour at age 16 (intbcsz) against median sleep duration (binarised to normal vs abnormal) derived from the Winkler et al. algorithm at age 46, adjusted for potential confounders (see main text Table 3) and mental health variables at age 42 (see Table S42). The parameter estimates in the “Estimate” column and the associated standard errors have not been exponentiated.

| Term | Estimate | Standard error | Statistic | Degrees of freedom | p-value |
|---|---|---|---|---|---|
| **(Intercept)** | -0.45 | 0.44 | -1.04 | 2471.45 | 0.30 |
| **extbcsz** | 0.05 | 0.03 | 1.81 | 1609.72 | 0.07 |
| **BD9MAL** | 0.98 | 0.14 | 6.97 | 2424.68 | 0.00 |
| **BD9WEMWB** | -1.08 | 0.23 | -4.65 | 2571.05 | 0.00 |
| **a0005a** | -0.20 | 0.15 | -1.34 | 8868.80 | 0.18 |
| **a0043b.L** | 0.13 | 0.06 | 2.13 | 3436.96 | 0.03 |
| **a0043b.Q** | 0.07 | 0.09 | 0.78 | 2717.32 | 0.44 |
| **a0043b.C** | 0.02 | 0.08 | 0.24 | 2764.74 | 0.81 |
| **a0043b^4** | -0.02 | 0.11 | -0.21 | 1984.65 | 0.83 |
| **a0043b^5** | -0.07 | 0.11 | -0.59 | 2818.88 | 0.56 |
| **a0195a** | -0.17 | 0.50 | -0.34 | 4259.61 | 0.73 |
| **a0278** | -0.55 | 0.25 | -2.22 | 7221.36 | 0.03 |
| **a0014.L** | 0.12 | 0.10 | 1.19 | 3227.56 | 0.23 |
| **a0014.Q** | -0.01 | 0.06 | -0.21 | 7228.37 | 0.84 |
| **a0014.C** | 0.12 | 0.08 | 1.57 | 3323.78 | 0.12 |
| **a0014^4** | 0.05 | 0.07 | 0.72 | 2604.05 | 0.47 |
| **dv_par_edu_birth** | -1.05 | 1.19 | -0.88 | 2948.00 | 0.38 |
| **dv_sc_age_5.L** | 0.15 | 0.10 | 1.43 | 1642.34 | 0.15 |
| **dv_sc_age_5.Q** | -0.05 | 0.04 | -1.11 | 4187.71 | 0.27 |
| **e216aNone** | 0.01 | 0.06 | 0.19 | 2801.37 | 0.85 |
| **dv_cog_abil_5** | -0.50 | 0.31 | -1.61 | 1652.17 | 0.11 |
| **dv_med_51** | 0.02 | 0.06 | 0.34 | 4205.47 | 0.74 |
| **d016a** | 0.20 | 0.05 | 3.68 | 4199.55 | 0.00 |
| **dv_bas_g** | -0.34 | 0.23 | -1.47 | 2328.32 | 0.14 |
| **dv_med_101** | -0.01 | 0.05 | -0.16 | 6413.43 | 0.87 |
| **bmi** | -0.03 | 0.24 | -0.11 | 3683.04 | 0.92 |
| **fclrg90.L** | -0.01 | 0.09 | -0.08 | 3306.77 | 0.93 |
| **fclrg90.Q** | 0.02 | 0.06 | 0.33 | 5407.46 | 0.74 |
| **fclrg90.C** | -0.08 | 0.08 | -1.09 | 3317.96 | 0.28 |
| **fclrg90^4** | 0.02 | 0.05 | 0.44 | 5617.82 | 0.66 |
| **tenure.L** | 0.10 | 0.05 | 2.07 | 2936.49 | 0.04 |
| **tenure.Q** | -0.06 | 0.06 | -1.02 | 3883.07 | 0.31 |
| **divorce1** | 0.05 | 0.06 | 0.86 | 3911.04 | 0.39 |
| **sepmumbcs1** | 0.08 | 0.11 | 0.72 | 5012.34 | 0.47 |
| **prmnh1** | 0.15 | 0.14 | 1.06 | 1859.29 | 0.29 |
| **crowdUp to 1** | -0.15 | 0.06 | -2.67 | 4605.73 | 0.01 |
| **ameniNo occasions** | -0.05 | 0.11 | -0.45 | 2632.29 | 0.65 |
| **brfed.L** | 0.08 | 0.05 | 1.49 | 3973.91 | 0.14 |
| **brfed.Q** | -0.05 | 0.06 | -0.79 | 3648.35 | 0.43 |
| **resmove.L** | 0.14 | 0.06 | 2.43 | 4130.24 | 0.02 |
| **resmove.Q** | -0.02 | 0.03 | -0.70 | 8970.89 | 0.48 |

*Table S73.* Raw output from modified Poisson regression (with log link) of externalising behaviour at age 16 (extbcsz) against self-reported average sleep duration (binarised to normal vs abnormal) at age 46, adjusted for potential confounders (see main text Table 3) and mental health variables at age 42 (see Table S42). The parameter estimates in the "Estimate" column and the associated standard errors have not been exponentiated.

| Term | Estimate | Standard error | Statistic | Degrees of freedom | p-value |
|---|---|---|---|---|---|
| **(Intercept)** | -0.75 | 0.50 | -1.49 | 1971.05 | 0.14 |
| **extbcsz** | 0.04 | 0.04 | 1.04 | 1235.73 | 0.30 |
| **BD9MAL** | 0.35 | 0.12 | 2.82 | 4475.82 | 0.00 |
| **BD9WEMWB** | -0.97 | 0.22 | -4.33 | 3328.08 | 0.00 |
| **a0005a** | 0.49 | 0.21 | 2.34 | 2770.62 | 0.02 |
| **a0043b.L** | 0.15 | 0.07 | 2.04 | 2560.26 | 0.04 |
| **a0043b.Q** | 0.17 | 0.10 | 1.69 | 2503.91 | 0.09 |
| **a0043b.C** | 0.04 | 0.09 | 0.42 | 2218.30 | 0.67 |
| **a0043b^4** | -0.10 | 0.09 | -1.09 | 2954.98 | 0.27 |
| **a0043b^5** | -0.07 | 0.12 | -0.60 | 3211.24 | 0.55 |
| **a0195a** | -0.07 | 0.66 | -0.11 | 2467.85 | 0.91 |
| **a0278** | 0.27 | 0.29 | 0.94 | 4558.81 | 0.35 |
| **a0014.L** | 0.16 | 0.14 | 1.12 | 1738.52 | 0.26 |
| **a0014.Q** | 0.05 | 0.08 | 0.64 | 2885.53 | 0.52 |
| **a0014.C** | 0.16 | 0.09 | 1.74 | 2473.56 | 0.08 |
| **a0014^4** | 0.09 | 0.06 | 1.56 | 4098.05 | 0.12 |
| **dv_par_edu_birth** | 0.17 | 0.88 | 0.19 | 2749.57 | 0.85 |
| **dv_sc_age_5.L** | 0.06 | 0.13 | 0.48 | 1245.81 | 0.63 |
| **dv_sc_age_5.Q** | 0.01 | 0.05 | 0.28 | 4259.27 | 0.78 |
| **e216aNone** | 0.10 | 0.07 | 1.41 | 2307.10 | 0.16 |
| **dv_cog_abil_5** | -1.34 | 0.42 | -3.19 | 1172.46 | 0.00 |
| **dv_med_51** | -0.03 | 0.10 | -0.32 | 1522.41 | 0.75 |
| **d016a** | 0.10 | 0.07 | 1.55 | 2643.91 | 0.12 |
| **dv_bas_g** | -0.43 | 0.31 | -1.40 | 1639.43 | 0.16 |
| **dv_med_101** | 0.04 | 0.06 | 0.61 | 3049.34 | 0.54 |
| **bmi** | -0.48 | 0.27 | -1.77 | 3217.40 | 0.08 |
| **fclrg90.L** | 0.21 | 0.10 | 2.17 | 2898.58 | 0.03 |
| **fclrg90.Q** | 0.04 | 0.07 | 0.65 | 3496.36 | 0.52 |
| **fclrg90.C** | -0.05 | 0.11 | -0.43 | 1840.34 | 0.66 |
| **fclrg90^4** | -0.02 | 0.08 | -0.26 | 1951.95 | 0.80 |
| **tenure.L** | 0.05 | 0.06 | 0.75 | 1839.19 | 0.45 |
| **tenure.Q** | -0.05 | 0.09 | -0.55 | 1999.40 | 0.58 |
| **divorce1** | 0.02 | 0.07 | 0.23 | 3325.00 | 0.82 |
| **sepmumbcs1** | 0.32 | 0.13 | 2.39 | 2665.13 | 0.02 |
| **prmnh1** | 0.32 | 0.15 | 2.19 | 1721.06 | 0.03 |
| **crowdUp to 1** | -0.10 | 0.07 | -1.41 | 2994.98 | 0.16 |
| **ameniNo occasions** | 0.01 | 0.11 | 0.14 | 3869.46 | 0.89 |
| **brfed.L** | -0.01 | 0.06 | -0.13 | 2223.13 | 0.89 |
| **brfed.Q** | 0.00 | 0.10 | -0.04 | 1371.76 | 0.97 |
| **resmove.L** | 0.21 | 0.09 | 2.42 | 1737.16 | 0.02 |
| **resmove.Q** | 0.06 | 0.04 | 1.39 | 3685.54 | 0.17 |

*Table S74.* Raw output from modified Poisson regression (with log link) of externalising behaviour at age 16 (extbcsz) against median sleep duration (binarised to normal vs abnormal) derived from the sleep diary at age 46, adjusted for potential confounders (see main text Table 3) and mental health variables at age 42 (see Table S42). The parameter estimates in the “Estimate” column and the associated standard errors have not been exponentiated.

| Term | Estimate | Standard error | Statistic | Degrees of freedom | p-value |
|---|---|---|---|---|---|
| **(Intercept)** | -0.42 | 0.25 | -1.69 | 2383.25 | 0.09 |
| **extbcsz** | 0.01 | 0.01 | 0.57 | 4138.69 | 0.57 |
| **BD9MAL** | 0.12 | 0.10 | 1.27 | 2163.55 | 0.20 |
| **BD9WEMWB** | 0.05 | 0.18 | 0.31 | 1651.34 | 0.76 |
| **a0005a** | -0.35 | 0.10 | -3.54 | 4958.28 | 0.00 |
| **a0043b.L** | -0.05 | 0.05 | -1.03 | 1775.35 | 0.30 |
| **a0043b.Q** | -0.02 | 0.06 | -0.40 | 2090.22 | 0.69 |
| **a0043b.C** | 0.00 | 0.06 | -0.01 | 1657.66 | 0.99 |
| **a0043b^4** | -0.06 | 0.04 | -1.30 | 3615.93 | 0.19 |
| **a0043b^5** | -0.01 | 0.07 | -0.19 | 2122.80 | 0.85 |
| **a0195a** | -0.39 | 0.46 | -0.84 | 1682.96 | 0.40 |
| **a0278** | -0.20 | 0.20 | -1.03 | 2429.18 | 0.30 |
| **a0014.L** | 0.01 | 0.07 | 0.20 | 1952.09 | 0.84 |
| **a0014.Q** | 0.03 | 0.05 | 0.55 | 1955.30 | 0.58 |
| **a0014.C** | 0.00 | 0.05 | 0.06 | 2568.38 | 0.95 |
| **a0014^4** | -0.01 | 0.04 | -0.26 | 1779.51 | 0.79 |
| **dv_par_edu_birth** | -0.38 | 0.46 | -0.82 | 2667.05 | 0.41 |
| **dv_sc_age_5.L** | -0.02 | 0.06 | -0.26 | 1522.82 | 0.80 |
| **dv_sc_age_5.Q** | 0.00 | 0.03 | -0.16 | 2947.82 | 0.87 |
| **e216aNone** | -0.01 | 0.05 | -0.11 | 1549.45 | 0.91 |
| **dv_cog_abil_5** | 0.04 | 0.28 | 0.16 | 858.47 | 0.87 |
| **dv_med_51** | 0.00 | 0.04 | 0.11 | 2850.13 | 0.92 |
| **d016a** | 0.06 | 0.03 | 1.68 | 3495.50 | 0.09 |
| **dv_bas_g** | -0.22 | 0.16 | -1.39 | 1848.52 | 0.17 |
| **dv_med_101** | 0.01 | 0.03 | 0.26 | 2812.43 | 0.79 |
| **bmi** | 0.19 | 0.17 | 1.09 | 2166.68 | 0.28 |
| **fclrg90.L** | 0.02 | 0.06 | 0.38 | 2530.62 | 0.71 |
| **fclrg90.Q** | 0.04 | 0.04 | 1.04 | 2462.96 | 0.30 |
| **fclrg90.C** | 0.06 | 0.04 | 1.50 | 3954.20 | 0.13 |
| **fclrg90^4** | 0.01 | 0.05 | 0.27 | 1785.43 | 0.79 |
| **tenure.L** | 0.01 | 0.03 | 0.45 | 2417.04 | 0.66 |
| **tenure.Q** | -0.03 | 0.05 | -0.70 | 2258.95 | 0.48 |
| **divorce1** | 0.03 | 0.04 | 0.71 | 4142.11 | 0.48 |
| **sepmumbcs1** | -0.06 | 0.09 | -0.73 | 3033.68 | 0.47 |
| **prmnh1** | 0.17 | 0.09 | 1.81 | 1789.20 | 0.07 |
| **crowdUp to 1** | -0.05 | 0.05 | -0.87 | 1883.99 | 0.38 |
| **ameniNo occasions** | -0.08 | 0.07 | -1.07 | 2170.41 | 0.29 |
| **brfed.L** | 0.03 | 0.03 | 0.82 | 2121.65 | 0.41 |
| **brfed.Q** | -0.01 | 0.03 | -0.41 | 3474.20 | 0.68 |
| **resmove.L** | -0.03 | 0.04 | -0.74 | 2837.74 | 0.46 |
| **resmove.Q** | 0.06 | 0.02 | 2.50 | 4296.23 | 0.01 |

*Table S75.* Raw output from modified Poisson regression (with log link) of externalising behaviour at age 16 (extbcsz) against median sleep duration (binarised to normal vs abnormal) derived from the activPAL algorithm at age 46, adjusted for potential confounders (see main text Table 3) and mental health variables at age 42 (see Table S42). The parameter estimates in the "Estimate" column and the associated standard errors have not been exponentiated.

| Term | Estimate | Standard error | Statistic | Degrees of freedom | p-value |
|---|---|---|---|---|---|
| **(Intercept)** | -0.89 | 0.38 | -2.35 | 1445.44 | 0.02 |
| **extbcsz** | 0.03 | 0.03 | 0.98 | 1215.66 | 0.33 |
| **BD9MAL** | 0.17 | 0.12 | 1.41 | 1810.49 | 0.16 |
| **BD9WEMWB** | -0.13 | 0.18 | -0.72 | 1823.63 | 0.47 |
| **a0005a** | -0.01 | 0.17 | -0.03 | 1766.08 | 0.97 |
| **a0043b.L** | 0.00 | 0.06 | 0.01 | 1607.54 | 0.99 |
| **a0043b.Q** | -0.07 | 0.07 | -0.99 | 1806.79 | 0.32 |
| **a0043b.C** | -0.18 | 0.07 | -2.80 | 1719.30 | 0.01 |
| **a0043b^4** | 0.10 | 0.06 | 1.65 | 2053.38 | 0.10 |
| **a0043b^5** | 0.03 | 0.08 | 0.42 | 2140.66 | 0.67 |
| **a0195a** | -0.26 | 0.47 | -0.56 | 1976.75 | 0.58 |
| **a0278** | 0.36 | 0.18 | 2.03 | 3940.72 | 0.04 |
| **a0014.L** | 0.03 | 0.09 | 0.30 | 1534.13 | 0.77 |
| **a0014.Q** | 0.04 | 0.07 | 0.61 | 1547.76 | 0.54 |
| **a0014.C** | -0.01 | 0.05 | -0.16 | 2662.44 | 0.87 |
| **a0014^4** | -0.02 | 0.04 | -0.48 | 2367.20 | 0.63 |
| **dv_par_edu_birth** | -0.10 | 0.41 | -0.24 | 3860.74 | 0.81 |
| **dv_sc_age_5.L** | 0.15 | 0.08 | 1.79 | 1160.76 | 0.07 |
| **dv_sc_age_5.Q** | -0.03 | 0.04 | -0.94 | 2193.47 | 0.35 |
| **e216aNone** | 0.07 | 0.05 | 1.42 | 1834.86 | 0.16 |
| **dv_cog_abil_5** | 0.01 | 0.28 | 0.05 | 1010.72 | 0.96 |
| **dv_med_51** | 0.04 | 0.05 | 0.75 | 1872.36 | 0.46 |
| **d016a** | 0.08 | 0.04 | 1.86 | 2402.92 | 0.06 |
| **dv_bas_g** | -0.37 | 0.22 | -1.67 | 1270.52 | 0.10 |
| **dv_med_101** | 0.04 | 0.05 | 0.76 | 1545.53 | 0.45 |
| **bmi** | -0.25 | 0.24 | -1.05 | 1546.02 | 0.29 |
| **fclrg90.L** | -0.01 | 0.07 | -0.21 | 2540.94 | 0.84 |
| **fclrg90.Q** | -0.03 | 0.06 | -0.47 | 1667.89 | 0.64 |
| **fclrg90.C** | -0.10 | 0.06 | -1.79 | 2168.18 | 0.07 |
| **fclrg90^4** | 0.01 | 0.05 | 0.20 | 2085.40 | 0.84 |
| **tenure.L** | 0.03 | 0.05 | 0.64 | 1369.22 | 0.52 |
| **tenure.Q** | 0.09 | 0.06 | 1.60 | 2234.21 | 0.11 |
| **divorce1** | 0.13 | 0.05 | 2.77 | 2631.19 | 0.01 |
| **sepmumbcs1** | -0.12 | 0.14 | -0.83 | 1575.98 | 0.41 |
| **prmnh1** | -0.24 | 0.19 | -1.29 | 1121.71 | 0.20 |
| **crowdUp to 1** | 0.03 | 0.06 | 0.54 | 1834.14 | 0.59 |
| **ameniNo occasions** | -0.08 | 0.08 | -1.09 | 2424.25 | 0.27 |
| **brfed.L** | 0.01 | 0.04 | 0.23 | 1612.67 | 0.82 |
| **brfed.Q** | 0.04 | 0.05 | 0.93 | 2262.06 | 0.35 |
| **resmove.L** | 0.00 | 0.04 | 0.06 | 3607.96 | 0.95 |
| **resmove.Q** | -0.06 | 0.03 | -2.20 | 3689.76 | 0.03 |

*Table S76.* Raw output from modified Poisson regression (with log link) of externalising behaviour at age 16 (extbcsz) against median sleep duration (binarised to normal vs abnormal) derived from the van der Berg et al. algorithm at age 46, adjusted for potential confounders (see main text Table 3) and mental health variables at age 42 (see Table S42). The parameter estimates in the “Estimate” column and the associated standard errors have not been exponentiated.

| Term | Estimate | Standard error | Statistic | Degrees of freedom | p-value |
|---|---|---|---|---|---|
| **(Intercept)** | -0.98 | 0.38 | -2.59 | 1316.01 | 0.01 |
| **extbcsz** | 0.06 | 0.02 | 2.73 | 1242.33 | 0.01 |
| **BD9MAL** | 0.25 | 0.13 | 1.95 | 1442.21 | 0.05 |
| **BD9WEMWB** | -0.08 | 0.24 | -0.32 | 1135.36 | 0.75 |
| **a0005a** | 0.16 | 0.20 | 0.80 | 1255.66 | 0.42 |
| **a0043b.L** | -0.05 | 0.04 | -1.31 | 2945.18 | 0.19 |
| **a0043b.Q** | 0.02 | 0.06 | 0.28 | 2134.98 | 0.78 |
| **a0043b.C** | 0.06 | 0.04 | 1.59 | 4802.94 | 0.11 |
| **a0043b^4** | 0.03 | 0.05 | 0.54 | 2667.82 | 0.59 |
| **a0043b^5** | 0.04 | 0.07 | 0.51 | 2450.67 | 0.61 |
| **a0195a** | -0.06 | 0.33 | -0.18 | 3957.40 | 0.86 |
| **a0278** | -0.09 | 0.27 | -0.34 | 1581.61 | 0.73 |
| **a0014.L** | 0.07 | 0.07 | 0.98 | 2113.23 | 0.33 |
| **a0014.Q** | 0.01 | 0.06 | 0.12 | 1554.38 | 0.91 |
| **a0014.C** | -0.03 | 0.09 | -0.39 | 1084.93 | 0.70 |
| **a0014^4** | 0.03 | 0.05 | 0.62 | 1516.36 | 0.54 |
| **dv_par_edu_birth** | 0.47 | 0.42 | 1.13 | 1884.06 | 0.26 |
| **dv_sc_age_5.L** | 0.01 | 0.09 | 0.15 | 1006.33 | 0.88 |
| **dv_sc_age_5.Q** | -0.01 | 0.05 | -0.17 | 1438.98 | 0.86 |
| **e216aNone** | -0.12 | 0.05 | -2.21 | 1399.59 | 0.03 |
| **dv_cog_abil_5** | -0.22 | 0.27 | -0.83 | 985.59 | 0.41 |
| **dv_med_51** | 0.02 | 0.04 | 0.46 | 2599.07 | 0.65 |
| **d016a** | 0.18 | 0.06 | 3.26 | 1369.51 | 0.00 |
| **dv_bas_g** | -0.20 | 0.18 | -1.08 | 1559.47 | 0.28 |
| **dv_med_101** | 0.03 | 0.05 | 0.55 | 1579.72 | 0.58 |
| **bmi** | 0.53 | 0.21 | 2.56 | 1638.44 | 0.01 |
| **fclrg90.L** | -0.13 | 0.08 | -1.60 | 1601.16 | 0.11 |
| **fclrg90.Q** | -0.06 | 0.06 | -0.97 | 1812.02 | 0.33 |
| **fclrg90.C** | 0.02 | 0.07 | 0.35 | 1508.85 | 0.72 |
| **fclrg90^4** | 0.01 | 0.05 | 0.10 | 1717.11 | 0.92 |
| **tenure.L** | 0.03 | 0.04 | 0.77 | 1468.10 | 0.44 |
| **tenure.Q** | -0.05 | 0.06 | -0.77 | 1582.91 | 0.44 |
| **divorce1** | -0.03 | 0.06 | -0.53 | 1570.27 | 0.60 |
| **sepmumbcs1** | 0.19 | 0.10 | 1.95 | 1932.92 | 0.05 |
| **prmnh1** | -0.08 | 0.11 | -0.77 | 2074.94 | 0.44 |
| **crowdUp to 1** | 0.08 | 0.05 | 1.63 | 2270.63 | 0.10 |
| **ameniNo occasions** | -0.16 | 0.10 | -1.71 | 1506.39 | 0.09 |
| **brfed.L** | 0.03 | 0.04 | 0.89 | 2244.40 | 0.37 |
| **brfed.Q** | -0.05 | 0.05 | -0.91 | 1600.30 | 0.36 |
| **resmove.L** | 0.02 | 0.06 | 0.34 | 1500.23 | 0.73 |
| **resmove.Q** | 0.02 | 0.02 | 1.02 | 4992.29 | 0.31 |

*Table S77.* Raw output from modified Poisson regression (with log link) of externalising behaviour at age 16 (extbcsz) against median sleep duration (binarised to normal vs abnormal) derived from the Winkler et al. algorithm at age 46, adjusted for potential confounders (see main text Table 3) and mental health variables at age 42 (see Table S42). The parameter estimates in the "Estimate" column and the associated standard errors have not been exponentiated.

### Results disaggregated by sex assigned at birth: Figures S2–7

The following pages contain forest plots detailing the results of regression analyses disaggregated by sex assigned at birth. The figures are as follows:

| **Sex assigned at birth** | **Initial Poisson regressions** | **Poisson mediation regressions** | **Multinomial regressions** |
|---|---|---|---|
| **Both** | Figure 1 (main text) | Figure 2 (main text) | Figure S1 (above) |
| **Male** | Figure S2 | Figure S3 | Figure S4 |
| **Female** | Figure S5 | Figure S6 | Figure S7 |

#### Self-reported average

| Variable (age at collection) | RR | p | E-value |
|---|---|---|---|
| Rutter score (5) | 1.104 | 0.002 | 1.230 |
| Rutter score (10) | 1.068 | 0.079 | 1.000 |
| Child Development Scale (10) | 1.161 | 0.002 | 1.308 |
| Malaise score (16)* | 1.203 | 0.000 | 1.446 |
| Behavioural & emotional problems (16) | 1.094 | 0.000 | 1.279 |
| Internalising behaviours (16) | 1.087 | 0.023 | 1.120 |
| Externalising behaviours (16) | 1.132 | 0.000 | 1.328 |

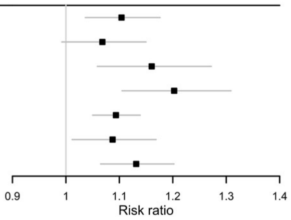

#### Diary-derived average

| Variable (age at collection) | RR | p | E-value |
|---|---|---|---|
| Rutter score (5) | 1.071 | 0.046 | 1.035 |
| Rutter score (10) | 1.086 | 0.010 | 1.164 |
| Child Development Scale (10) | 1.127 | 0.001 | 1.271 |
| Malaise score (16)* | 1.108 | 0.006 | 1.204 |
| Behavioural & emotional problems (16) | 1.065 | 0.045 | 1.040 |
| Internalising behaviours (16) | 1.083 | 0.004 | 1.189 |
| Externalising behaviours (16) | 1.069 | 0.090 | 1.000 |

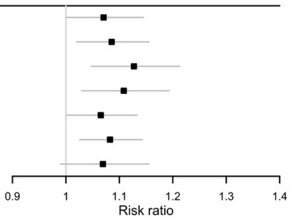

#### activPAL algorithm

| Variable (age at collection) | RR | p | E-value |
|---|---|---|---|
| Rutter score (5) | 0.992 | 0.742 | 1.000 |
| Rutter score (10) | 1.022 | 0.366 | 1.000 |
| Child Development Scale (10) | 0.992 | 0.706 | 1.000 |
| Malaise score (16)* | 0.994 | 0.839 | 1.000 |
| Behavioural & emotional problems (16) | 1.018 | 0.505 | 1.000 |
| Internalising behaviours (16) | 1.016 | 0.583 | 1.000 |
| Externalising behaviours (16) | 1.010 | 0.573 | 1.000 |

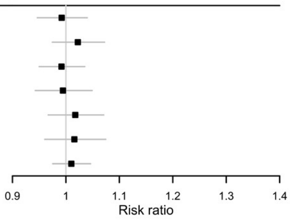

#### van der Berg et al. algorithm

| Variable (age at collection) | RR | p | E-value |
|---|---|---|---|
| Rutter score (5) | 1.025 | 0.248 | 1.000 |
| Rutter score (10) | 1.036 | 0.146 | 1.000 |
| Child Development Scale (10) | 1.022 | 0.424 | 1.000 |
| Malaise score (16)* | 1.004 | 0.882 | 1.000 |
| Behavioural & emotional problems (16) | 0.958 | 0.062 | 1.000 |
| Internalising behaviours (16) | 1.012 | 0.736 | 1.000 |
| Externalising behaviours (16) | 1.034 | 0.314 | 1.000 |

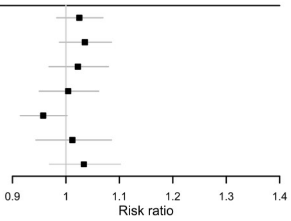

#### Winkler et al. algorithm

| Variable (age at collection) | RR | p | E-value |
|---|---|---|---|
| Rutter score (5) | 1.082 | 0.001 | 1.223 |
| Rutter score (10) | 1.029 | 0.199 | 1.000 |
| Child Development Scale (10) | 1.090 | 0.002 | 1.216 |
| Malaise score (16)* | 1.075 | 0.000 | 1.216 |
| Behavioural & emotional problems (16) | 1.013 | 0.568 | 1.000 |
| Internalising behaviours (16) | 1.031 | 0.241 | 1.000 |
| Externalising behaviours (16) | 1.072 | 0.001 | 1.194 |

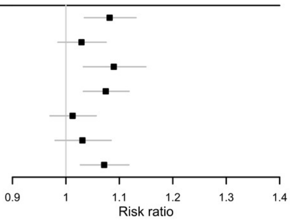

*Figure S2.* Estimated risk ratios quantifying the associations among male participants between the childhood mental health variables (standardised) and the presence of abnormal sleep duration in adulthood (derived from five separate measures of sleep). The E-value, quantifying the minimum unmeasured confounding risk ratio that would be needed to nullify each association estimated here, is also shown. Each exposure-outcome relationship was assessed in a separate model. All models were adjusted for a range of socioeconomic, perinatal and health-related covariates. Missing data were handled by multiple imputation. RR = risk ratio. * = self-reported.

**Self-reported average**

| Variable (age at collection) | RR | p | E-value |
|---|---|---|---|
| Rutter score (5) | 1.104 | 0.002 | 1.230 |
| (adjusted for age 42 mental health) | 1.047 | 0.124 | 1.000 |
| Rutter score (10) | 1.068 | 0.079 | 1.000 |
| (adjusted for age 42 mental health) | 1.013 | 0.730 | 1.000 |
| Child Development Scale (10) | 1.161 | 0.002 | 1.308 |
| (adjusted for age 42 mental health) | 1.114 | 0.011 | 1.185 |
| Malaise score (16)* | 1.203 | 0.000 | 1.446 |
| (adjusted for age 42 mental health) | 1.074 | 0.129 | 1.000 |
| Behavioural & emotional problems (16) | 1.094 | 0.000 | 1.279 |
| (adjusted for age 42 mental health) | 1.066 | 0.002 | 1.180 |
| Internalising behaviours (16) | 1.087 | 0.023 | 1.120 |
| (adjusted for age 42 mental health) | 1.002 | 0.965 | 1.000 |
| Externalising behaviours (16) | 1.132 | 0.000 | 1.328 |
| (adjusted for age 42 mental health) | 1.070 | 0.026 | 1.098 |

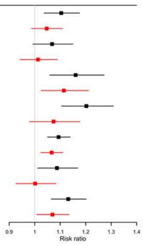

**Diary-derived average**

| Variable (age at collection) | RR | p | E-value |
|---|---|---|---|
| Rutter score (5) | 1.071 | 0.046 | 1.035 |
| (adjusted for age 42 mental health) | 1.037 | 0.299 | 1.000 |
| Rutter score (10) | 1.086 | 0.010 | 1.164 |
| (adjusted for age 42 mental health) | 1.052 | 0.104 | 1.000 |
| Child Development Scale (10) | 1.127 | 0.001 | 1.271 |
| (adjusted for age 42 mental health) | 1.102 | 0.011 | 1.177 |
| Malaise score (16)* | 1.108 | 0.006 | 1.204 |
| (adjusted for age 42 mental health) | 1.040 | 0.350 | 1.000 |
| Behavioural & emotional problems (16) | 1.065 | 0.045 | 1.040 |
| (adjusted for age 42 mental health) | 1.051 | 0.104 | 1.000 |
| Internalising behaviours (16) | 1.083 | 0.004 | 1.189 |
| (adjusted for age 42 mental health) | 1.036 | 0.219 | 1.000 |
| Externalising behaviours (16) | 1.069 | 0.090 | 1.000 |
| (adjusted for age 42 mental health) | 1.035 | 0.395 | 1.000 |

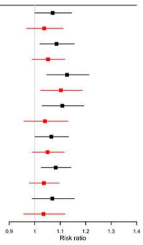

**activPAL algorithm**

| Variable (age at collection) | RR | p | E-value |
|---|---|---|---|
| Rutter score (5) | 0.992 | 0.742 | 1.000 |
| (adjusted for age 42 mental health) | 0.990 | 0.660 | 1.000 |
| Rutter score (10) | 1.022 | 0.366 | 1.000 |
| (adjusted for age 42 mental health) | 1.020 | 0.422 | 1.000 |
| Child Development Scale (10) | 0.992 | 0.706 | 1.000 |
| (adjusted for age 42 mental health) | 0.989 | 0.646 | 1.000 |
| Malaise score (16)* | 0.994 | 0.839 | 1.000 |
| (adjusted for age 42 mental health) | 0.988 | 0.685 | 1.000 |
| Behavioural & emotional problems (16) | 1.018 | 0.505 | 1.000 |
| (adjusted for age 42 mental health) | 1.016 | 0.527 | 1.000 |
| Internalising behaviours (16) | 1.016 | 0.583 | 1.000 |
| (adjusted for age 42 mental health) | 1.013 | 0.667 | 1.000 |
| Externalising behaviours (16) | 1.010 | 0.573 | 1.000 |
| (adjusted for age 42 mental health) | 1.008 | 0.680 | 1.000 |

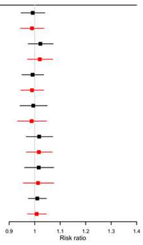

**van der Berg et al. algorithm**

| Variable (age at collection) | RR | p | E-value |
|---|---|---|---|
| Rutter score (5) | 1.025 | 0.248 | 1.000 |
| (adjusted for age 42 mental health) | 1.015 | 0.500 | 1.000 |
| Rutter score (10) | 1.036 | 0.146 | 1.000 |
| (adjusted for age 42 mental health) | 1.026 | 0.302 | 1.000 |
| Child Development Scale (10) | 1.022 | 0.424 | 1.000 |
| (adjusted for age 42 mental health) | 1.015 | 0.591 | 1.000 |
| Malaise score (16)* | 1.004 | 0.882 | 1.000 |
| (adjusted for age 42 mental health) | 0.979 | 0.483 | 1.000 |
| Behavioural & emotional problems (16) | 0.958 | 0.062 | 1.000 |
| (adjusted for age 42 mental health) | 0.953 | 0.030 | 1.073 |
| Internalising behaviours (16) | 1.012 | 0.736 | 1.000 |
| (adjusted for age 42 mental health) | 0.997 | 0.936 | 1.000 |
| Externalising behaviours (16) | 1.034 | 0.314 | 1.000 |
| (adjusted for age 42 mental health) | 1.023 | 0.527 | 1.000 |

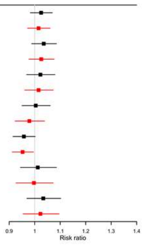

**Winkler et al. algorithm**

| Variable (age at collection) | RR | p | E-value |
|---|---|---|---|
| Rutter score (5) | 1.082 | 0.001 | 1.223 |
| (adjusted for age 42 mental health) | 1.073 | 0.004 | 1.176 |
| Rutter score (10) | 1.029 | 0.199 | 1.000 |
| (adjusted for age 42 mental health) | 1.021 | 0.365 | 1.000 |
| Child Development Scale (10) | 1.090 | 0.002 | 1.216 |
| (adjusted for age 42 mental health) | 1.083 | 0.008 | 1.169 |
| Malaise score (16)* | 1.075 | 0.000 | 1.216 |
| (adjusted for age 42 mental health) | 1.056 | 0.013 | 1.119 |
| Behavioural & emotional problems (16) | 1.013 | 0.568 | 1.000 |
| (adjusted for age 42 mental health) | 1.008 | 0.725 | 1.000 |
| Internalising behaviours (16) | 1.031 | 0.241 | 1.000 |
| (adjusted for age 42 mental health) | 1.017 | 0.554 | 1.000 |
| Externalising behaviours (16) | 1.072 | 0.001 | 1.194 |
| (adjusted for age 42 mental health) | 1.062 | 0.005 | 1.153 |

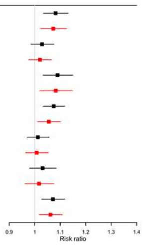

*Figure S3.* Estimated risk ratios quantifying the associations among male participants between the childhood mental health variables (standardised) and the status of sleep duration as abnormally short, normal or abnormally long in adulthood (derived from five separate measures of sleep), also including (in red) those adjusting for Malaise and Warwick–Edinburgh Mental Well-Being Scale scores at age 42. The E-value, quantifying the minimum unmeasured confounding risk ratio that would be needed to nullify each association estimated here, is also shown. Each exposure-outcome relationship was assessed in a separate model. All models were adjusted for a range of socioeconomic, perinatal and health-related covariates. Missing data were handled by multiple imputation. RR = risk ratio. * = self-reported.

### Abnormally short sleep duration

**Self-reported average**

| Variable (age at collection) | RRR | p | E-value |
|---|---|---|---|
| Rutter score (5) | 1.137 | 0.003 | 1.267 |
| Rutter score (10) | 1.076 | 0.134 | 1.000 |
| Child Development Scale (10) | 1.195 | 0.003 | 1.323 |
| Malaise score (16)* | 1.264 | 0.000 | 1.497 |
| Behavioural & emotional problems (16) | 1.109 | 0.006 | 1.207 |
| Internalising behaviours (16) | 1.086 | 0.096 | 1.000 |
| Externalising behaviours (16) | 1.152 | 0.001 | 1.304 |

**Diary-derived average**

| Variable (age at collection) | RRR | p | E-value |
|---|---|---|---|
| Rutter score (5) | 1.039 | 0.398 | 1.000 |
| Rutter score (10) | 1.067 | 0.164 | 1.000 |
| Child Development Scale (10) | 1.142 | 0.073 | 1.000 |
| Malaise score (16)* | 1.127 | 0.119 | 1.000 |
| Behavioural & emotional problems (16) | 1.091 | 0.185 | 1.000 |
| Internalising behaviours (16) | 1.064 | 0.396 | 1.000 |
| Externalising behaviours (16) | 1.053 | 0.288 | 1.000 |

**activPAL algorithm**

| Variable (age at collection) | RRR | p | E-value |
|---|---|---|---|
| Rutter score (5) | 0.989 | 0.868 | 1.000 |
| Rutter score (10) | 1.024 | 0.727 | 1.000 |
| Child Development Scale (10) | 0.969 | 0.669 | 1.000 |
| Malaise score (16)* | 1.043 | 0.543 | 1.000 |
| Behavioural & emotional problems (16) | 1.084 | 0.194 | 1.000 |
| Internalising behaviours (16) | 1.020 | 0.729 | 1.000 |
| Externalising behaviours (16) | 1.016 | 0.797 | 1.000 |

**van der Berg et al. algorithm**

| Variable (age at collection) | RRR | p | E-value |
|---|---|---|---|
| Rutter score (5) | 1.052 | 0.537 | 1.000 |
| Rutter score (10) | 1.073 | 0.438 | 1.000 |
| Child Development Scale (10) | 1.096 | 0.380 | 1.000 |
| Malaise score (16)* | 1.087 | 0.358 | 1.000 |
| Behavioural & emotional problems (16) | 0.924 | 0.835 | 1.000 |
| Internalising behaviours (16) | 1.071 | 0.506 | 1.000 |
| Externalising behaviours (16) | 1.091 | 0.357 | 1.000 |

**Winkler et al. algorithm**

| Variable (age at collection) | RRR | p | E-value |
|---|---|---|---|
| Rutter score (5) | 1.115 | 0.026 | 1.131 |
| Rutter score (10) | 1.030 | 0.454 | 1.000 |
| Child Development Scale (10) | 1.159 | 0.001 | 1.313 |
| Malaise score (16)* | 1.114 | 0.006 | 1.210 |
| Behavioural & emotional problems (16) | 1.009 | 0.819 | 1.000 |
| Internalising behaviours (16) | 1.052 | 0.301 | 1.000 |
| Externalising behaviours (16) | 1.111 | 0.004 | 1.227 |

### Abnormally long sleep duration

**Self-reported average**

| Variable (age at collection) | RRR | p | E-value |
|---|---|---|---|
| Rutter score (5) | 1.122 | 0.453 | 1.000 |
| Rutter score (10) | 1.014 | 0.939 | 1.000 |
| Child Development Scale (10) | 1.110 | 0.613 | 1.000 |
| Malaise score (16)* | 1.022 | 0.924 | 1.000 |
| Behavioural & emotional problems (16) | 1.175 | 0.067 | 1.000 |
| Internalising behaviours (16) | 1.210 | 0.331 | 1.000 |
| Externalising behaviours (16) | 1.196 | 0.161 | 1.000 |

**Diary-derived average**

| Variable (age at collection) | RRR | p | E-value |
|---|---|---|---|
| Rutter score (5) | 1.073 | 0.391 | 1.000 |
| Rutter score (10) | 1.039 | 0.662 | 1.000 |
| Child Development Scale (10) | 1.126 | 0.133 | 1.000 |
| Malaise score (16)* | 1.070 | 0.630 | 1.000 |
| Behavioural & emotional problems (16) | 1.009 | 0.932 | 1.000 |
| Internalising behaviours (16) | 1.017 | 0.846 | 1.000 |
| Externalising behaviours (16) | 1.009 | 0.904 | 1.000 |

**activPAL algorithm**

| Variable (age at collection) | RRR | p | E-value |
|---|---|---|---|
| Rutter score (5) | 0.993 | 0.859 | 1.000 |
| Rutter score (10) | 1.030 | 0.481 | 1.000 |
| Child Development Scale (10) | 0.991 | 0.805 | 1.000 |
| Malaise score (16)* | 1.016 | 0.734 | 1.000 |
| Behavioural & emotional problems (16) | 1.033 | 0.430 | 1.000 |
| Internalising behaviours (16) | 1.012 | 0.761 | 1.000 |
| Externalising behaviours (16) | 1.005 | 0.907 | 1.000 |

**van der Berg et al. algorithm**

| Variable (age at collection) | RRR | p | E-value |
|---|---|---|---|
| Rutter score (5) | 1.038 | 0.202 | 1.000 |
| Rutter score (10) | 1.060 | 0.244 | 1.000 |
| Child Development Scale (10) | 1.014 | 0.806 | 1.000 |
| Malaise score (16)* | 1.007 | 0.905 | 1.000 |
| Behavioural & emotional problems (16) | 0.990 | 0.863 | 1.000 |
| Internalising behaviours (16) | 1.053 | 0.366 | 1.000 |
| Externalising behaviours (16) | 1.056 | 0.298 | 1.000 |

**Winkler et al. algorithm**

| Variable (age at collection) | RRR | p | E-value |
|---|---|---|---|
| Rutter score (5) | 1.066 | 0.423 | 1.000 |
| Rutter score (10) | 0.995 | 0.962 | 1.000 |
| Child Development Scale (10) | 0.995 | 0.954 | 1.000 |
| Malaise score (16)* | 0.949 | 0.641 | 1.000 |
| Behavioural & emotional problems (16) | 1.011 | 0.872 | 1.000 |
| Internalising behaviours (16) | 0.990 | 0.897 | 1.000 |
| Externalising behaviours (16) | 0.992 | 0.913 | 1.000 |

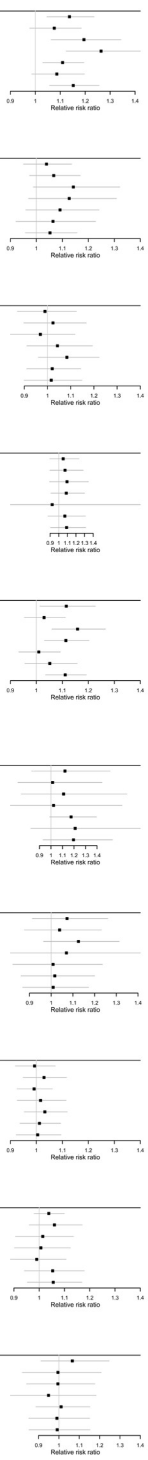

*Figure S4.* Estimated relative risk ratios quantifying the associations among male participants between the childhood mental health variables (standardised) and the presence of abnormally short, and abnormally long, sleep duration in adulthood (derived from five separate measures of sleep). The E-value, quantifying the minimum unmeasured confounding risk ratio that would be needed to nullify each association estimated here, is also shown. Each exposure-outcome relationship was assessed in a separate model. All models were adjusted for a range of socioeconomic, perinatal and health-related covariates. Missing data were handled by multiple imputation. RRR = relative risk ratio. * = self-reported.

### Self-reported average

| Variable (age at collection) | RR | p | E-value |
|---|---|---|---|
| Rutter score (5) | 1.109 | 0.002 | 1.237 |
| Rutter score (10) | 1.098 | 0.004 | 1.204 |
| Child Development Scale (10) | 1.177 | 0.000 | 1.405 |
| Malaise score (16)* | 1.168 | 0.000 | 1.392 |
| Behavioural & emotional problems (16) | 1.066 | 0.022 | 1.106 |
| Internalising behaviours (16) | 1.054 | 0.147 | 1.000 |
| Externalising behaviours (16) | 1.107 | 0.023 | 1.133 |

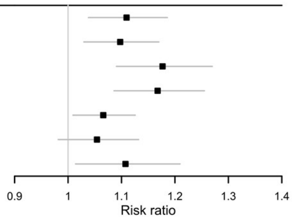

### Diary-derived average

| Variable (age at collection) | RR | p | E-value |
|---|---|---|---|
| Rutter score (5) | 1.119 | 0.022 | 1.147 |
| Rutter score (10) | 1.139 | 0.002 | 1.272 |
| Child Development Scale (10) | 1.145 | 0.007 | 1.238 |
| Malaise score (16)* | 1.096 | 0.041 | 1.064 |
| Behavioural & emotional problems (16) | 1.062 | 0.152 | 1.000 |
| Internalising behaviours (16) | 1.070 | 0.076 | 1.000 |
| Externalising behaviours (16) | 1.079 | 0.133 | 1.000 |

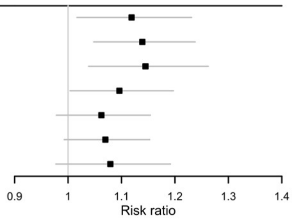

### activPAL algorithm

| Variable (age at collection) | RR | p | E-value |
|---|---|---|---|
| Rutter score (5) | 0.998 | 0.943 | 1.000 |
| Rutter score (10) | 1.016 | 0.425 | 1.000 |
| Child Development Scale (10) | 0.992 | 0.730 | 1.000 |
| Malaise score (16)* | 0.999 | 0.982 | 1.000 |
| Behavioural & emotional problems (16) | 1.032 | 0.096 | 1.000 |
| Internalising behaviours (16) | 1.016 | 0.396 | 1.000 |
| Externalising behaviours (16) | 1.016 | 0.430 | 1.000 |

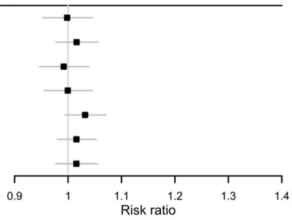

### van der Berg et al. algorithm

| Variable (age at collection) | RR | p | E-value |
|---|---|---|---|
| Rutter score (5) | 1.022 | 0.352 | 1.000 |
| Rutter score (10) | 1.016 | 0.580 | 1.000 |
| Child Development Scale (10) | 1.013 | 0.699 | 1.000 |
| Malaise score (16)* | 0.998 | 0.929 | 1.000 |
| Behavioural & emotional problems (16) | 0.967 | 0.432 | 1.000 |
| Internalising behaviours (16) | 1.007 | 0.811 | 1.000 |
| Externalising behaviours (16) | 1.032 | 0.265 | 1.000 |

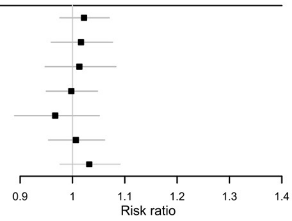

### Winkler et al. algorithm

| Variable (age at collection) | RR | p | E-value |
|---|---|---|---|
| Rutter score (5) | 1.086 | 0.001 | 1.218 |
| Rutter score (10) | 1.016 | 0.480 | 1.000 |
| Child Development Scale (10) | 1.099 | 0.004 | 1.211 |
| Malaise score (16)* | 1.086 | 0.003 | 1.200 |
| Behavioural & emotional problems (16) | 1.012 | 0.586 | 1.000 |
| Internalising behaviours (16) | 1.027 | 0.314 | 1.000 |
| Externalising behaviours (16) | 1.077 | 0.025 | 1.106 |

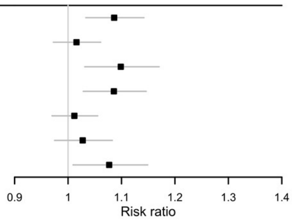

*Figure S5.* Estimated risk ratios quantifying the associations among female participants between the childhood mental health variables (standardised) and the presence of abnormal sleep duration in adulthood (derived from five separate measures of sleep). The E-value, quantifying the minimum unmeasured confounding risk ratio that would be needed to nullify each association estimated here, is also shown. Each exposure-outcome relationship was assessed in a separate model. All models were adjusted for a range of socioeconomic, perinatal and health-related covariates. Missing data were handled by multiple imputation. RR = risk ratio. * = self-reported.

**Self-reported average**

| Variable (age at collection) | RR | p | E-value |
|---|---|---|---|
| Rutter score (5) | 1.109 | 0.002 | 1.237 |
| (adjusted for age 42 mental health) | 1.051 | 0.141 | 1.000 |
| Rutter score (10) | 1.098 | 0.004 | 1.204 |
| (adjusted for age 42 mental health) | 1.030 | 0.358 | 1.000 |
| Child Development Scale (10) | 1.177 | 0.000 | 1.405 |
| (adjusted for age 42 mental health) | 1.134 | 0.001 | 1.291 |
| Malaise score (16)* | 1.168 | 0.000 | 1.392 |
| (adjusted for age 42 mental health) | 1.061 | 0.121 | 1.000 |
| Behavioural & emotional problems (16) | 1.066 | 0.022 | 1.106 |
| (adjusted for age 42 mental health) | 1.041 | 0.136 | 1.000 |
| Internalising behaviours (16) | 1.054 | 0.147 | 1.000 |
| (adjusted for age 42 mental health) | 0.979 | 0.566 | 1.000 |
| Externalising behaviours (16) | 1.107 | 0.023 | 1.133 |
| (adjusted for age 42 mental health) | 1.049 | 0.280 | 1.000 |

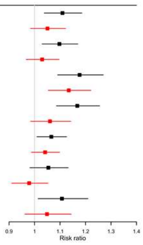

**Diary-derived average**

| Variable (age at collection) | RR | p | E-value |
|---|---|---|---|
| Rutter score (5) | 1.119 | 0.022 | 1.147 |
| (adjusted for age 42 mental health) | 1.079 | 0.110 | 1.000 |
| Rutter score (10) | 1.139 | 0.002 | 1.272 |
| (adjusted for age 42 mental health) | 1.097 | 0.040 | 1.069 |
| Child Development Scale (10) | 1.145 | 0.007 | 1.238 |
| (adjusted for age 42 mental health) | 1.119 | 0.024 | 1.139 |
| Malaise score (16)* | 1.096 | 0.041 | 1.064 |
| (adjusted for age 42 mental health) | 1.035 | 0.466 | 1.000 |
| Behavioural & emotional problems (16) | 1.062 | 0.152 | 1.000 |
| (adjusted for age 42 mental health) | 1.047 | 0.270 | 1.000 |
| Internalising behaviours (16) | 1.070 | 0.076 | 1.000 |
| (adjusted for age 42 mental health) | 1.025 | 0.523 | 1.000 |
| Externalising behaviours (16) | 1.079 | 0.133 | 1.000 |
| (adjusted for age 42 mental health) | 1.044 | 0.395 | 1.000 |

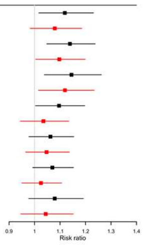

**activPAL algorithm**

| Variable (age at collection) | RR | p | E-value |
|---|---|---|---|
| Rutter score (5) | 0.998 | 0.943 | 1.000 |
| (adjusted for age 42 mental health) | 0.996 | 0.856 | 1.000 |
| Rutter score (10) | 1.016 | 0.425 | 1.000 |
| (adjusted for age 42 mental health) | 1.013 | 0.494 | 1.000 |
| Child Development Scale (10) | 0.992 | 0.730 | 1.000 |
| (adjusted for age 42 mental health) | 0.990 | 0.673 | 1.000 |
| Malaise score (16)* | 0.999 | 0.982 | 1.000 |
| (adjusted for age 42 mental health) | 0.995 | 0.827 | 1.000 |
| Behavioural & emotional problems (16) | 1.032 | 0.096 | 1.000 |
| (adjusted for age 42 mental health) | 1.031 | 0.106 | 1.000 |
| Internalising behaviours (16) | 1.016 | 0.396 | 1.000 |
| (adjusted for age 42 mental health) | 1.013 | 0.515 | 1.000 |
| Externalising behaviours (16) | 1.016 | 0.430 | 1.000 |
| (adjusted for age 42 mental health) | 1.013 | 0.496 | 1.000 |

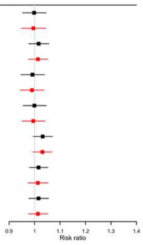

**van der Berg et al. algorithm**

| Variable (age at collection) | RR | p | E-value |
|---|---|---|---|
| Rutter score (5) | 1.022 | 0.352 | 1.000 |
| (adjusted for age 42 mental health) | 1.016 | 0.518 | 1.000 |
| Rutter score (10) | 1.016 | 0.580 | 1.000 |
| (adjusted for age 42 mental health) | 1.010 | 0.730 | 1.000 |
| Child Development Scale (10) | 1.013 | 0.699 | 1.000 |
| (adjusted for age 42 mental health) | 1.010 | 0.779 | 1.000 |
| Malaise score (16)* | 0.998 | 0.929 | 1.000 |
| (adjusted for age 42 mental health) | 0.987 | 0.624 | 1.000 |
| Behavioural & emotional problems (16) | 0.967 | 0.432 | 1.000 |
| (adjusted for age 42 mental health) | 0.964 | 0.383 | 1.000 |
| Internalising behaviours (16) | 1.007 | 0.811 | 1.000 |
| (adjusted for age 42 mental health) | 0.999 | 0.978 | 1.000 |
| Externalising behaviours (16) | 1.032 | 0.265 | 1.000 |
| (adjusted for age 42 mental health) | 1.027 | 0.343 | 1.000 |

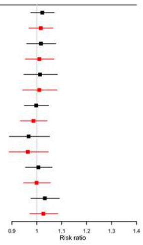

**Winkler et al. algorithm**

| Variable (age at collection) | RR | p | E-value |
|---|---|---|---|
| Rutter score (5) | 1.086 | 0.001 | 1.218 |
| (adjusted for age 42 mental health) | 1.075 | 0.006 | 1.165 |
| Rutter score (10) | 1.016 | 0.480 | 1.000 |
| (adjusted for age 42 mental health) | 1.003 | 0.891 | 1.000 |
| Child Development Scale (10) | 1.099 | 0.004 | 1.211 |
| (adjusted for age 42 mental health) | 1.091 | 0.009 | 1.172 |
| Malaise score (16)* | 1.086 | 0.003 | 1.200 |
| (adjusted for age 42 mental health) | 1.065 | 0.029 | 1.088 |
| Behavioural & emotional problems (16) | 1.012 | 0.586 | 1.000 |
| (adjusted for age 42 mental health) | 1.006 | 0.763 | 1.000 |
| Internalising behaviours (16) | 1.027 | 0.314 | 1.000 |
| (adjusted for age 42 mental health) | 1.011 | 0.709 | 1.000 |
| Externalising behaviours (16) | 1.077 | 0.025 | 1.106 |
| (adjusted for age 42 mental health) | 1.065 | 0.061 | 1.000 |

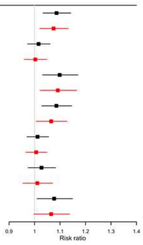

*Figure S6.* Estimated risk ratios quantifying the associations among female participants between the childhood mental health variables (standardised) and the status of sleep duration as abnormally short, normal or abnormally long in adulthood (derived from five separate measures of sleep), also including (in red) those adjusting for Malaise and Warwick–Edinburgh Mental Well-Being Scale scores at age 42. The E-value, quantifying the minimum unmeasured confounding risk ratio that would be needed to nullify each association estimated here, is also shown. Each exposure-outcome relationship was assessed in a separate model. All models were adjusted for a range of socioeconomic, perinatal and health-related covariates. Missing data were handled by multiple imputation. RR = risk ratio. * = self-reported.

### Abnormally short sleep duration

**Self-reported average**

| Variable (age at collection) | RRR | p | E-value |
|---|---|---|---|
| Rutter score (5) | 1.139 | 0.003 | 1.263 |
| Rutter score (10) | 1.123 | 0.005 | 1.226 |
| Child Development Scale (10) | 1.235 | 0.000 | 1.462 |
| Malaise score (16)* | 1.232 | 0.000 | 1.501 |
| Behavioural & emotional problems (16) | 1.068 | 0.148 | 1.000 |
| Internalising behaviours (16) | 1.064 | 0.211 | 1.000 |
| Externalising behaviours (16) | 1.137 | 0.005 | 1.240 |

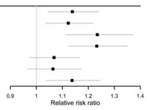

**Diary-derived average**

| Variable (age at collection) | RRR | p | E-value |
|---|---|---|---|
| Rutter score (5) | 1.095 | 0.089 | 1.000 |
| Rutter score (10) | 1.097 | 0.165 | 1.000 |
| Child Development Scale (10) | 1.168 | 0.046 | 1.064 |
| Malaise score (16)* | 1.103 | 0.161 | 1.000 |
| Behavioural & emotional problems (16) | 1.108 | 0.144 | 1.000 |
| Internalising behaviours (16) | 1.053 | 0.383 | 1.000 |
| Externalising behaviours (16) | 1.056 | 0.389 | 1.000 |

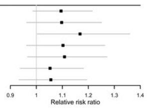

**activPAL algorithm**

| Variable (age at collection) | RRR | p | E-value |
|---|---|---|---|
| Rutter score (5) | 0.989 | 0.861 | 1.000 |
| Rutter score (10) | 1.014 | 0.842 | 1.000 |
| Child Development Scale (10) | 1.024 | 0.746 | 1.000 |
| Malaise score (16)* | 1.043 | 0.485 | 1.000 |
| Behavioural & emotional problems (16) | 1.109 | 0.132 | 1.000 |
| Internalising behaviours (16) | 1.024 | 0.664 | 1.000 |
| Externalising behaviours (16) | 1.033 | 0.581 | 1.000 |

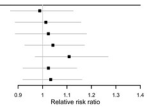

**van der Berg et al. algorithm**

| Variable (age at collection) | RRR | p | E-value |
|---|---|---|---|
| Rutter score (5) | 1.044 | 0.679 | 1.000 |
| Rutter score (10) | 1.089 | 0.361 | 1.000 |
| Child Development Scale (10) | 1.092 | 0.383 | 1.000 |
| Malaise score (16)* | 1.016 | 0.845 | 1.000 |
| Behavioural & emotional problems (16) | 1.024 | 0.752 | 1.000 |
| Internalising behaviours (16) | 1.083 | 0.395 | 1.000 |
| Externalising behaviours (16) | 1.063 | 0.615 | 1.000 |

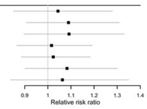

**Winkler et al. algorithm**

| Variable (age at collection) | RRR | p | E-value |
|---|---|---|---|
| Rutter score (5) | 1.131 | 0.037 | 1.098 |
| Rutter score (10) | 1.022 | 0.584 | 1.000 |
| Child Development Scale (10) | 1.167 | 0.007 | 1.256 |
| Malaise score (16)* | 1.125 | 0.003 | 1.247 |
| Behavioural & emotional problems (16) | 1.014 | 0.762 | 1.000 |
| Internalising behaviours (16) | 1.040 | 0.493 | 1.000 |
| Externalising behaviours (16) | 1.115 | 0.041 | 1.077 |

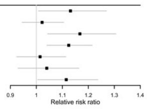

### Abnormally long sleep duration

**Self-reported average**

| Variable (age at collection) | RRR | p | E-value |
|---|---|---|---|
| Rutter score (5) | 1.046 | 0.720 | 1.000 |
| Rutter score (10) | 1.020 | 0.888 | 1.000 |
| Child Development Scale (10) | 1.114 | 0.483 | 1.000 |
| Malaise score (16)* | 1.113 | 0.535 | 1.000 |
| Behavioural & emotional problems (16) | 0.984 | 0.973 | 1.000 |
| Internalising behaviours (16) | 1.112 | 0.520 | 1.000 |
| Externalising behaviours (16) | 1.187 | 0.189 | 1.000 |

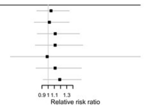

**Diary-derived average**

| Variable (age at collection) | RRR | p | E-value |
|---|---|---|---|
| Rutter score (5) | 1.070 | 0.483 | 1.000 |
| Rutter score (10) | 1.106 | 0.247 | 1.000 |
| Child Development Scale (10) | 1.132 | 0.186 | 1.000 |
| Malaise score (16)* | 1.079 | 0.512 | 1.000 |
| Behavioural & emotional problems (16) | 0.999 | 0.990 | 1.000 |
| Internalising behaviours (16) | 0.972 | 0.728 | 1.000 |
| Externalising behaviours (16) | 0.976 | 0.793 | 1.000 |

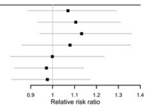

**activPAL algorithm**

| Variable (age at collection) | RRR | p | E-value |
|---|---|---|---|
| Rutter score (5) | 0.990 | 0.781 | 1.000 |
| Rutter score (10) | 1.005 | 0.907 | 1.000 |
| Child Development Scale (10) | 0.993 | 0.852 | 1.000 |
| Malaise score (16)* | 1.016 | 0.747 | 1.000 |
| Behavioural & emotional problems (16) | 1.057 | 0.182 | 1.000 |
| Internalising behaviours (16) | 1.010 | 0.789 | 1.000 |
| Externalising behaviours (16) | 1.003 | 0.936 | 1.000 |

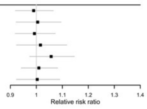

**van der Berg et al. algorithm**

| Variable (age at collection) | RRR | p | E-value |
|---|---|---|---|
| Rutter score (5) | 1.019 | 0.685 | 1.000 |
| Rutter score (10) | 1.030 | 0.463 | 1.000 |
| Child Development Scale (10) | 1.021 | 0.717 | 1.000 |
| Malaise score (16)* | 1.020 | 0.685 | 1.000 |
| Behavioural & emotional problems (16) | 0.991 | 0.850 | 1.000 |
| Internalising behaviours (16) | 1.047 | 0.231 | 1.000 |
| Externalising behaviours (16) | 1.040 | 0.433 | 1.000 |

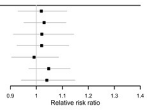

**Winkler et al. algorithm**

| Variable (age at collection) | RRR | p | E-value |
|---|---|---|---|
| Rutter score (5) | 0.985 | 0.819 | 1.000 |
| Rutter score (10) | 1.013 | 0.848 | 1.000 |
| Child Development Scale (10) | 1.005 | 0.958 | 1.000 |
| Malaise score (16)* | 1.084 | 0.338 | 1.000 |
| Behavioural & emotional problems (16) | 0.938 | 0.539 | 1.000 |
| Internalising behaviours (16) | 0.982 | 0.782 | 1.000 |
| Externalising behaviours (16) | 1.026 | 0.757 | 1.000 |

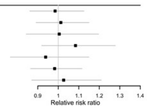

*Figure S7.* Estimated relative risk ratios quantifying the associations among female participants between the childhood mental health variables (standardised) and the presence of abnormally short, and abnormally long, sleep duration in adulthood (derived from five separate measures of sleep). The E-value, quantifying the minimum unmeasured confounding risk ratio that would be needed to nullify each association estimated here, is also shown. Each exposure-outcome relationship was assessed in a separate model. All models were adjusted for a range of socioeconomic, perinatal and health-related covariates. Missing data were handled by multiple imputation. RRR = relative risk ratio. * = self-reported.
